# Clinical investigations to evaluate high-risk orthopaedic devices: systematic review of the peer-reviewed medical literature

**DOI:** 10.1101/2023.08.24.23294479

**Authors:** Anne Lübbeke, Christophe Combescure, Christophe Barea, Amanda Inez Gonzalez, Keith Tucker, Per Kjærsgaard-Andersen, Tom Melvin, Alan G Fraser, Rob Nelissen, James A Smith

## Abstract

**Purpose:** The objective of this systematic review was to give an overview of clinical investigations regarding hip and knee arthroplasty implants published in peer-reviewed scientific medical journals before entry into force of the EU Medical Device Regulation in May 2021.

**Methods:** We systematically reviewed the medical literature for a random selection of hip and knee implants, to identify all peer-reviewed clinical investigations published within 10years before and up to 20years after regulatory approval. We report study characteristics, methodologies, outcomes, measures to prevent bias, and timing of clinical investigations, of 30 current implants. The review process was conducted according to the Preferred Reporting Items for Systematic Reviews and Meta-Analyses (PRISMA) guidelines.

**Results:** We identified 2912 publications and finally included 151 papers published between 1995 and 2021 (63 on hip stems, 34 on hip cups, 54 on knee systems). We identified no clinical studies published before CE-marking for any selected device, and no studies even up to 20 years after CE-marking in one quarter of devices. There were very few randomized controlled trials, and registry-based studies generally had larger sample sizes and better methodology.

**Conclusions:** The peer-reviewed literature alone is insufficient as source of clinical investigations of these high-risk devices intended for life-long use. A more systematic, efficient and faster way to evaluating safety and performance is necessary. Using a phased introduction approach, nesting comparative studies of observational and experimental design in existing registries, increasing use of benefit measures, and accelerating surrogate outcomes research, will help to minimise risks and maximise benefits.

## Introduction

Little is known about the clinical evidence used to establish the safety and performance of medical devices before and after market access in Europe. Unlike medicines in Europe and in the USA, and medical devices that are subject to pre-market authorisation in the USA, there has been no requirement for summaries of clinical evidence to be made publicly available. Under the Medical Device Directive 93/42/EEC (MDD) system, which is still the legal basis for the marketing of the vast majority of medical devices today, it is not possible to identify the clinical evidence supporting device CE-marking (Conformité Européene) as this is considered to be commercially confidential (Article 20 of the MDD). This might be the reason for the very few detailed analyses on evidence for medical device being published.

The Medical Device Regulation ((EU) 2017/745) is changing the requirements for certification (CE-marking) of implantable medical devices in Europe. The MDR will increase transparency of the clinical investigations supporting device CE-marking, by requiring the publication of clinical investigation reports (MDR, Article 77), and it may increase the clinical evidence requirements for some devices. For example, a clinical investigation is required for Class III devices, unless the use of existing clinical data is sufficiently justified. The MDR has also introduced restrictions with respect to the use of data from equivalent devices for the purpose of market entry, with a contract required between manufacturers for high-risk devices (MDR Article 61(5)).

The peer-reviewed medical literature is an established major source of clinical evidence regarding medical devices^1^. In orthopaedic surgery, information derived from the published literature is complemented by annual reports from registries, which monitor real-world safety and performance of implants at national or regional level over the long-term^2^. EU regulatory and health technology assessment bodies have recognized the importance of high-quality registries and wish to optimise their use to generate evidence to support decision-making in clinical practice^3^.

The European Commission has funded the Coordinating Research and Evidence for Medical Devices (CORE-MD) consortium to review and recommend methodologies for the improved clinical investigation and evaluation of high-risk medical devices^4^. An important component for recommending how devices should be evaluated in future is understanding how they have been assessed as well as addressing strengths and limitations of previous evaluation approaches. The aim of the current project is to review the evidence for high-risk orthopaedic devices; the quality and validity of registries are covered elsewhere by the CORE-MD consortium^5^.

Despite changes to the clinical evidence requirements for medical devices under the MDR, a systematic review of studies supporting CE-marking under the MDD is useful for several reasons. Firstly, it will provide better understanding of the availability of published evidence for clinicians and healthcare systems. Secondly, it will provide a useful baseline against which to evaluate the impact of the MDR on clinical investigations and the evidence available in future. Thirdly, it will allow comparison to evidence available for devices in other regulatory environments, which in our project refers specifically to those devices, which have received US FDA market clearance or approval (hereafter clearance).

The objective of this systematic review was to give an overview of clinical investigations regarding hip and knee arthroplasty published in peer-reviewed scientific medical journals, with a focus on methodology and clinically relevant outcomes, before and after regulatory approval (CE-marking).

## Methods

We selected for inclusion a total of 30 hip and knee devices used for primary hip or knee replacement. For each device, we attempted to discover the date of first CE-marking, and we conducted a systematic literature search to identify all published literature available 10 years before and 20 years after introduction of these implants. We identified studies assessing patients who would receive the hip or knee implant under its typical intended use, and we described evidence reported in the studies.

The systematic review is reported according to the relevant items of the Preferred Reporting Items for Systematic Reviews and Meta-Analyses (PRISMA)^6^ statement, and it was registered on the open science framework (https://osf.io/6gmyx).

### Selection of devices (implants) for inclusion in the review

This review aimed to assess a representative sample of CE-marked medical devices. Since a complete list is not available, two sources were used: the Orthopaedic Data Evaluation Panel (ODEP, https://www.odep.org.uk/, accessed 8^th^ June 2021), and European national registries. Consultation with CORE-MD members including regulatory agencies identified ODEP as having one of the most complete lists of hip and knee implants available on the European market. National registries from Denmark, Finland, Germany, Netherlands, Norway, Sweden, Switzerland and the UK were also searched. Merging these two sources, we obtained lists of hip cups (n=138), hip stems (n=165) and knee (n=97) implants. From that pool of CE-marked implants, ten devices were then randomly selected from each of the three lists.

The unit of analysis used was determined for the hip by the implant name and the type of fixation (i.e. cemented or cementless), and for the knee by the implant name and the type of stability, in accordance with ISAR Benchmarking recommendations^7^.

### CE-marking and FDA clearance dates

We identified CE-marking dates by asking ODEP, to which manufacturers often provide them. If unsuccessful, we then searched the internet for press releases, manufacturers’ brochures, or mentions in academic papers that stated the date or that indicated the approximate date.

We searched for the selected medical devices in the FDA medical device databases to establish whether they had FDA clearance and, if so, to record the date of clearance (https://www.accessdata.fda.gov/scripts/cdrh/cfdocs/search/default.cfm).

### Search strategy

For the published literature, we searched Embase through Ovid, PubMed, and Web of Science. All Web of Science core collection editions, apart from Conference Proceedings Citation Index – Science (CPCI-S)--1990-present and Conference Proceedings Citation Index –Social Science & Humanities (CPCI-SSH)--1990-present, were searched. We used the general structure of “Device name” AND “Hip” [or “Knee”] AND “Humans” for all searches. Search results were combined and automatically de-duplicated in Endnote web, and one author (JAS) manually de-duplicated the results before screening for inclusion and exclusion was done. Full details of searches are provided in Appendix II. Searches were limited to 10 years before the CE-marking date and 20 years afterwards. References of relevant systematic reviews were reviewed to identify additional clinical investigations.

### Inclusion and exclusion criteria

We included studies that reported clinical investigations (defined by MDR Article 2(45)) of the devices of interest. We operationalized “undertaken to assess the safety or performance of a device” as: i) the study specifically aimed to assess the device in question using at least one of the safety and performance outcomes of interest (defined below) in the context of usual use of the device; and ii) the outcomes were presented by device. Studies that tested something other than the device were excluded (for example, testing of different wound dressings in two groups which both received the implant of interest).

We included case reports and series, case-control studies, registry-based cohorts, cohort studies, and randomised controlled trials.

The outcomes of interest were:

- All-cause revision, assessed at a specific time point (a count of events without any information about when those events occurred would not be included)
- Assessment of implant migration or periprosthetic osteolysis (recognized surrogate markers for implant failure)
- Assessment of the patient-reported outcomes (PROs)
- Frequency of postoperative orthopaedic complications relevant to arthroplasty (if these were defined as a distinct outcome in the study)

We only included studies describing the results of the selected implants in the context of primary total joint replacement. Studies describing results in the context of revision surgery, after hip fracture only, or in any other unusual subpopulation or in cadavers, and conference abstracts, were excluded.

If more than one paper described the findings of a study then the most comprehensively reported paper was included to avoid duplicate data. Studies written in a language spoken by one of the investigators (English, French, German) were included.

### Data collection and management

Details are provided in Appendix III. We collected information such as CE-mark date, manufacturer, and FDA clearance date, in Microsoft Excel. Data extracted from published literature were documented in a database created for this project in REDCap. Two reviewers (AL and JAS) screened all records, and two reviewers (hip stems and knees: AL and JAS; hip cups: AL and AIG) extracted all data in duplicate, and discussed and consolidated any differences, with the exception of non-English language studies, which were only extracted by the reviewer (AL) who spoke that language.

### Analysis

Characteristics of clinical investigations in the published literature were described in terms of study location, year, study design, methodology and outcomes. We intended to describe investigations performed before and after CE-mark dates separately, but no investigations before CE-marking were identified. We considered studies published up to 2 years after the date of CE-marking or FDA clearance to have been performed pre-CE mark or FDA clearance. Where information was available, we compared studies available pre- and post-FDA clearance.

## Results

Through the literature search, 2901 peer-reviewed publications were identified, and 11 additional papers were found via their references. After de-duplication, in most cases using the full-text, we finally included 151 published between 1995 and 2021, of which 63 were for the 10 hip stems, 34 for the 10 hip cups, and 54 for the 10 knee systems (Appendix I). Table 1 summarises the number of studies identified and included at each stage of the systematic review.

**Table 1.** Literature search results.

| **Implant type** | **Embase** | **PubMed** | **Web of science** | **N before deduplication** | **N after deduplication\*** | **N other sources\*** | **N studies included** |
| --- | --- | --- | --- | --- | --- | --- | --- |
| Hip stem | 408 | 238 | 293 | 939 | 751 | 9 | 63 |
| Hip cup | 199 | 50 | 137 | 386 | 302 | 1 | 34 |
| Knee | 825 | 399 | 352 | 1576 | 1078 | 1 | 54 |
| **Total** | **1432** | **687** | **782** | **2901** | **2131** | **11** | **151** |

\*The sum of these two columns is the total number of articles reviewed for inclusion.

Information on the CE-mark year was found for 28 of the 30 implants (Table 2 and Appendix IV). For those 28, all publications dated after their CE mark (median 9 years later, range 3-13 years). No peer-reviewed publication was found for eight implants (27%), of which one was a hip stem, four were hip cups, and three were knee systems.

**Table 2.** Device names and corresponding pre- and post-market publications.

|  | **Device name** | **N pre-market publications** | **CE-mark year found** | **N post-market publications** |
| --- | --- | --- | --- | --- |
| **Hip Stem** | Accolade II | 0 | Yes | 12 |
|  | Alloclassic Zweymuller SL | 0 | Yes | 19 |
|  | Avenir | 0 | Yes | 4 |
|  | BiContact Cementless | 0 | Yes | 8 |
|  | COLLO-MIS | 0 | Yes | 2 |
|  | C-Stem AMT Total Hip System | 0 | Yes | 2 |
|  | Filler 3ND | 0 | Yes | 1 |
|  | MiniHip | 0 | Yes | 8 |
|  | QUADRA | 0 | Yes | 7 |
|  | Stelia stem | 0 | Yes | 0 |
| **Hip Cup** | ANA.NOVA cup | 0 | Yes | 2 |
|  | aneXys | 0 | Yes | 0 |
|  | Cenator | 0 | Yes | 0 |
|  | EcoFit Cementless | 0 | Yes | 0 |
|  | Exceed ABT Cup | 0 | Yes | 4 |
|  | IP X-LINKed acetabular cup | 0 | Yes | 0 |
|  | Plasmacup SC | 0 | Yes | 9 |
|  | POLARCUP™ Cemented | 0 | Yes | 3 |
|  | RM pressfit Vitamys | 0 | Yes | 8 |
|  | Versafit CC Trio | 0 | Yes | 8 |
| **Knee system** | ACS Unc, Unicondylar | 0 | No | 0 |
|  | balanSys CR | 0 | Yes | 4 |
|  | Innex Gender | 0 | Yes | 0 |
|  | LCS Complete | 0 | Yes | 10 |
|  | Logic PS | 0 | Yes | 4 |
|  | NexGen CR | 0 | Yes | 18 |
|  | Optetrak CR | 0 | Yes | 0 |
|  | Sigma High Performance Partial Knee | 0 | Yes | 3 |
|  | TREKKING CR | 0 | No | 2* |
|  | Vanguard CR | 0 | Yes | 13 |

\*No CE-mark year information for Trekking; identified publications were from 2012 and 2018

### Study characteristics, methodology and outcomes overall and by device group

The majority of studies had been conducted in Europe (64%), (Table 3). This proportion was similar for hip stems, cups and knee systems. On average there were 5 publications (range 0- 19) per implant within the period up to 20 years after the CE mark year. The median time between inclusion of the first patient into a study and publication of the results was 10 years (range 2-22 years).

**Table 3.** Study characteristics and study methodology.

|  | **Hip stems**<br>**(N =63)** | **Hip cups**<br>**(N =34)** | **Knees**<br>**(N =54)** | **All**<br>**(N =151)** |
| --- | --- | --- | --- | --- |
| **Publication period** | 1995-2021 | 2007-2021 | 2002-2021 | 1995-2021 |
| **Location (%)** |  |  |  |  |
| Europe | 66.7 | 70.6 | 61.1 | 63.6 |
| Americas | 23.8 | 0 | 29.6 | 19.9 |
| Asia | 1.6 | 23.5 | 9.3 | 9.3 |
| Other | 7.9 | 11.8 | 1.9 | 5.3 |
| **Study type** |  |  |  |  |
| Case report | 3.2% | 11.8% | 1.9% | 4.6% |
| Case-control | - | - | 5.6% | 2% |
| Cohort registry-based | 7.9% | 11.8% | 18.5% | 12.6% |
| Other cohorts | 84.1% | 67.6% | 59.3% | 71.5% |
| Retrospective* | 83.0% | 56.5% | 62.5% | 72.2% |
| RCT | 4.8% | 8.8% | 14.8% | 9.3% |
| **Comparator group(s) yes** | 41.3% | 23.5% | 59.3% | 43.7% |
| **Adjusted\*\* analysis yes** | 25.4% | 5.9% | 38.9% | 25.8% |
| **N prostheses included,**<br>**mean - median (range)** | 615 - 139<br>(1-14'147) | 613 - 95<br>(1-14'147) | 1460 - 180<br>(1-27'193) | 917 - 139<br>(1-27'193) |
| **Inclusion period, median**<br>**years** | 3 | 2 | 3 | 3 |
| **Follow-up, median years,**<br>**range** | 5.5 (0.1-17.8) | 5.0 (0.3-15.0) | 3.4 (1-13.4) | 4.6 (0.1-17.8) |
| **First inclusion date to**<br>**publication in years,**<br>**median, range** | 10 (4-22) | 9 (2-21) | 11 (3-20) | 10 (2-22) |
| **FDA approval to first**<br>**publication in years,**<br>**median, range** | 5 ((-8)-10) | 2 (1-3) | 5 ((-3)-8) | 5 ((-8)-10) |
| **CE-mark date to first**<br>**publication in years,**<br>**median, range** | 9 (3-13) | 10 (7-12) | 7 (5-10) | 9 (3-13) |

\*Percentage of Other cohorts; \*\* Matching instead of adjustment was used in 1 study

The FDA had approved 16 of the 30 randomly selected implants for use in the US (Appendix IV). Overall, devices had been approved by CE-marking earlier in the EU, at a median interval before approval by the FDA in the USA of 4.6 years (range −1 year to + 17.8 years). In 6 cases regulatory approval was obtained around the same year (within a period of 1 year). On average, the first publication for those hip and knee devices appeared 5 years after approval by the FDA (median interval 5.0 years, range 8 years before to 10 years afterwards).

The median duration of follow-up in the selected studies was 4.6 years, ranging from 0.1 to 17.8 years, and the mean duration was 5.2 years (SD±3.7). Median follow-up was 1.7 years longer in studies evaluating hip prostheses compared to knee implants (Mann-Whitney U test p=0.033). More than half of the hip studies (56% of cup and 52% of stem studies) reported follow-up times between 5 and 17 years, while 37% of the knee studies reported follow-up times between 5 and 13 years (Figure 1 and Table 3).

**Figure 1.**
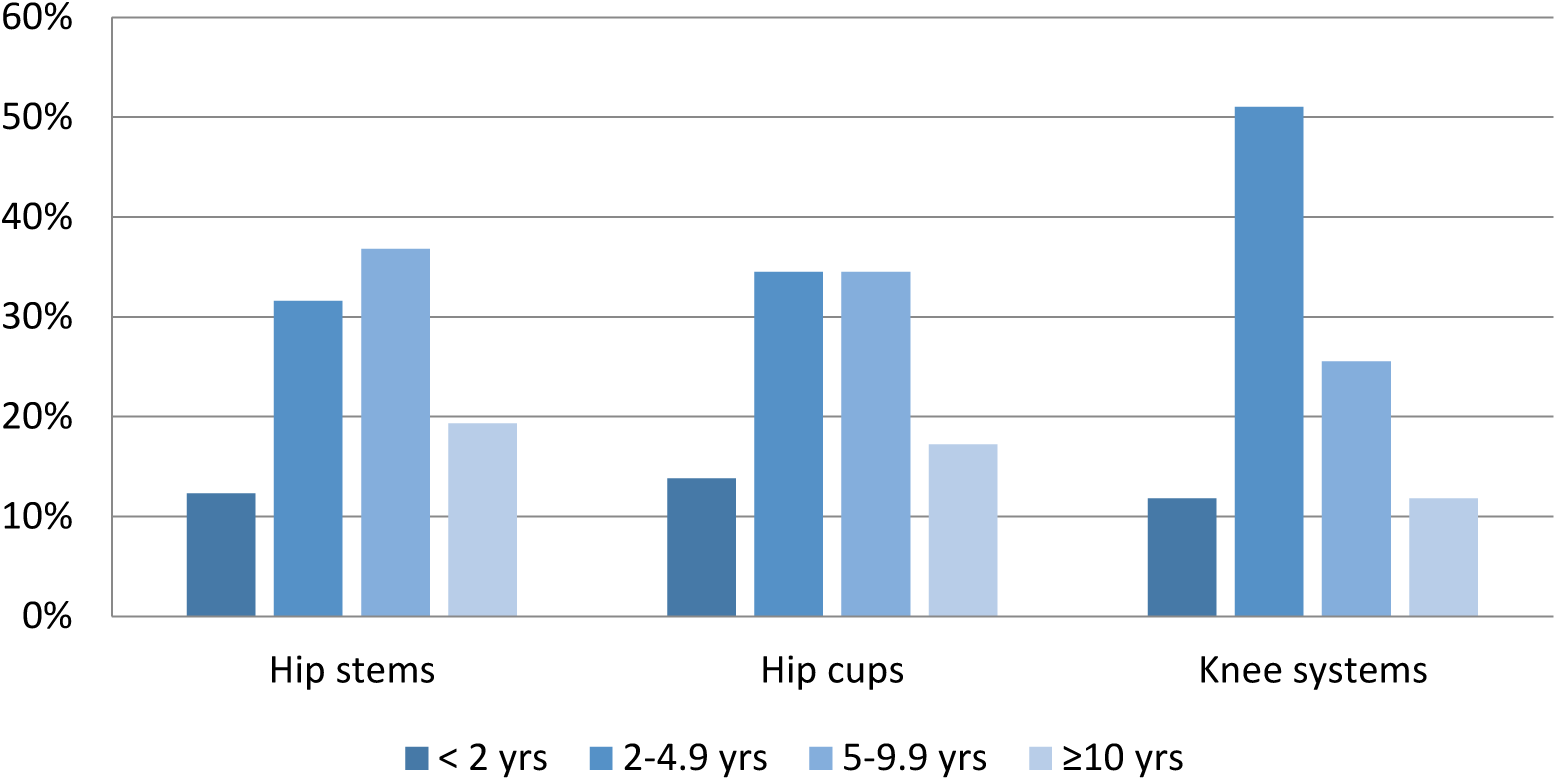
Length of study follow-up by device group.

The median number of implants evaluated in a study (counting only the selected implant, not its comparators) was 139, ranging from 1 to 27,193. Forty-four percent of studies included a comparator group, which was more common for knee than for hip implant studies (59% vs. 35%, Pearson chi-square p=0.004). Regarding study design, the majority were cohort studies (72%), which were mostly retrospective and conducted in one or more academic institutions/hospitals. Adjustment for baseline imbalances in prognostic factors was performed in 26% of studies. Cohort studies based on prospectively collected national or regional registry data made up 13% of the studies. Randomized controlled trials (RCT) constituted 9%. In 6 of the 14 RCTs (43%) blinding of the assessor or the patient was indicated. Knee arthroplasty tended to be more frequently assessed by registry-based cohort studies and RCTs, than were hip arthroplasty devices (Fisher’s exact test p=0.085 and Pearson chi-square p=0.08, respectively; Table 3).

The mean age of subjects (in all studies taken together) was 63 years (range 24-88 years). Women represented 55%, and in 80% of the participants the diagnosis was primary osteoarthritis (OA). Demographics differed between hip and knee arthroplasty patients (Table 4).

**Table 4.** Patient characteristics.

|  | **Hip stems**<br>**(N =63)** | **Hip cups**<br>**(N =34)** | **Knees**<br>**(N =54)** | **All**<br>**(N =151)** |
| --- | --- | --- | --- | --- |
| **Patient characteristics** |  |  |  |  |
| **Age, years, mean (range)** | 60.6 (41-88) | 60.6 (24-75) | 68 (54-77) | 63.3 (24-88) |
| **Women (%), mean (range)** | 50.1 (0-100) | 46.9 (0-100) | 66.8 (10-100) | 55.3 (0-100) |
| **Primary OA (%), mean (range)** | 73.8 (15-100) | 68.3 (0-100) | 94.5 (78-100) | 79.5 (0-100) |
| **Mortality (%), mean - median ( range)** | 11.2 - 7.8 (0-42.8) | 6 - 2.2 (0-30.3) | 7.7 - 0.9 (0-44.2) | 8.7 - 3 (0-44.2) |
| **Lost-to-follow-up (%), mean - median (range)** | 6.4 - 5 (0-22.1) | 5.8 - 4.7 (0-21.1) | 6.2 - 5.5 (0-23.4) | 6.3 - 5 (0-23.4) |

Complete information on the devices used – including cup-stem combination, fixation of the combination, and bearing surface, for the hip; and stability, mobility, fixation, and patella resurfacing, for the knee – was found in 32% of the publications. Information was incomplete in 52%, and no information other than the device name was reported in 16%.

The most frequently reported outcome was all-cause revision (74% of studies), followed by orthopaedic complications (73%), and by imaging results (72%) (Table 5). Complications recorded were prosthetic joint infection, dislocation, or periprosthetic fracture, or else a thromboembolic event or myocardial infarction. Occurrence of these complications overall and by device group is detailed in Table 5. Patient-reported outcomes were assessed in 36% of the studies. There were fewer imaging results reported in knee as compared to hip (stem and cup combined) studies (56% vs. 80%, Pearson chi-square p=0.001), and more functional outcomes in knee studies (59% vs. 2%, Pearson chi-square p<0.001).

**Table 5.** Outcomes reported.

|  | Hip stems<br>(N =63) | Hip cups<br>(N =34) | Knees<br>(N =54) | All<br>(N =151) |
| --- | --- | --- | --- | --- |
| **Type of outcome reported** |  |  |  |  |
| All-cause revision | 81% | 67.6% | 70.4% | 74.2% |
| N revisions reported,<br>median, range | 4.5 (0-440) | 1.5 (0-440) | 3 (0-437) | 4 (0-440) |
| Time-to-event analysis<br>(95%CI) | 25.4% | 29.4% | 33.3% | 29.1% |
| Patient-reported outcomes | 23.8% | 44.1% | 46.3% | 36.4% |
| Imaging | 77.8% | 85.3% | 55.6% | 71.5% |
| RSA study | 8.3% | 5.9% | 9.3% | 7.3% |
| Functional measures | 1.6% | 2.9% | 59.3% | 22.5% |
| Complications (excl. revision) | 79.4% | 73.5% | 66.7% | 73.5% |
| Reported complications %, mean, median (range) |  |  |  |  |
| Prosthesis infection (%) | 0.9 - 0.8 (0-2.4) | 0.5 - 0.1 (0-2.9) | 0.6 - 0.4 (0-2.1) | 0.7 - 0.7 (0-2.9) |
| Dislocation (%) | 1.5 - 0.9 (0-8.2) | 5.5 - 1.0 (0-100) | 0 - 0 (0-0.4) | 2.2 - 0 (0-100) |
| Fracture (%) | 8.3 - 1.6 (0-100) | 1.3 - 1.1 (0-4.4) | 0.2 - 0 (0-1.8) | 4.4 - 0.5 (0-100) |
| Thromboembolic event (%) | 2.5 - 1.9 (0-8) | 1.6 - 0 (0-4.7) | 1.0 - 0.2 (0-4.8) | 1.9 - 1.4 (0-8) |
| Myocardial infarction (%) | 0.3 - 0 (0-1.2) | 0 - 0 (0) | 0 - 0 (0) | 0.2 - 0 (0-1.2) |
| **“Concern” reported in study** |  |  |  |  |
| No concern expressed | 87.3% | 82.4% | 90.7% | 87.4% |
| Potential | 4.8% | 11.7% | 7.4% | 7.3% |
| Yes | 7.9% | 5.9% | 1.9% | 5.3% |
| **“Concern” yes/potential based on** |  |  |  |  |
| Imaging/Revision/PROs/Other | 38/25/0/38% | 78/11/0/11% | 0/60/40/0% | 46/27/9/18% |

RSA=Radiostereometric analysis

A safety concern or an inferior result as compared to another group on one of the outcomes was clearly expressed in 5%, and a potential concern in another 7% of the studies (Table 5). In hip arthroplasty studies it was most frequently based on imaging results (especially radiographs), whereas in knee arthroplasty it was based mostly on revision rates and PROs.

### Study methodology and outcomes by device name

There were large variations between implants in sample size, follow-up period, study methodology, and outcomes, for the published studies (Table 6). For 10 of the 30 implants we found no comparative study, and for 12 no prospective study. For 11 implants, no study reporting on PROs was found. Comparative PRO information was published for 12 implants (40%). Information on revision rates was missing for the 8 implants with no post-market publication. Comparative revision rates including reporting of cumulative failure or survival and 95% CIs were available for 11 implants (37%).

**Table 6.**
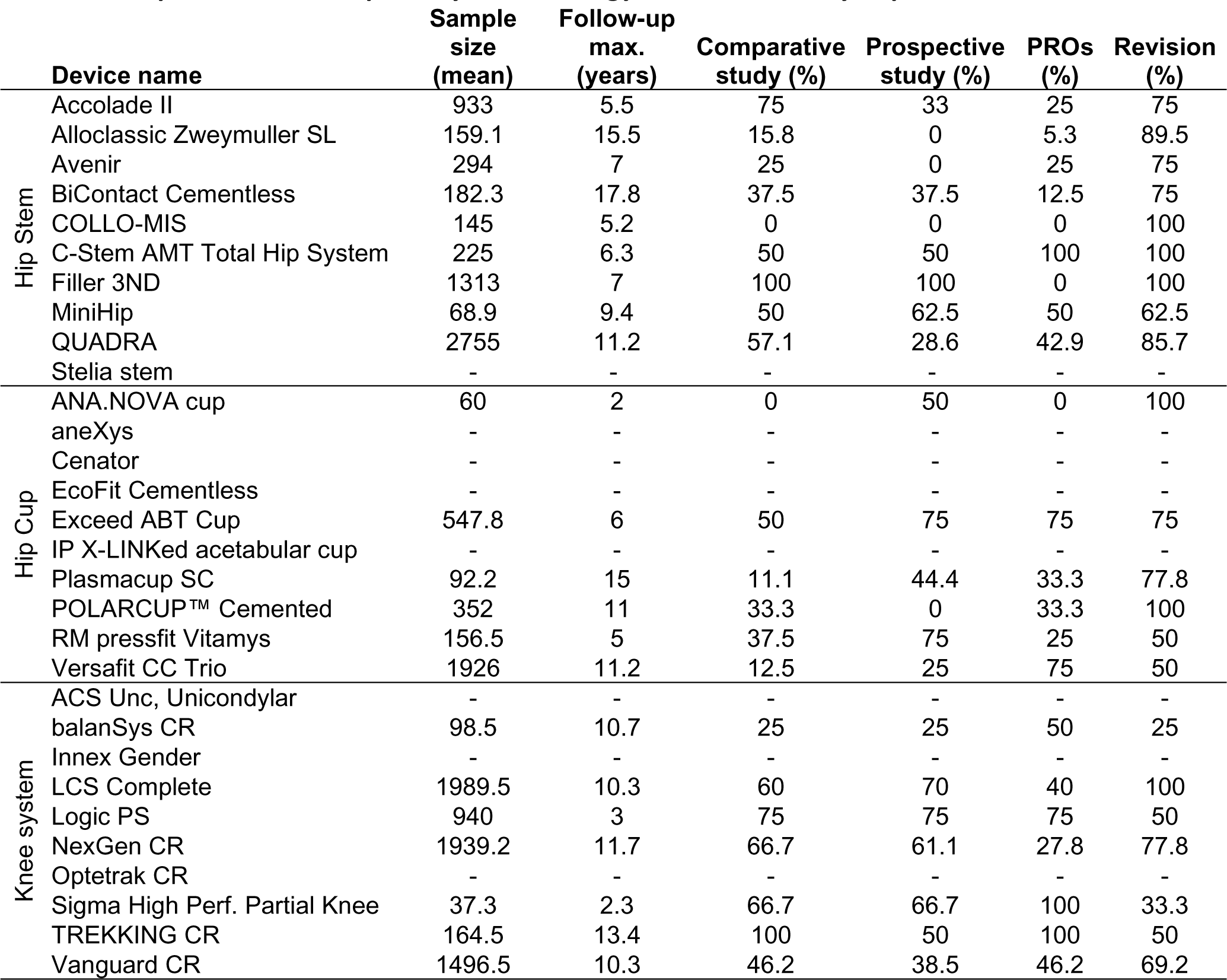
Sample size, follow-up, study methodology and outcomes by implant.

|  | **Device name** | **Sample size (mean)** | **Follow-up max. (years)** | **Comparative study (%)** | **Prospective study (%)** | **PROs (%)** | **Revision (%)** |
| --- | --- | --- | --- | --- | --- | --- | --- |
| Hip Stem | Accolade II | 933 | 5.5 | 75 | 33 | 25 | 75 |
|  | Alloclassic Zweymuller SL | 159.1 | 15.5 | 15.8 | 0 | 5.3 | 89.5 |
|  | Avenir | 294 | 7 | 25 | 0 | 25 | 75 |
|  | BiContact Cementless | 182.3 | 17.8 | 37.5 | 37.5 | 12.5 | 75 |
|  | COLLO-MIS | 145 | 5.2 | 0 | 0 | 0 | 100 |
|  | C-Stem AMT Total Hip System | 225 | 6.3 | 50 | 50 | 100 | 100 |
|  | Filler 3ND | 1313 | 7 | 100 | 100 | 0 | 100 |
|  | MiniHip | 68.9 | 9.4 | 50 | 62.5 | 50 | 62.5 |
|  | QUADRA | 2755 | 11.2 | 57.1 | 28.6 | 42.9 | 85.7 |
|  | Stelia stem | - | - | - | - | - | - |
| Hip Cup | ANA.NOVA cup | 60 | 2 | 0 | 50 | 0 | 100 |
|  | aneXys | - | - | - | - | - | - |
|  | Cenator | - | - | - | - | - | - |
|  | EcoFit Cementless | - | - | - | - | - | - |
|  | Exceed ABT Cup | 547.8 | 6 | 50 | 75 | 75 | 75 |
|  | IP X-LINKed acetabular cup | - | - | - | - | - | - |
|  | Plasmacup SC | 92.2 | 15 | 11.1 | 44.4 | 33.3 | 77.8 |
|  | POLARCUP™ Cemented | 352 | 11 | 33.3 | 0 | 33.3 | 100 |
|  | RM pressfit Vitamys | 156.5 | 5 | 37.5 | 75 | 25 | 50 |
|  | Versafit CC Trio | 1926 | 11.2 | 12.5 | 25 | 75 | 50 |
| Knee system | ACS Unc, Unicondylar | - | - | - | - | - | - |
|  | balanSys CR | 98.5 | 10.7 | 25 | 25 | 50 | 25 |
|  | Innex Gender | - | - | - | - | - | - |
|  | LCS Complete | 1989.5 | 10.3 | 60 | 70 | 40 | 100 |
|  | Logic PS | 940 | 3 | 75 | 75 | 75 | 50 |
|  | NexGen CR | 1939.2 | 11.7 | 66.7 | 61.1 | 27.8 | 77.8 |
|  | Optetrak CR | - | - | - | - | - | - |
|  | Sigma High Perf. Partial Knee | 37.3 | 2.3 | 66.7 | 66.7 | 100 | 33.3 |
|  | TREKKING CR | 164.5 | 13.4 | 100 | 50 | 100 | 50 |
|  | Vanguard CR | 1496.5 | 10.3 | 46.2 | 38.5 | 46.2 | 69.2 |

### Comparison of study methodology and outcomes in registry-based studies vs not

There were large differences in sample size, reported methodology and outcomes, between cohort studies that were based in registries and those that were not (Figure 2). The median numbers of prostheses were 3341 and 149, respectively, and median numbers of revision events were 102 and 3. Studies based in registries more often were prospective, had a comparison group, had more precise reporting of all-cause revision reporting, and more often adjusted analyses. The variety of outcomes assessed was lower in registry-based than in other types of studies.

**Figure 2.**
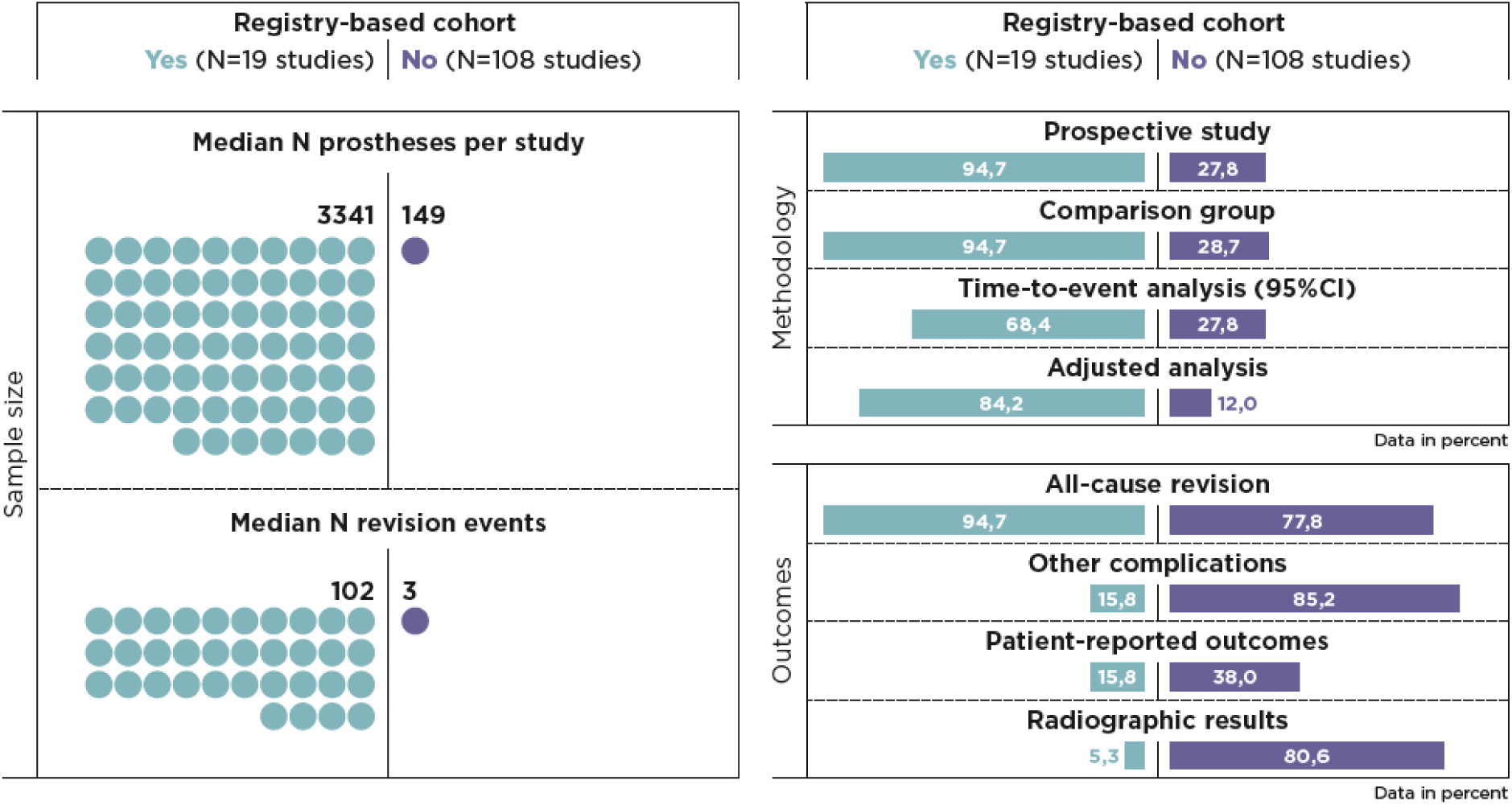
Comparison of cohort studies according to whether or not they were conducted within a registry (median follow-up 5 years for both)

### Trends in study methodology and outcomes

Temporal trends in selected characteristics and outcomes are shown in Figures 3 and 4, combining data from hip and knee arthroplasty studies. There was an increase in comparative, prospective, and registry-based RCTs and radiostereometric analysis (RSA) studies, in particular between the first period (1995-2003) and the second period (2004- 2012). The largest increase was in the reporting of PROs, from 0 in the first to 46% in the third period (2013-2021). There was a substantial decrease (from 94% to 64%) in the reporting of radiographic results.

**Figure 3.**
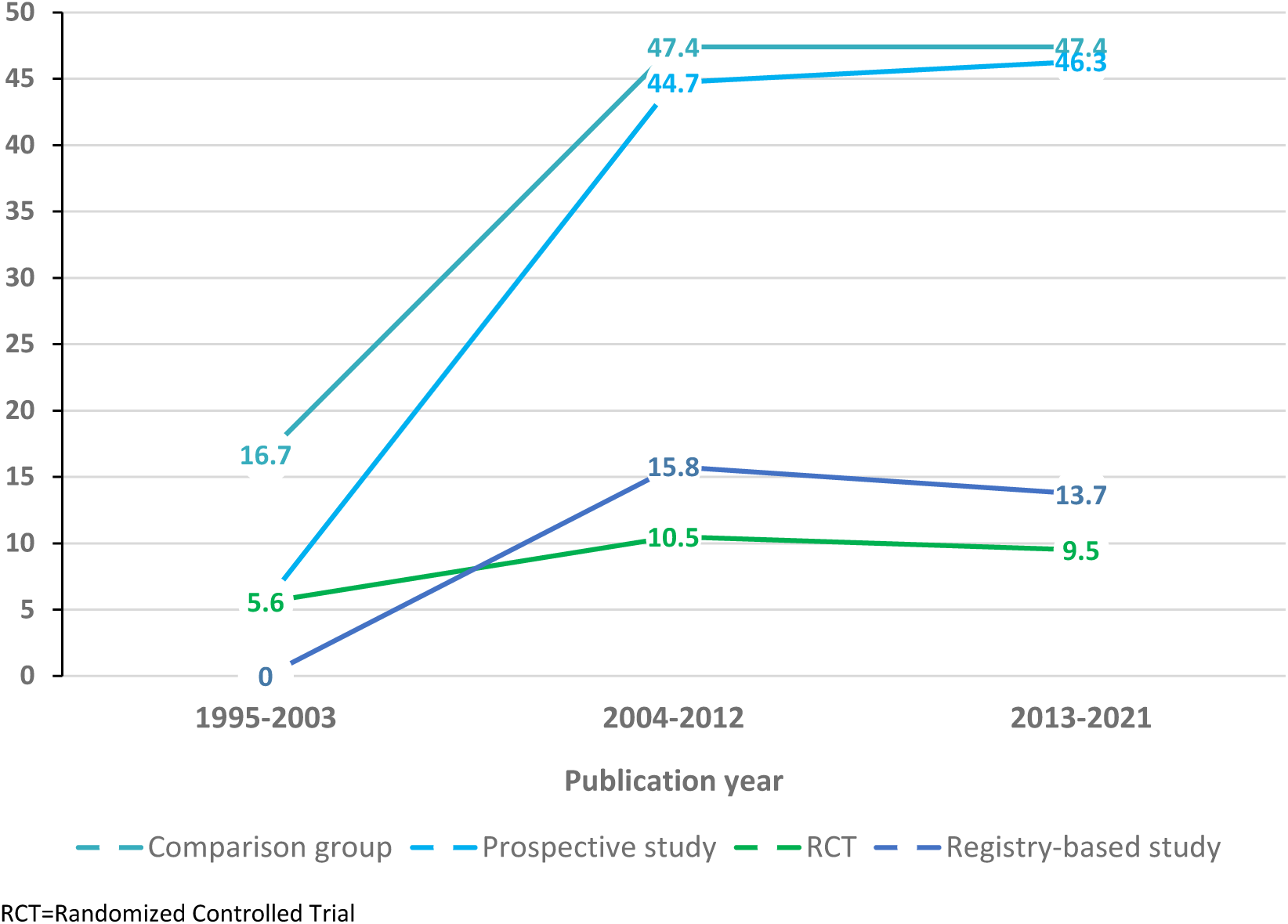
Trends in reported study methodologies for hip and knee implants 1995-2021 (% per period)

**Figure 4.**
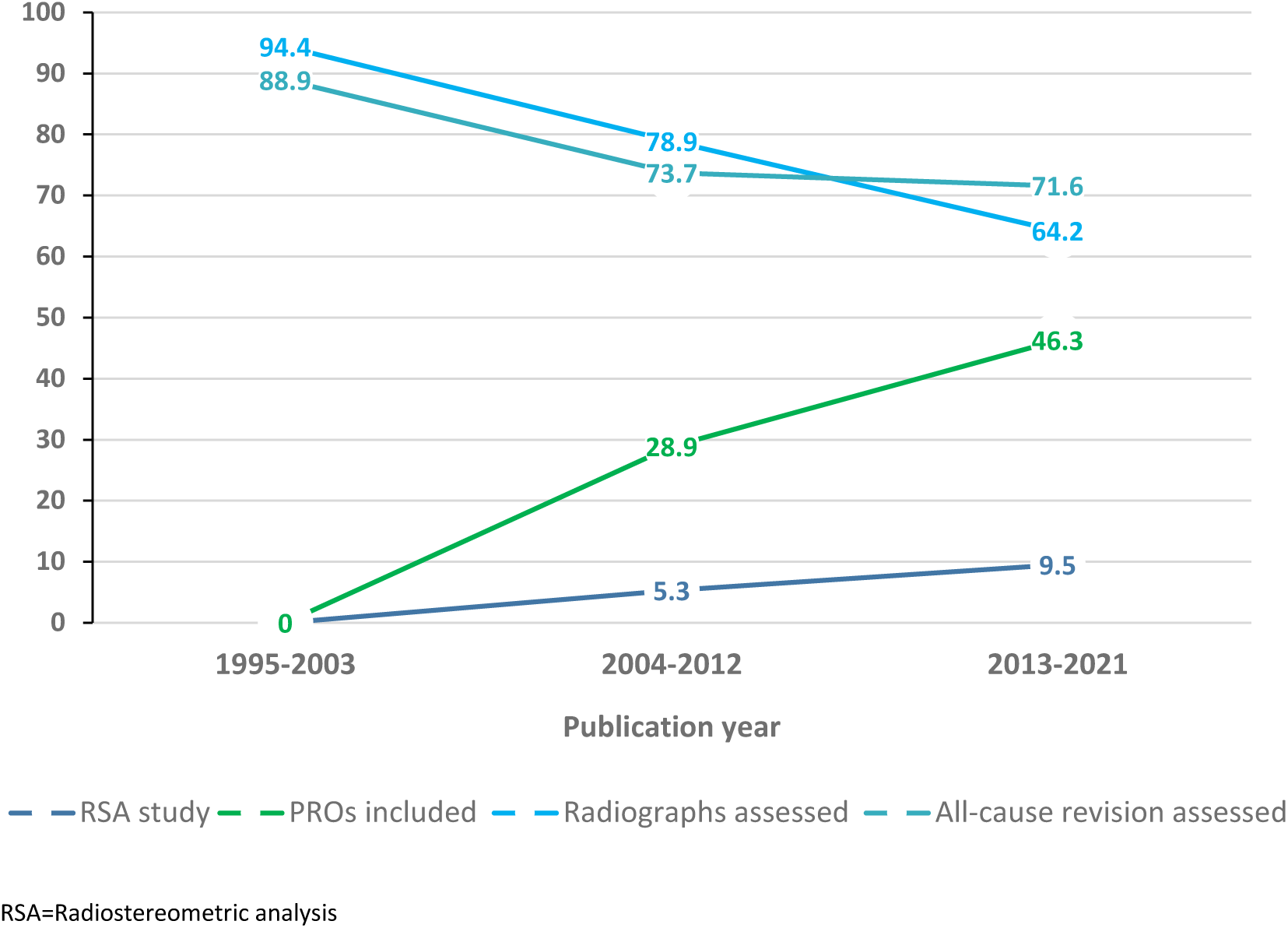
Trends in reported study outcomes on hip and knee implants 1995-2021 (% per period)

## Discussion

This systematic review reports study characteristics, methodologies, outcomes, and timing of clinical investigations in relationship to the CE-marking of high-risk medical devices in orthopaedics (hip and knee implants) before entry into force of the EU MDR in May 2021. We identified no clinical studies published before CE-marking for any selected device and no studies, even up to 20 years after CE marking, in one quarter of devices. There were very few randomized controlled trials, and registry-based studies generally had larger sample sizes and better methodology.

Previous systematic reviews of hip and knee arthroplasty implants largely corroborate our findings. The lack of evidence in 27% of the hip and knee implants in our review is very similar to the proportions reported in publications from the UK (24%), Norway (30%), and Catalonia (23%)^8–10^. The absence of clinical studies published before CE-marking reflects the regulatory situation under the former MDD (93/42/EEC) and confirms literature focusing on medical devices in general^11–12^. Our finding that RCTs were done to assess only 9% of these hip and knee implants, is identical to results from an evaluation of evidence available for implants used in Norway between 1996 and 2000^9^ and to the review of levels of evidence of studies published in major orthopedic journals^13^. The observed absolute and proportional increases in reporting of PROs in our study are in accordance with Siljander et al.^14^ who found an increase from 21% in 2004 to 48% in 2016 in arthroplasty publications in four major orthopedic journals.

### Lack of premarket evidence

The lack of evidence published before CE-marking that was observed in this review is consistent with other studies^12^. Several calls have been made for more evidence to be available before regulatory approval, and particularly for high-risk devices for which alternatives are available, higher evidence requirements would inform better clinical decisions. Limited pre-market evidence might sometimes be acceptable, if complemented by appropriate post-market studies for similar devices, but that should not be commonplace as in the past. For several devices, however, we found neither pre- nor post-market published studies. Considering the high revision rates of some devices, a phased introduction of new implants is paramount to assure optimal safety^15^.

### Post-market evidence and its adequacy

Post-market clinical follow-up (PMCF) studies must resolve questions that are unanswered at the time of regulatory approval, regarding clinical benefit throughout the expected lifetime of a device, its safety under widespread use, the generalisability of pre-market findings, and the continuing acceptability of its benefit-risk ratio. Under the MDR, post-market surveillance is expected to be proactive and continuous, with clinically meaningful comparator(s) and clinically relevant endpoints (risks and benefits). The evidence identified in this review would often not have met those expectations. For 27% of implants we found no published post-market evidence. Comparative studies reporting on PROs were missing for 60% of the implants, and comparative studies reporting cumulative failures or survival rates (with 95%CI) were missing for about two-thirds of the implants.

Of the outcomes included in our review, all-cause revision is the main performance indicator (and risk) of hip and knee arthroplasty, in published PMCF studies. Unless a study was nested in a registry (which was the case in 13% of those in our review) the number of revisions in the evaluated publications was generally low. A challenge with revision as a clinically relevant outcome is that the evaluation of implant longevity requires at least 5 years of follow-up, followed by re-evaluations at regular intervals^16^. This explains the long follow-up times of about half of the studies in this review.

To reduce the duration of follow-up needed before a new implant can be marketed, an alternative clinically meaningful endpoint should be used in early clinical evaluations. Recognized surrogate outcomes that predict the effect of a therapeutic intervention for long-term implant failure are based on imaging, using RSA, Einzel-Bild-Röntgen-Analyse (EBRA), or another similar validated radiographic analysis of implant migration^17–18^. A majority of the reviewed hip studies (>85%) and two-thirds of the knee studies reported either radiographic or RSA results. This confirms that there is an important role for academic institutions to evaluate new implants compared to a standard legacy device, before their market approval. Studies to estimate the risk of revision will assess implant migration and osteolysis on radiographs, and other surrogate markers.

Recognized measures of benefit include patient-reported outcomes, which were assessed in half of the more recent studies selected for this review. Another way of measuring benefit is clinician-reported scores, which most of the earlier studies reported. Collection of PROs was more frequent in non-registry-based than in registry-based studies, but collection of PROs in registries has greatly increased over the past decade. Currently, 16 out of 25 arthroplasty registries worldwide record PROs^19^.

“Traditional” (non-registry-based) follow-up studies alone are unable to document either clinical benefit throughout the expected lifetime of an implant, or its safety under widespread use, because those tasks require much larger sample sizes, more comparators, longer follow-up, and real-world results. Registries or large observational population-based cohorts are better because they generate high-quality post-market clinical evidence for legacy and new devices faster and more efficiently^2,20–23^. They are now recognized by regulators as a preferred source and platform for post-market surveillance and clinical studies^24^. Randomized trials using highly accurate methodology such as implant migration analysis in small studies of up to 50 patients per group, and observational studies including large numbers of patients, should both be nested within registries^25–26^. These studies should be independent, transparent and of high quality^27^. This will require more resources for registries or alternative funding schemes.

### Limitations

There are several limitations of this study. Firstly, we focused on the peer-reviewed medical literature as the source of information about clinical investigations of 30 selected hip and knee implants. There are other publicly available sources, such as annual registry reports, so our findings likely under-represent the total available evidence for the studied implants. Secondly, we limited the outcomes that were included, so the identified papers do not necessarily represent all those investigating a given implant. Thirdly, we constructed and sampled from a list of medical devices that is unlikely to be exhaustive, because there is no list of CE-marked devices currently available. This means that the reported averages in our study refer to the random sample of our list of hip cups, stems and knee implants, but not to all CE-marked hip and knee implants, or to other devices such as shoulders. The sources that we used to identify devices (ODEP and registries) preferentially include those that are used in practice, and we would expect such devices generally to have more evidence available for them than is available for those that are used less often. If so, then the included sample may have had more evidence available than would be found for a sample of all CE-marked devices.

## Conclusions

There is a common perception that more clinical evidence is needed for high-risk medical devices before they are approved for implantation into patients within the EU. One of the goals of the new EU regulation is to achieve that, and one of the objectives of the CORE-MD project is to identify if that will require more clinical studies, better designed clinical trials, better use of real-world data from high-quality registries, and/or more transparency of the results of clinical investigations. Our systematic review suggests that all those measures will be required.

Publication on the EUDAMED portal of a Summary of Safety and Clinical Performance (SSCP) for each new high-risk device will make clinical evidence available at the time that it is approved, instead of many years later when the first paper appears. Since the peer-reviewed literature provides insufficient evidence from clinical investigations of high-risk devices, a more systematic, efficient and faster approach to evaluating safety and performance is necessary. Performing randomized studies in small groups of patients using imaging should detect badly or underperforming orthopaedic implants, before CE-marking. After market approval, nesting studies of observational and experimental design within existing registries or cohorts, increasing the use of benefit measures, and accelerating surrogate outcomes research, would optimize an implant’s benefit-risk ratio.

## Data Availability

All data produced in the present work are contained in the manuscript

## Acknowledgements

We are grateful for review and input to the study protocol from CORE-MD consortium members, in particular: Stephan Windecker, André Frenk, Gearoid McGauran, and Perla J. Marang-van de Mheen. We would like to thank Olga Taylor from ODEP for her assistance.

## Appendix I: list of included devices and corresponding manufacturers

**Hip stems:**

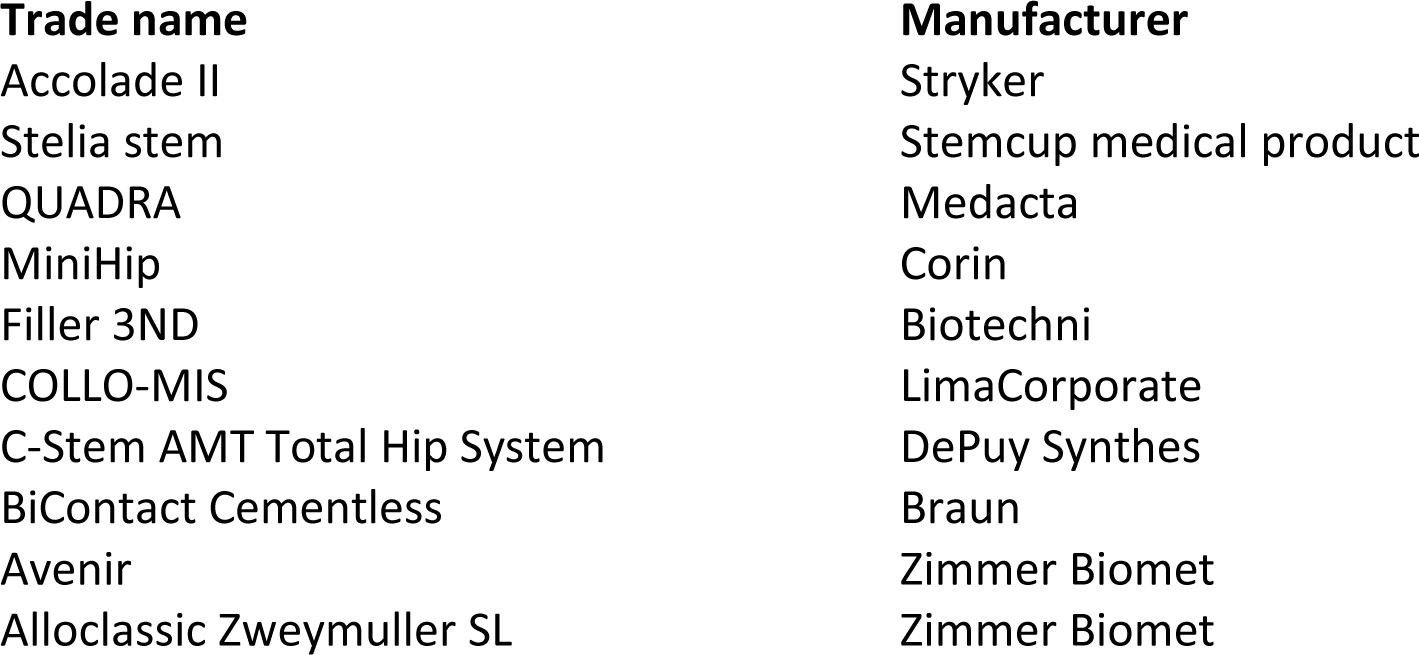

Appendix I: list of included devices and corresponding manufacturers Hip stems:
| Trade name | Manufacturer |
| --- | --- |
| Accolade II | Stryker |
| Stelia stem | Stemcup medical product |
| QUADRA | Medacta |
| MiniHip | Corin |
| Filler 3ND | Biotechni |
| COLLO-MIS | LimaCorporate |
| C-Stem AMT Total Hip System | DePuy Synthes |
| BiContact Cementless | Braun |
| Avenir | Zimmer Biomet |
| Alloclassic Zweymuller SL | Zimmer Biomet |

**Hip cups:**

|  |  |
| --- | --- |
| Versafit CC Trio | Medacta |
| RM pressfit Vitamys | Mathys |
| POLARCUP™ Cemented | Smith & Nephew |
| Plasmacup SC | Braun |
| IP X-LINKed acetabular cup | Waldemar Link |
| Exceed ABT Cup | Zimmer Biomet |
| EcoFit Cementless | Implantcast |
| Cenator | Corin |
| aneXys | Mathys |
| ANA.NOVA cup | Implantec |

**Knee systems:**

|  |  |
| --- | --- |
| LCS Complete | DePuy Synthes |

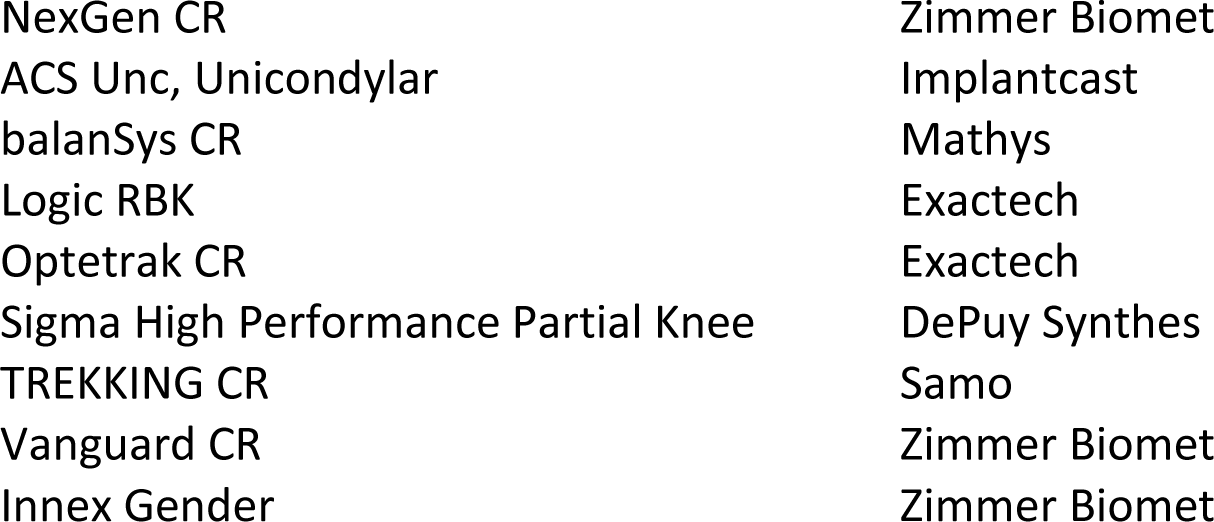

## Appendix II: searches conducted

This document lists all the searches conducted for each device with screenshots showing exactly what was entered into the search database. The title of each section contains the device name and the date on which the search was conducted. The following table summarises the information by device.

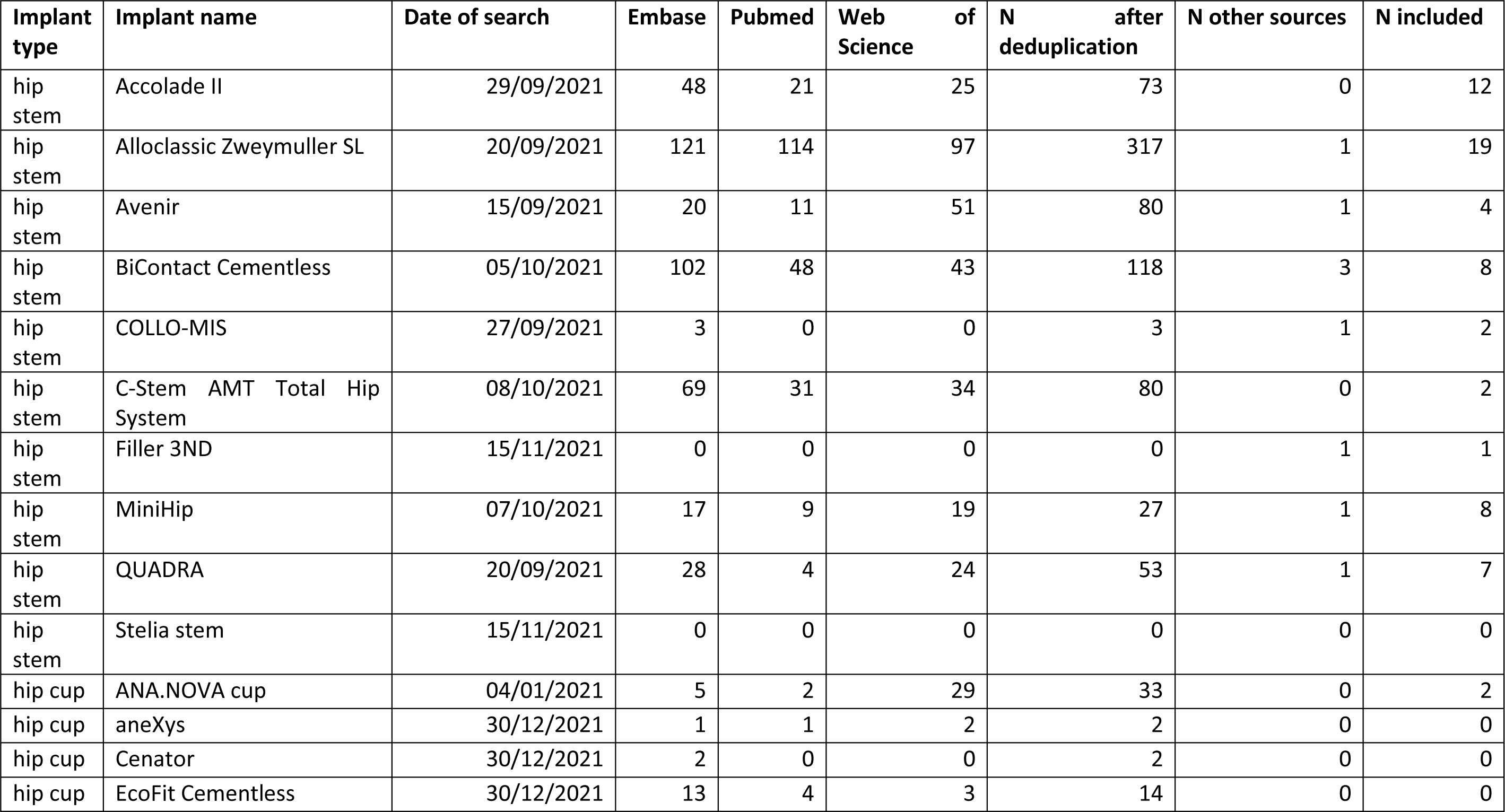

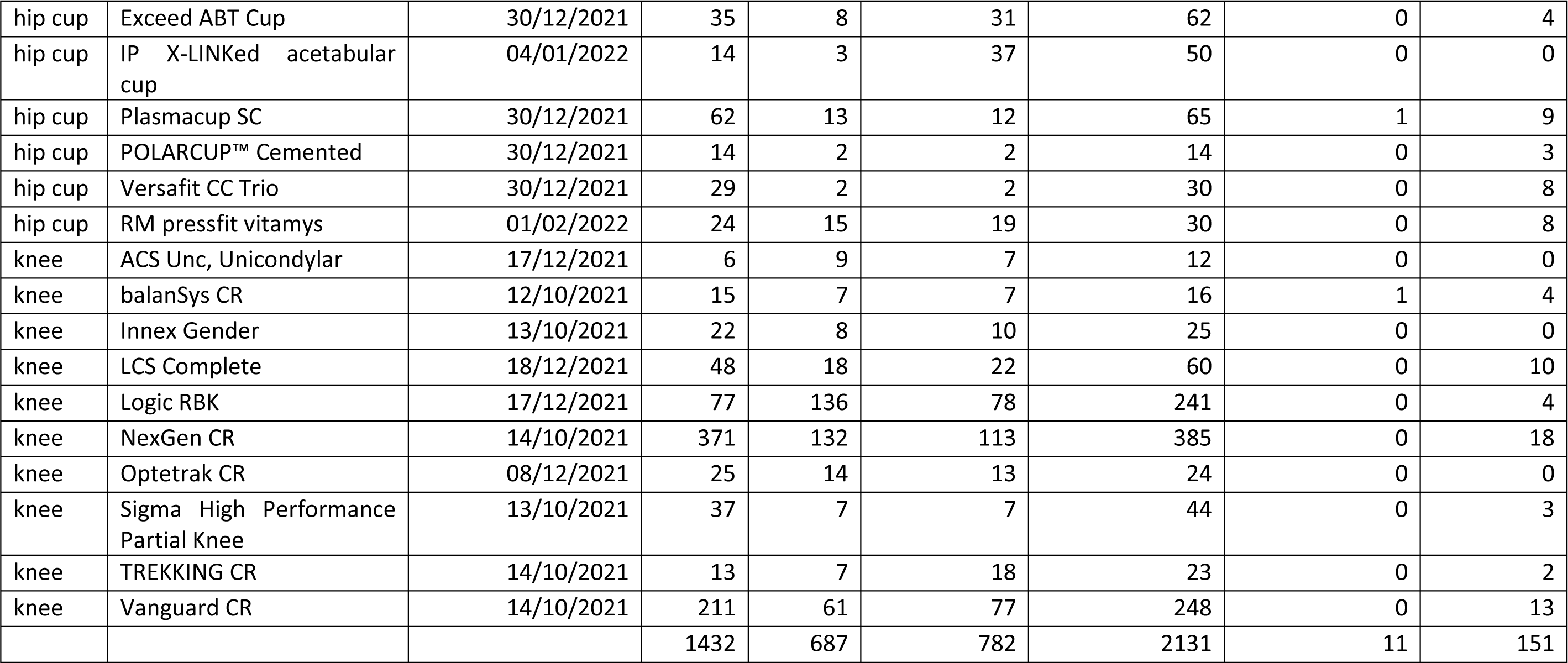

### Hip stem searches

#### Quadra 20^th^ September 2021

Embase: 28

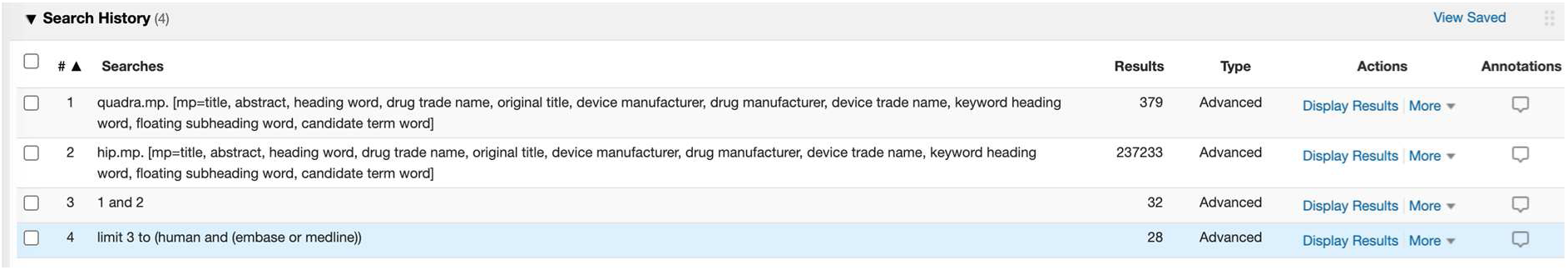

PubMed: 4

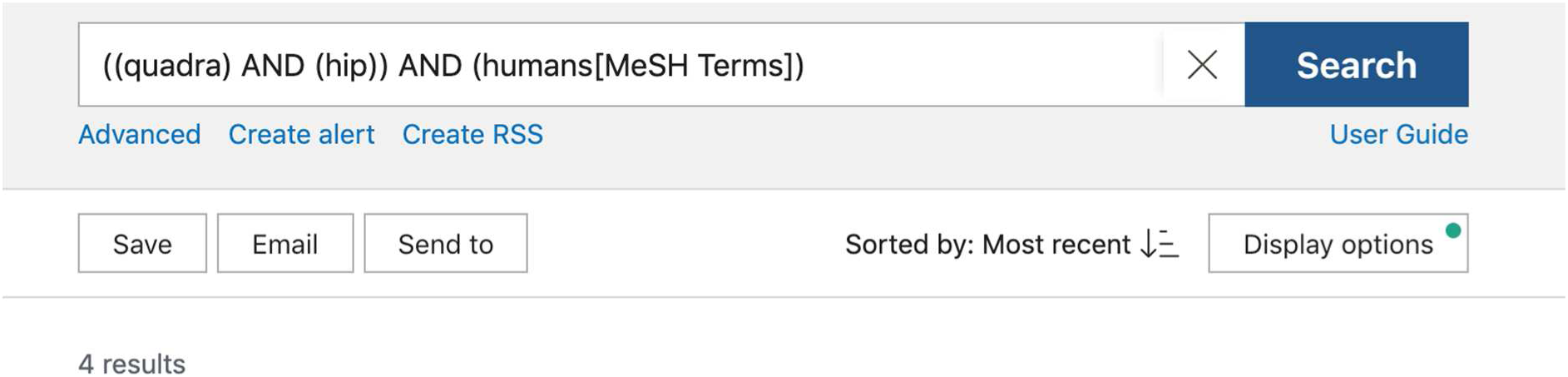

Web of Science: 24

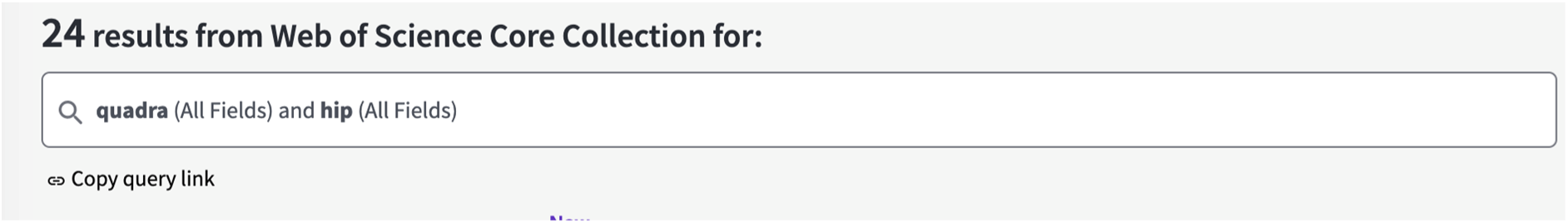

#### Avenir 15^th^ September 2021

Embase: 20

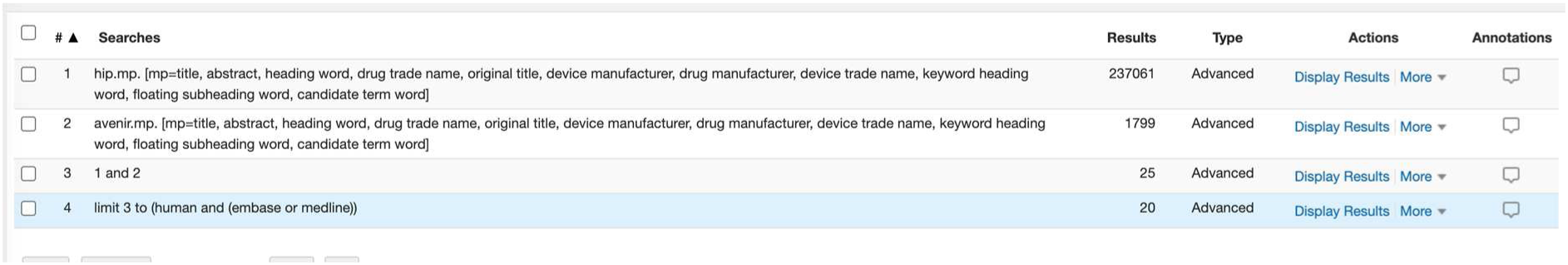

PubMed: 11

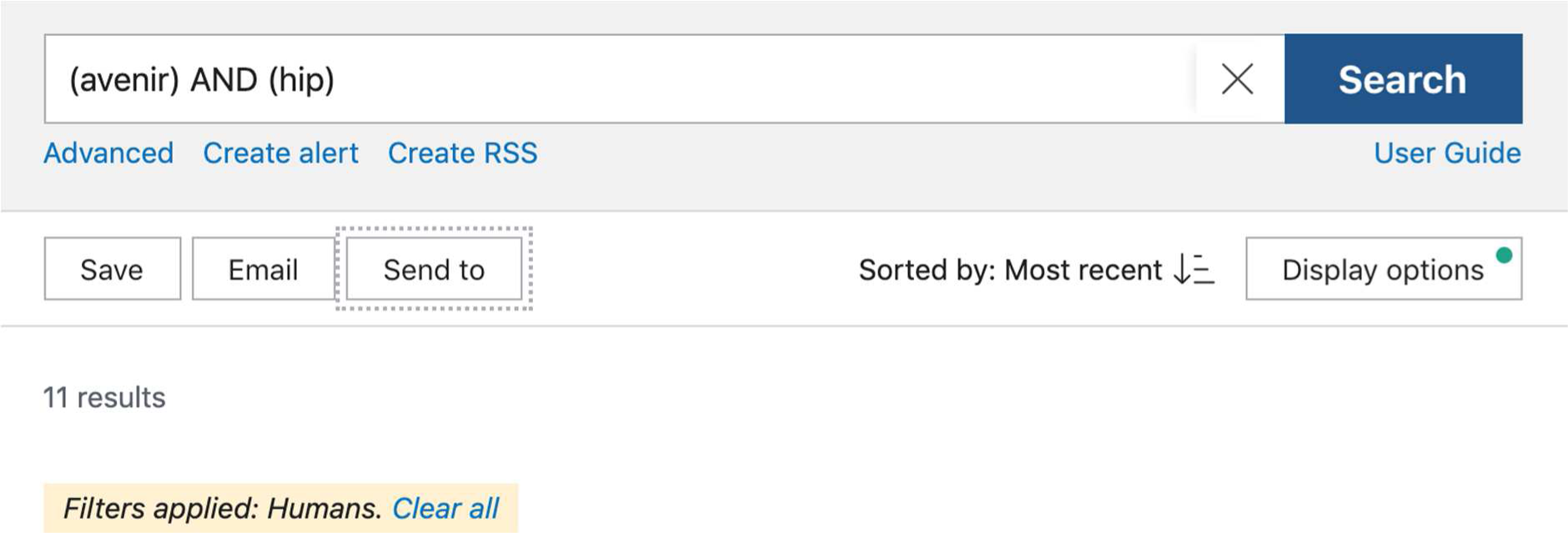

Web of Science

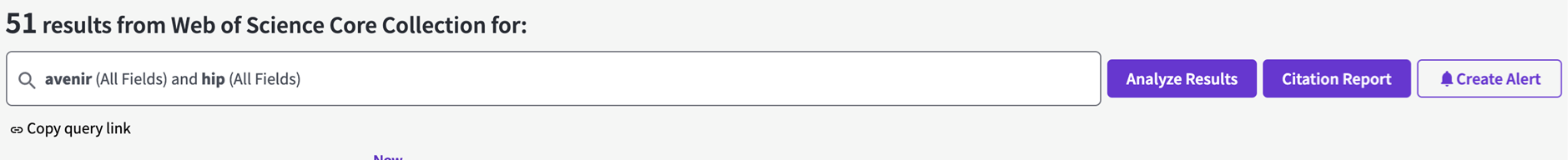

#### Alloclassic Zweymuller 20^th^ September 2021

Embase: 121

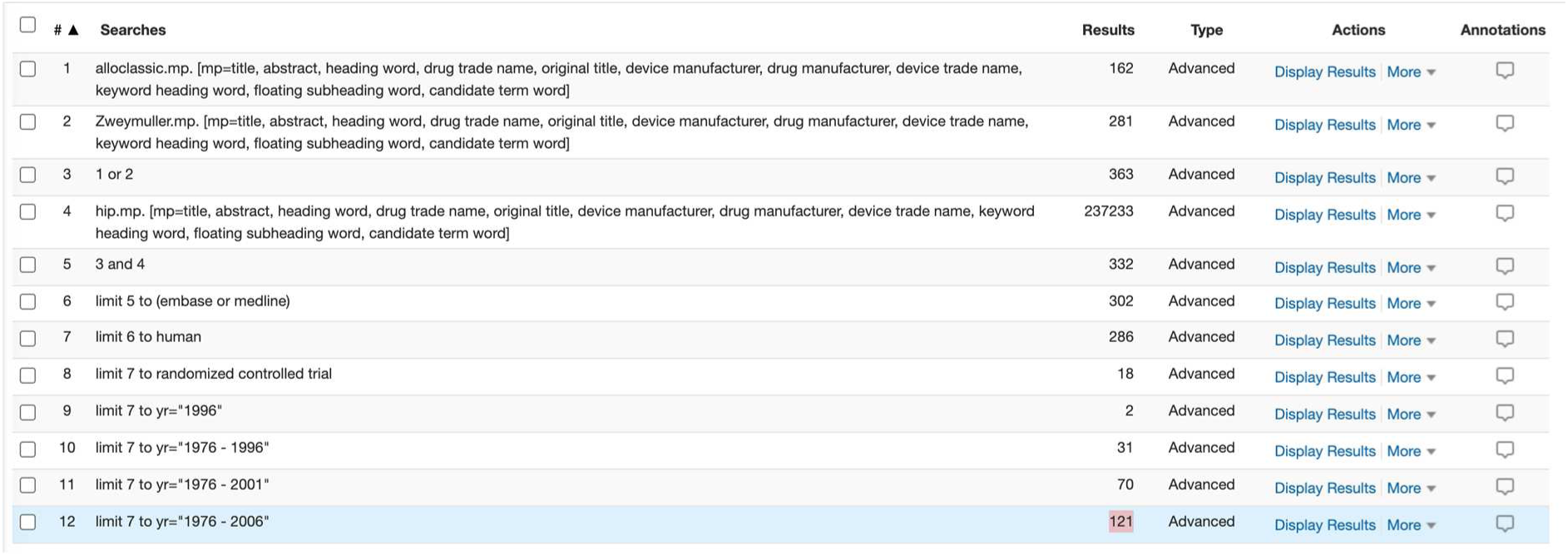

PubMed: 114

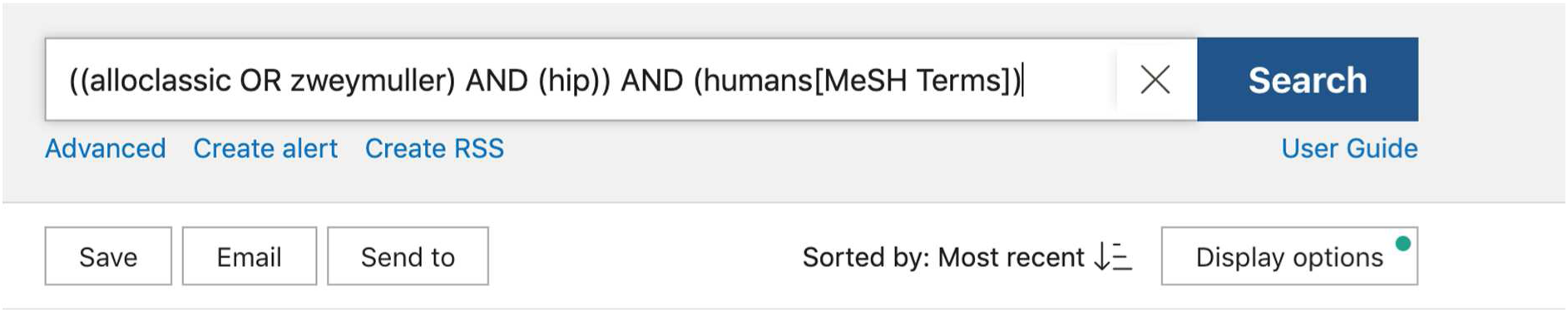

Web of Science: 97

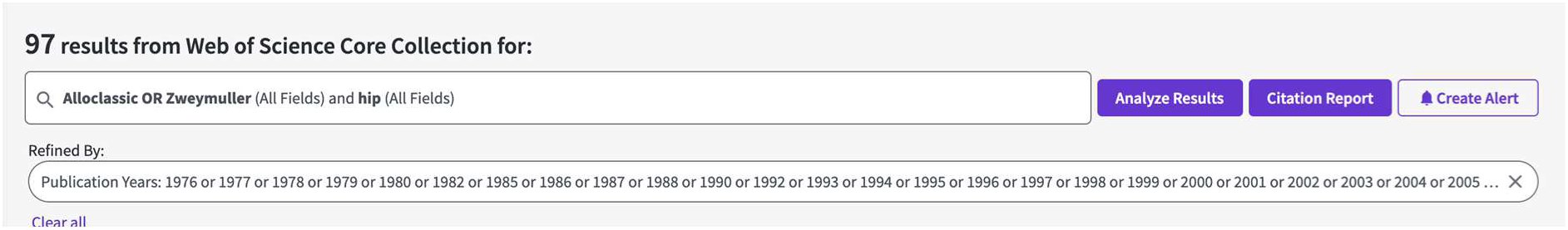

#### COLLO-MIS 27^th^ September 2021

Embase: 3

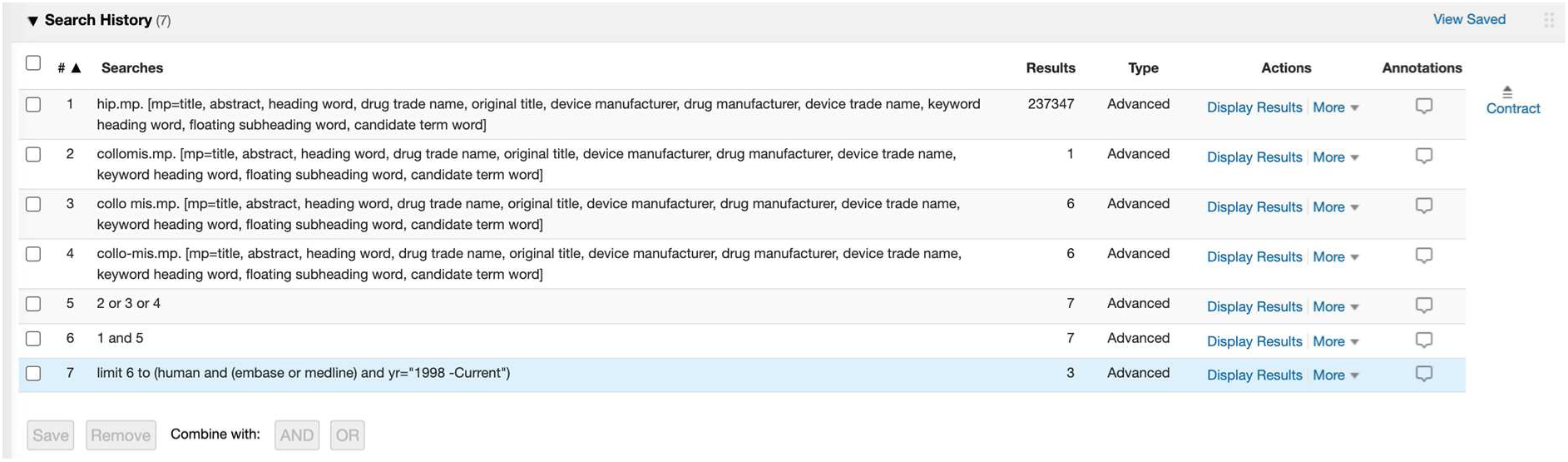

PubMed: 0

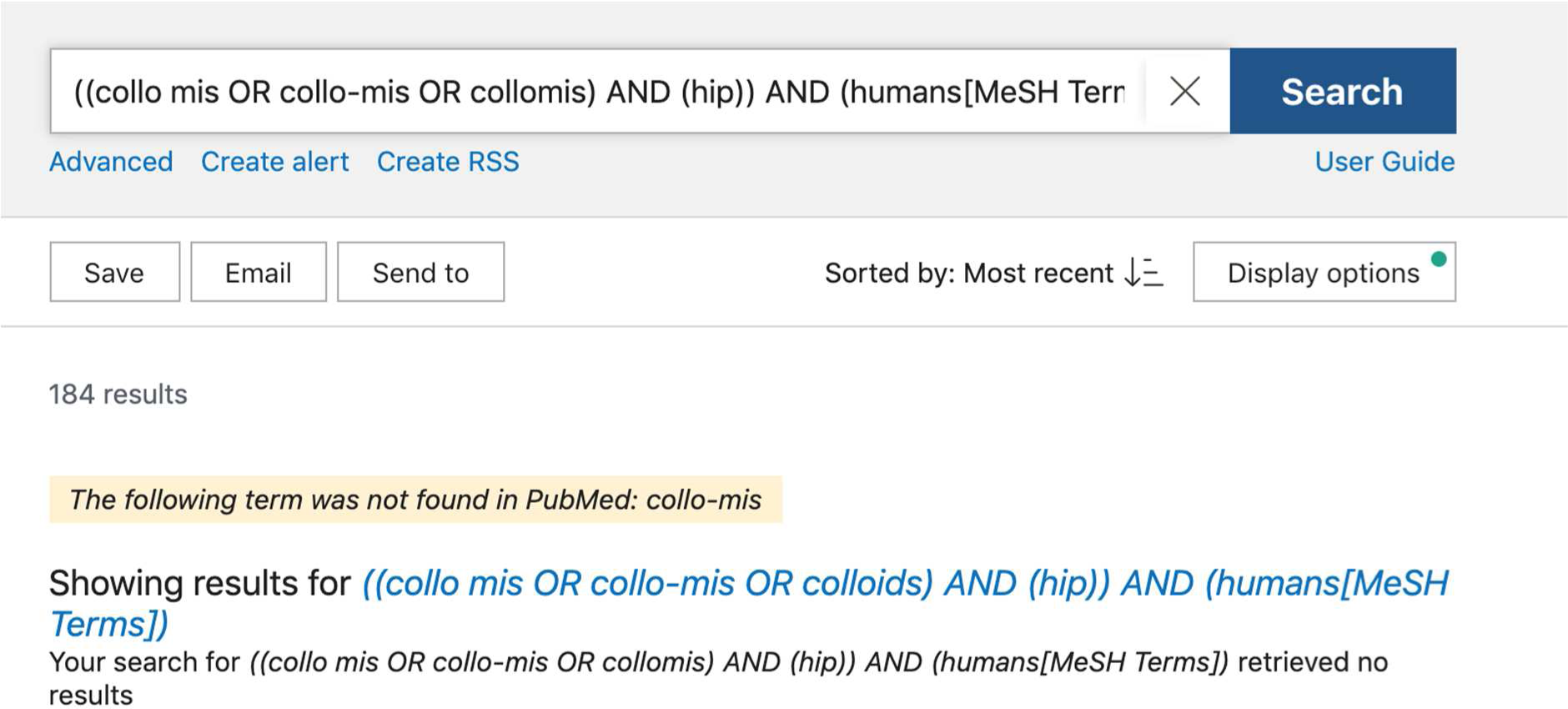

Web of Science: 0

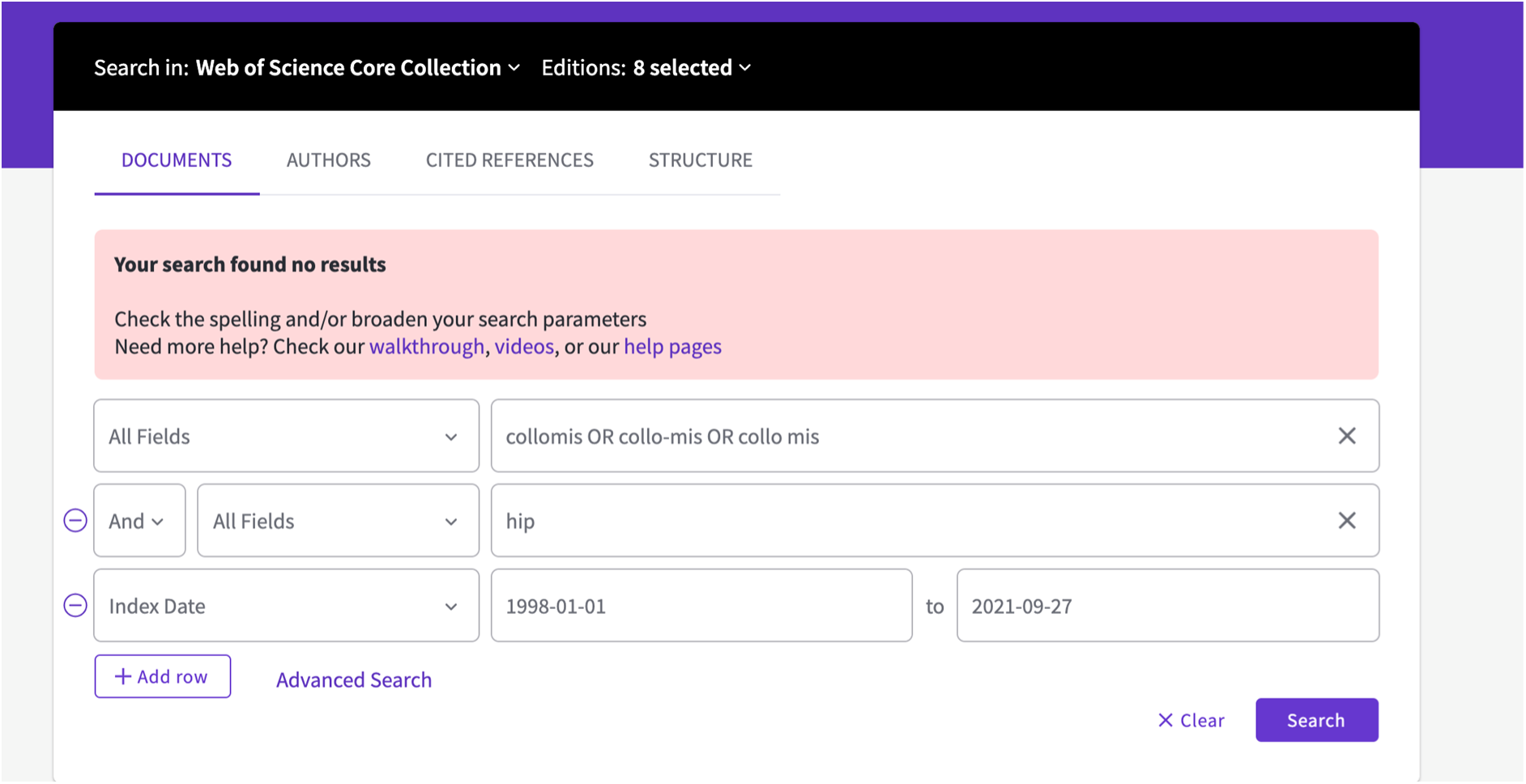

#### Accolade II: 29^th^ September 2021

Embase: 48

(#1 was hip.mp, #2 was accolade II.mp, missing in image below)

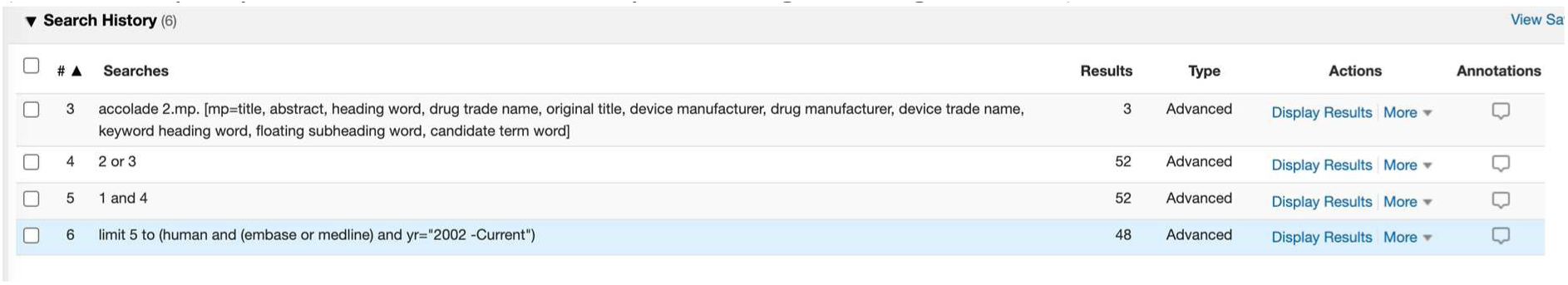

PubMed: 21

(((accolade II OR accolade 2) AND (hip)) AND (humans[MeSH Terms])) AND ((“2002/01/01”[Date - Publication]: “3000”[Date - Publication]))

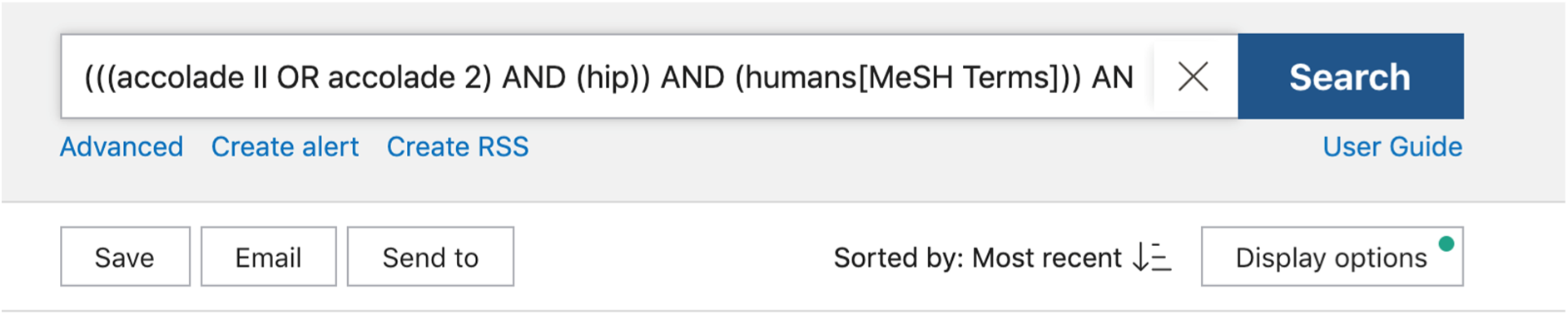

Web of Science: 25

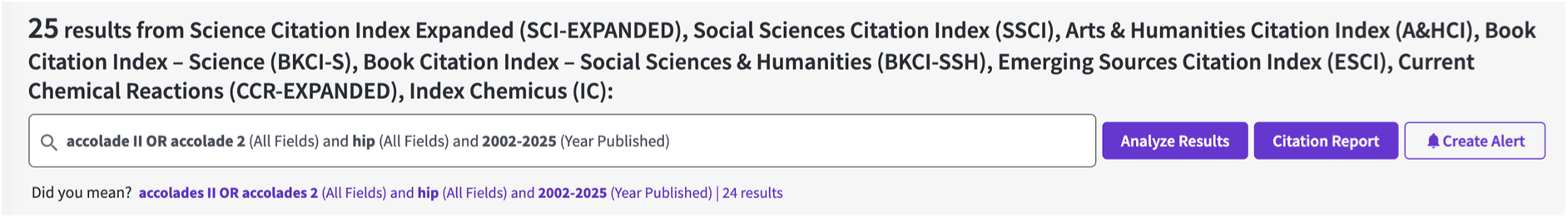

#### Bicontact: 5^th^ October 2021

Embase: 102

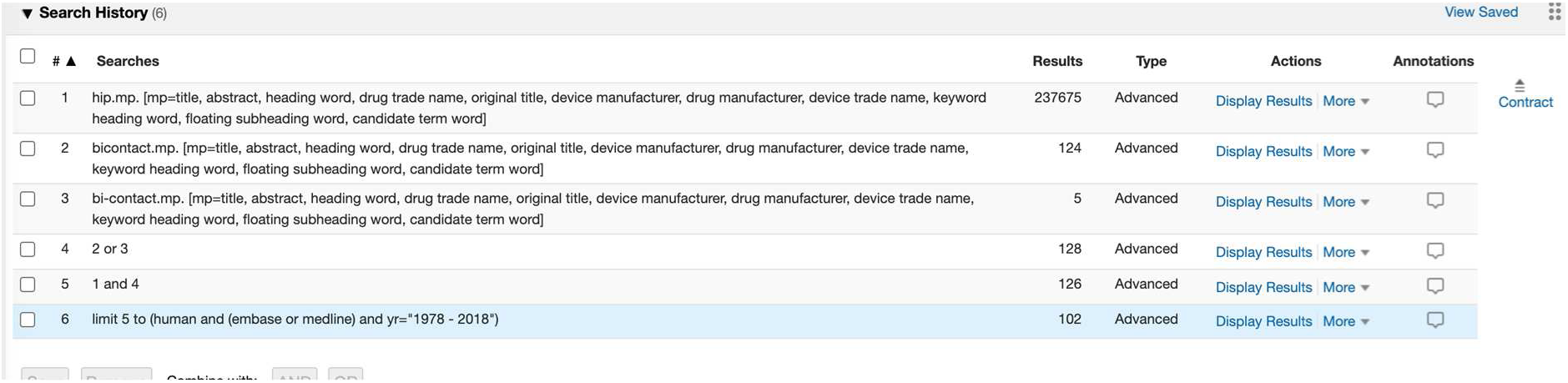

PubMed: 48

(((bi-contact OR bicontact) AND (hip)) AND (humans[MeSH Terms])) AND ((“1978”[Date - Publication]: “2018”[Date - Publication]))

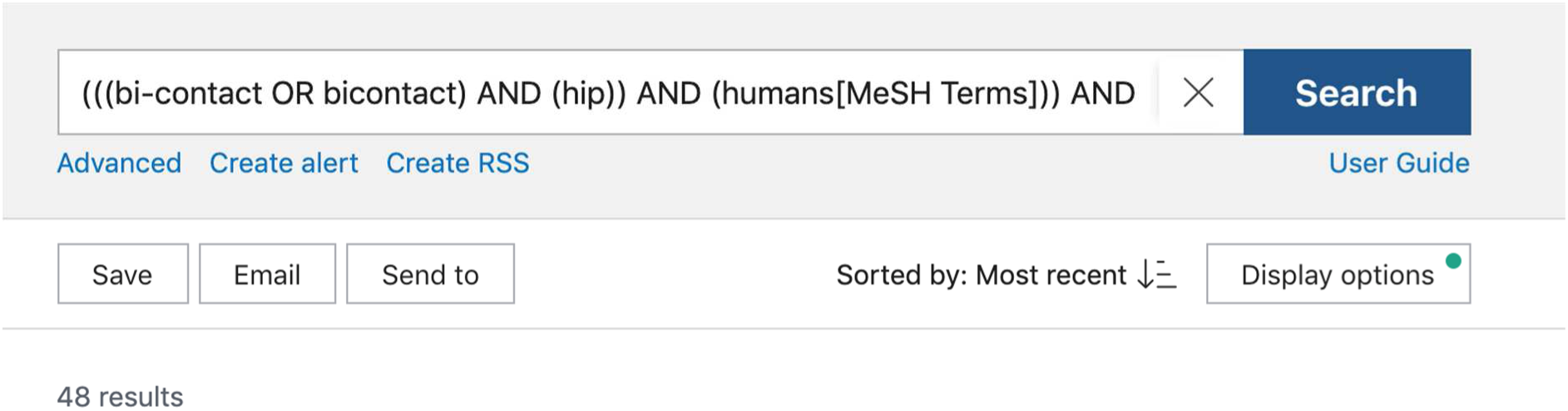

Web of Science: 43

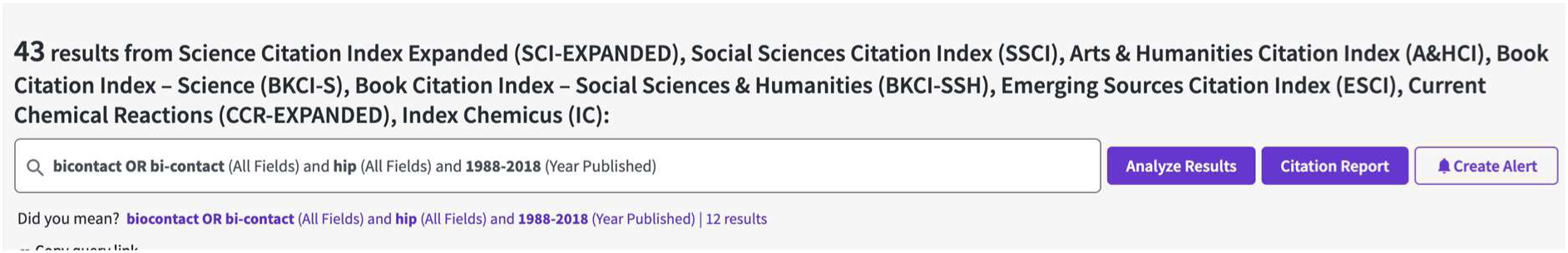

#### Minihip 7^th^ October 2021

Embase: 17

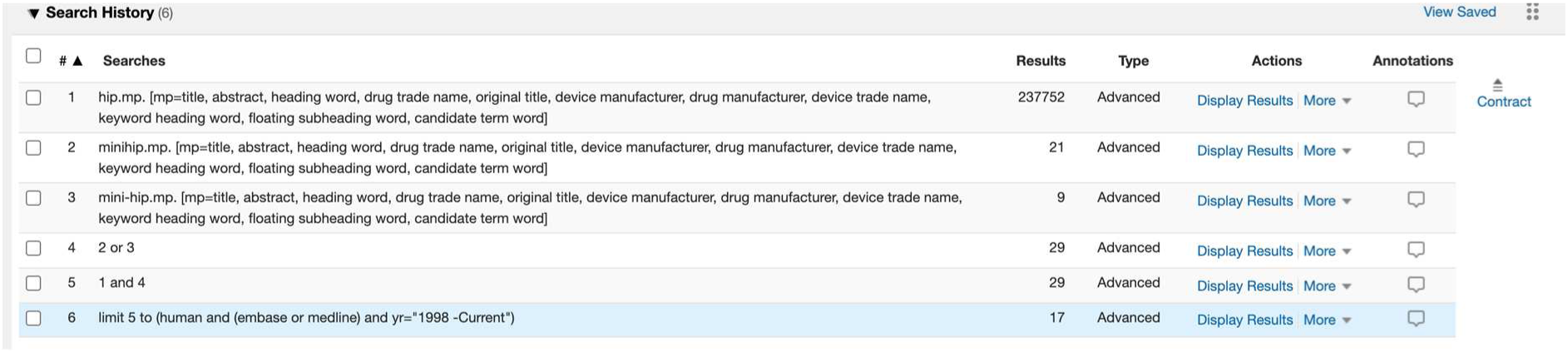

PubMed: 9

(((minihip OR mini-hip) AND (hip)) AND (humans[MeSH Terms])) AND ((“1998”[Date - Publication]: “3000”[Date - Publication]))

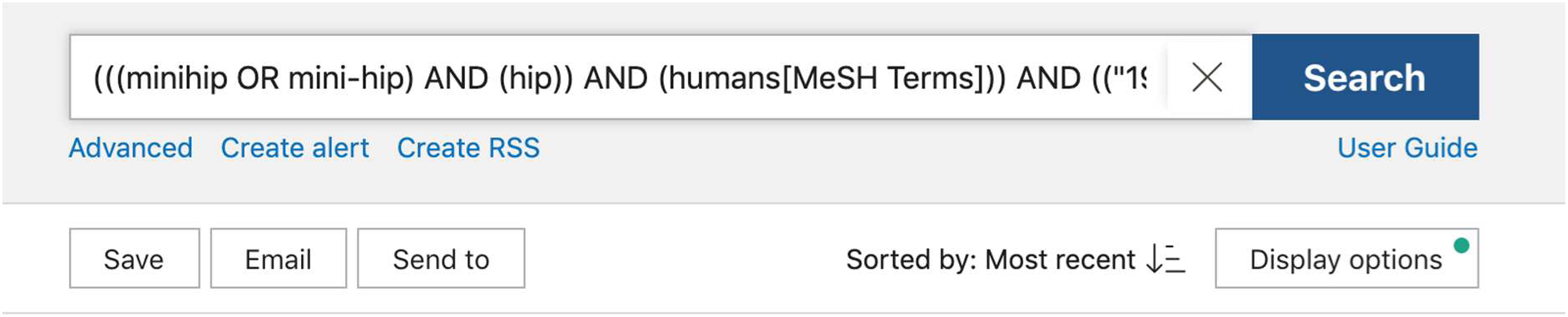

Web of Science: 19

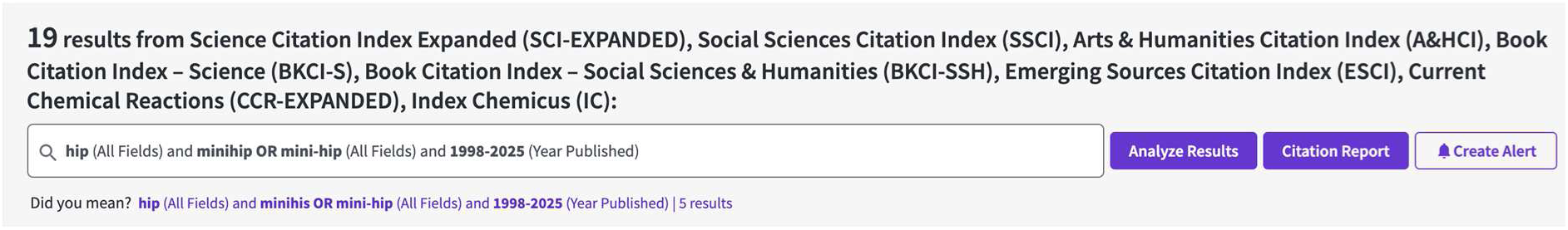

#### CSTEM 8^th^ October 2021

Embase 69

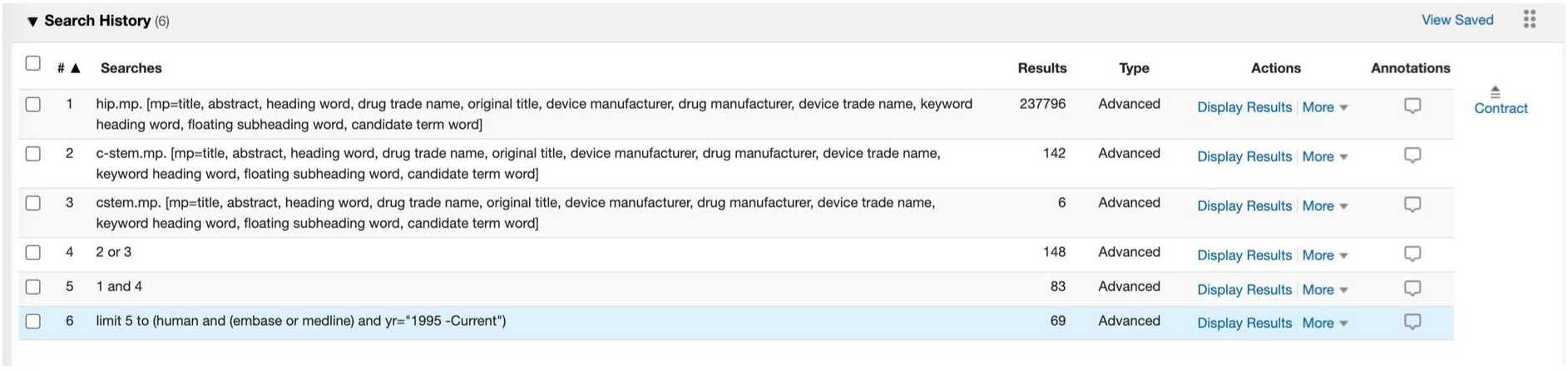

PubMed: 31

(((c-stem OR cstem) AND (hip)) AND (humans[MeSH Terms])) AND ((“1995”[Date - Publication]: “3000”[Date - Publication]))

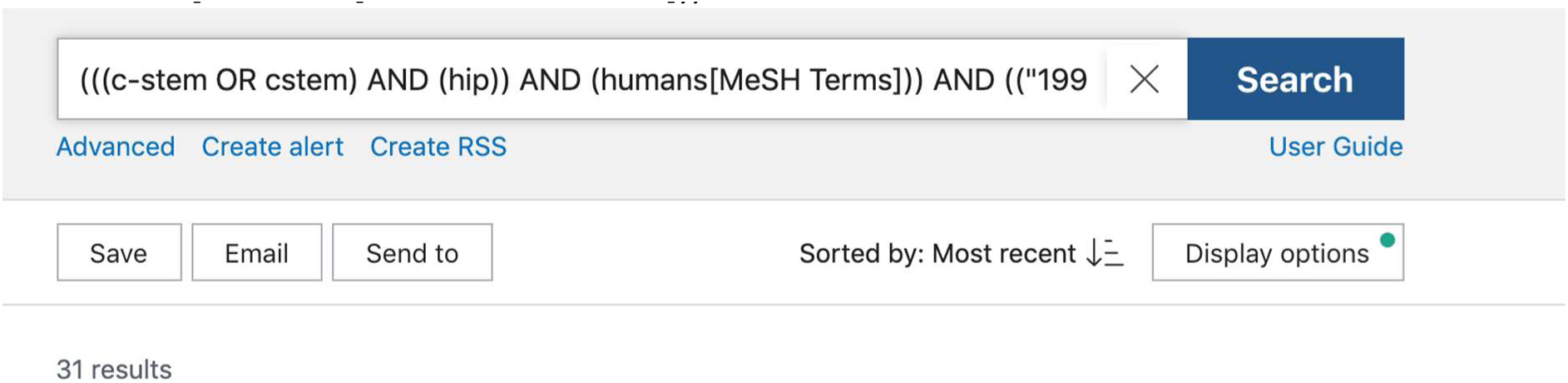

Web of Science: 34

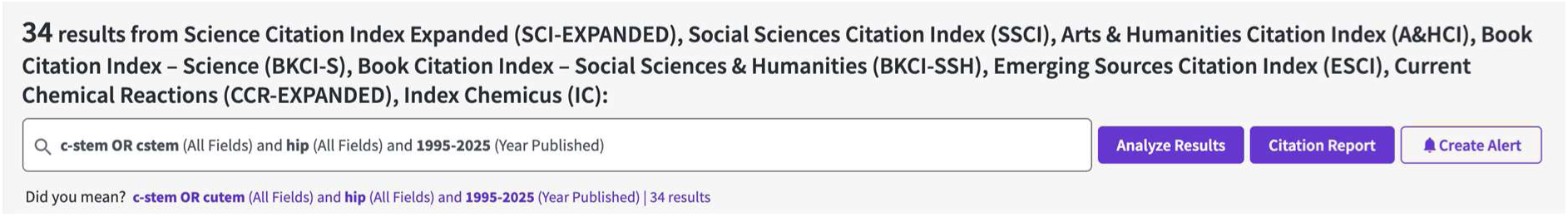

#### Filler 3ND 15^th^ November 2021

Searches were not restricted by date or to humans as no results were returned)

Embase 0

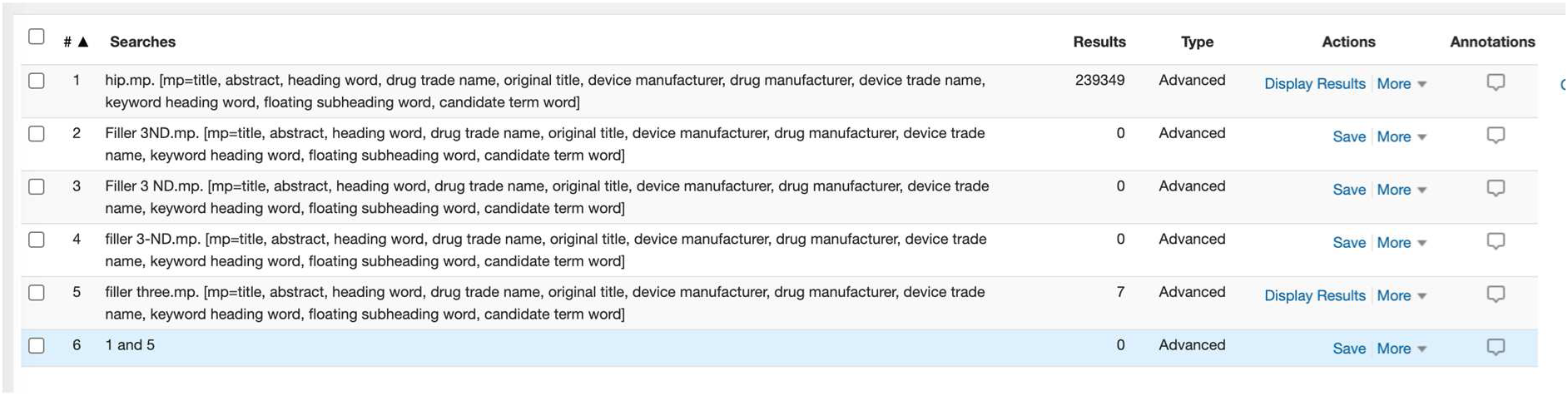

PubMed: 0

It returns 8 but none of them include the phrases.

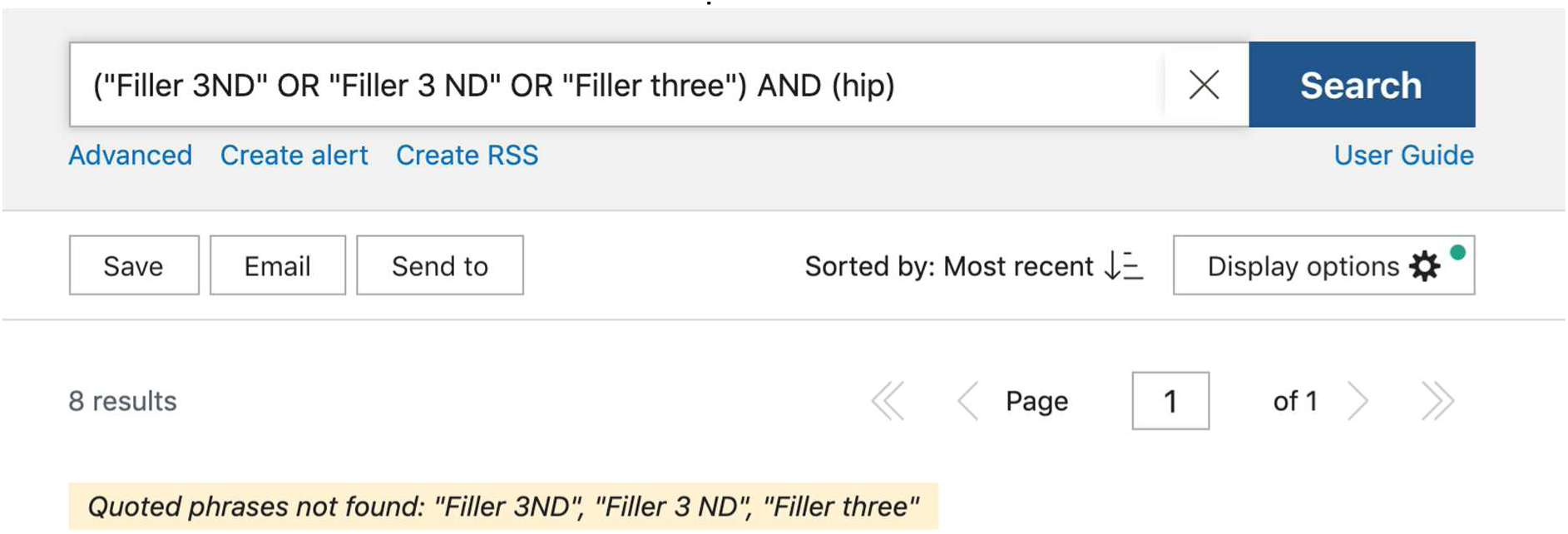

Web of Science: 0

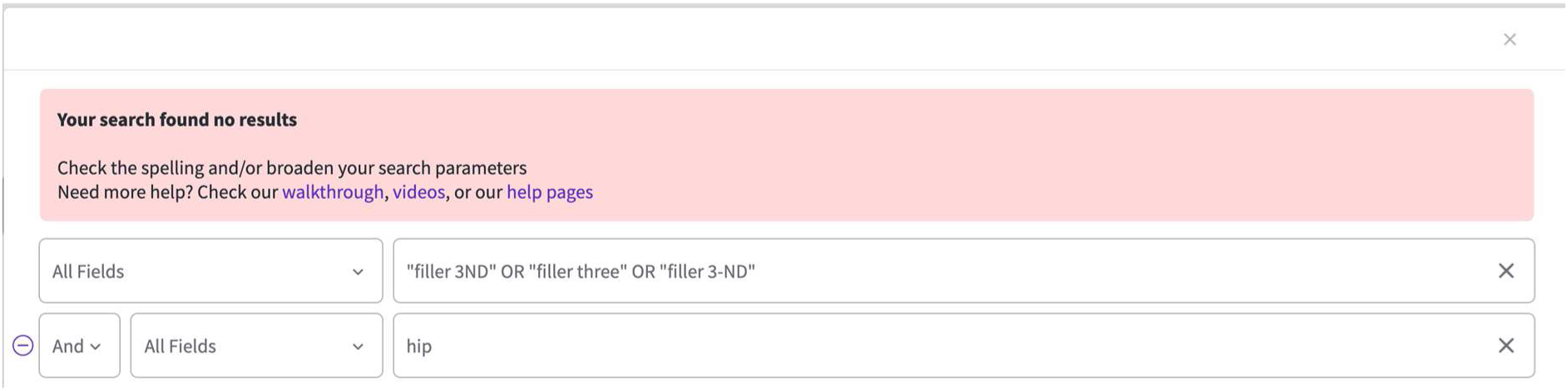

#### Stelia 15^th^ November 2021

Embase: 0

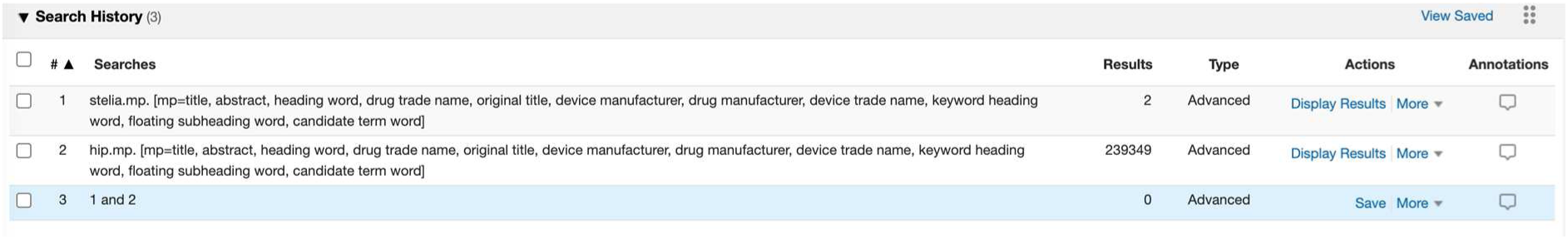

PubMed: 0

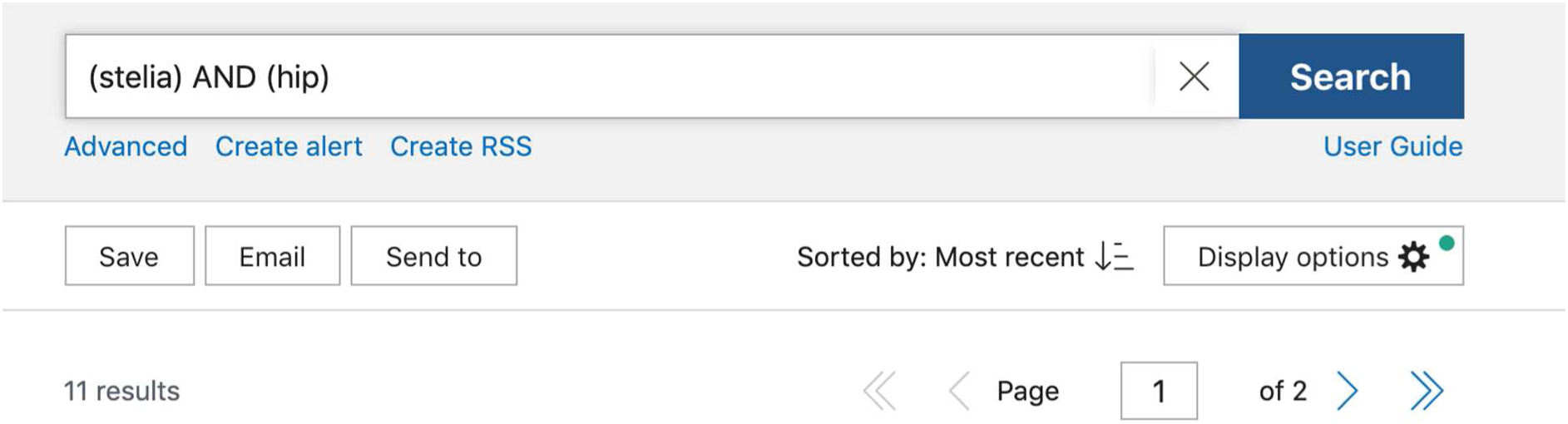

Web of Science: 0

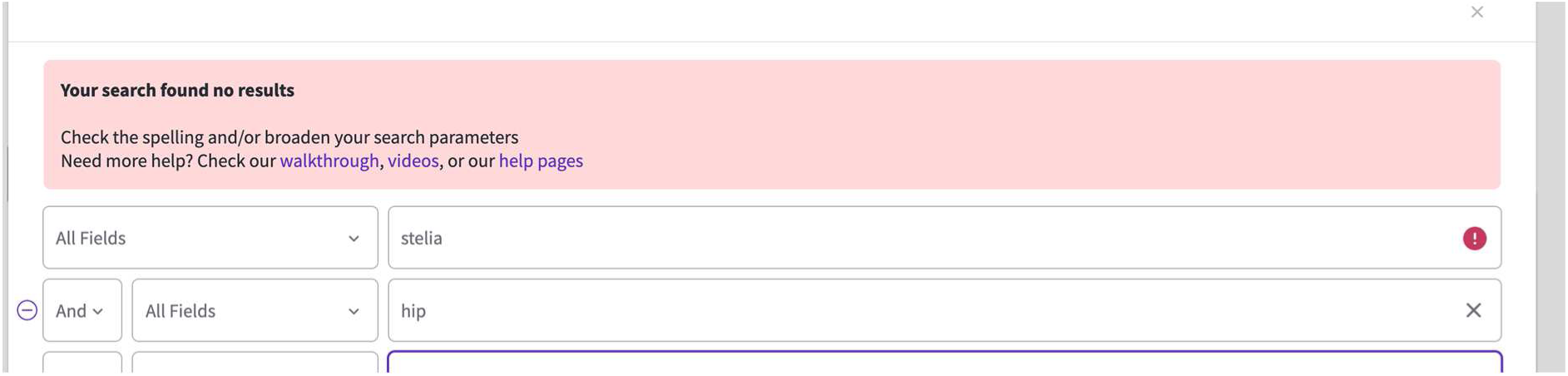

### Cup searches

#### Versafit 30^th^ December 2021

Embase: 29

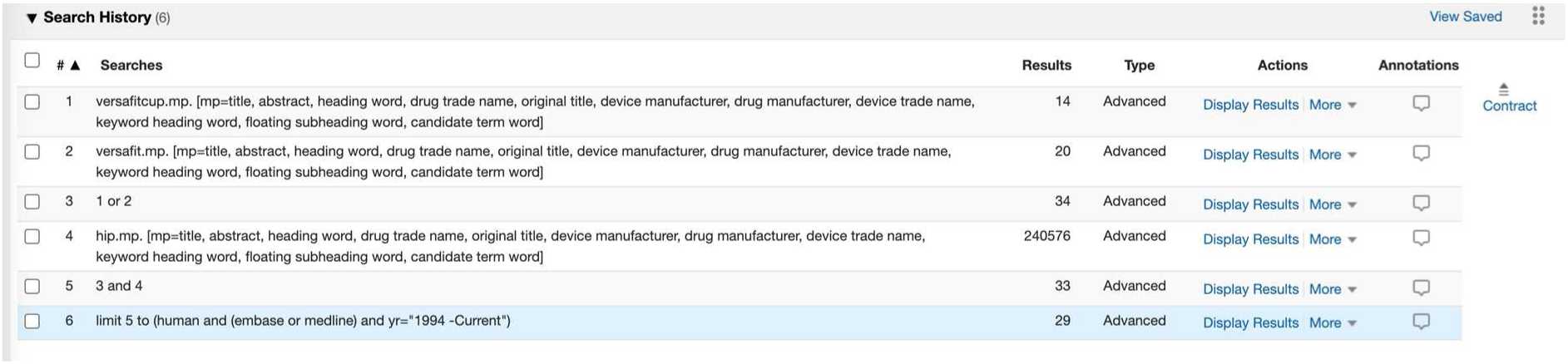

PubMed: 2

(((versafit OR versafitcup) AND (hip)) AND (humans[MeSH Terms])) AND ((“1994”[Date - Publication]: “3000”[Date - Publication]))

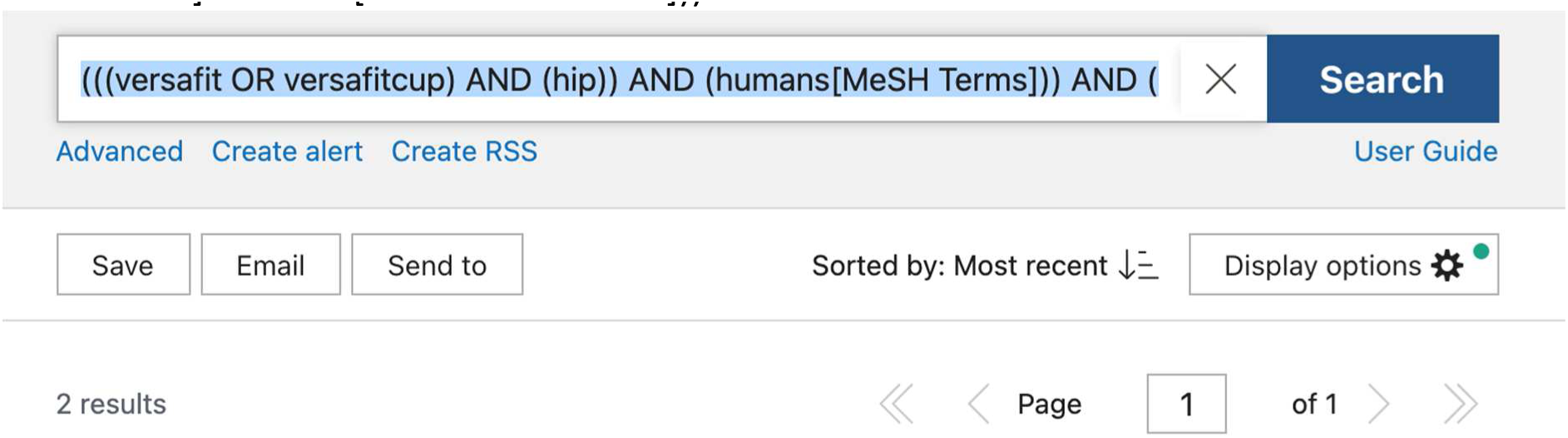

Web of Science: 2

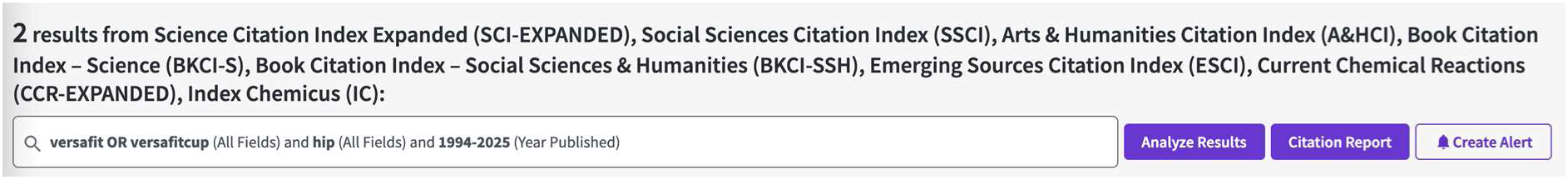

#### Polar cup cemented 30^th^ December 2021

Embase 14

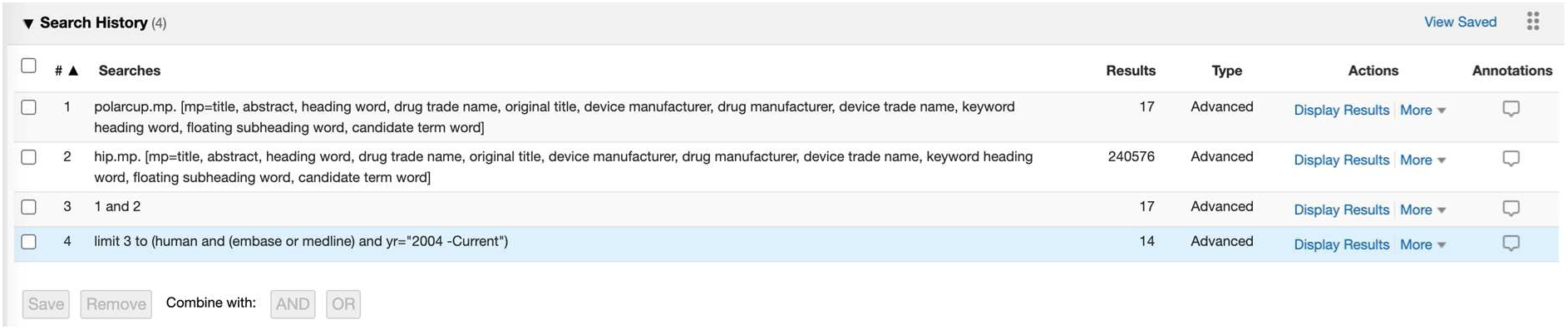

PubMed: 2

(((polarcup) AND (hip)) AND (humans[MeSH Terms])) AND ((“2004”[Date - Publication]: “3000”[Date - Publication]))

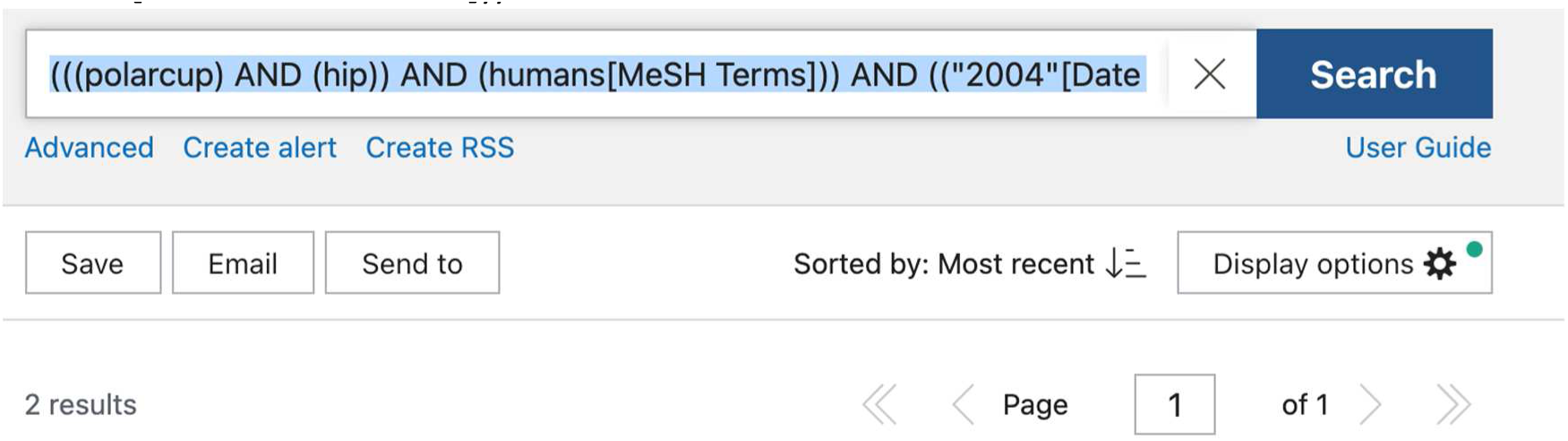

Web of Science: 2

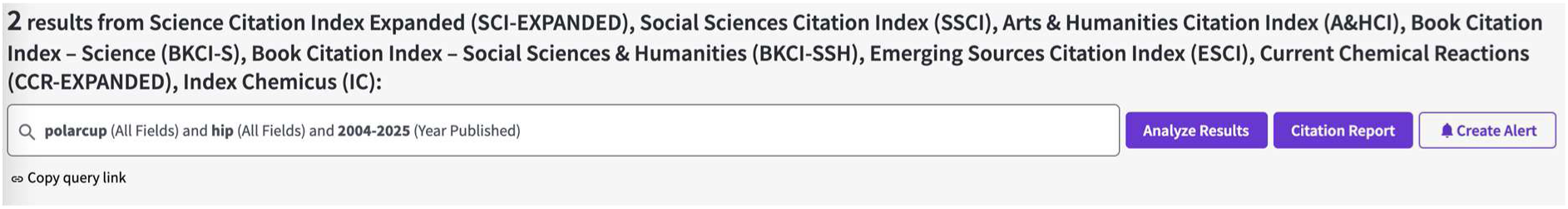

#### Plasmacup 30^th^ December 2021

Embase: 62

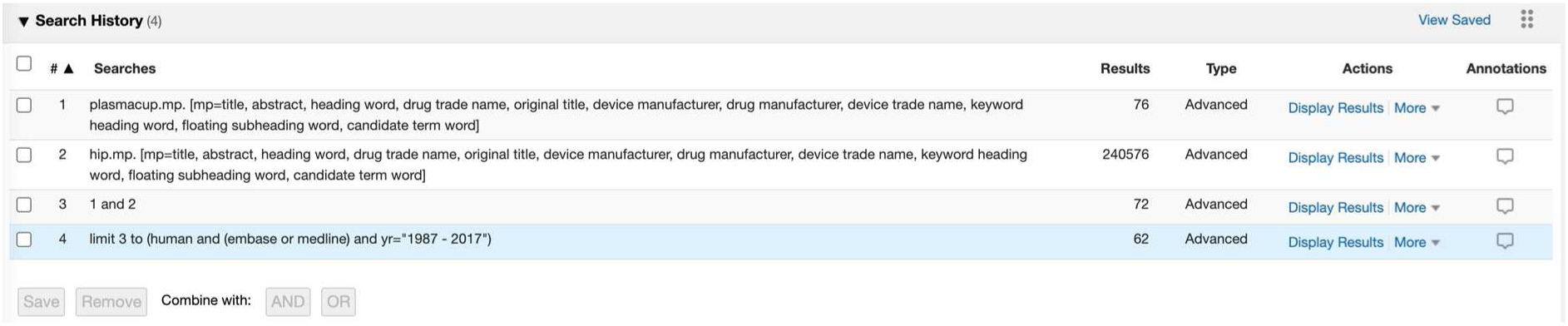

PubMed: 13

(((plasmacup) AND (hip)) AND (humans[MeSH Terms])) AND ((“1987”[Date - Publication]: “2017”[Date - Publication]))

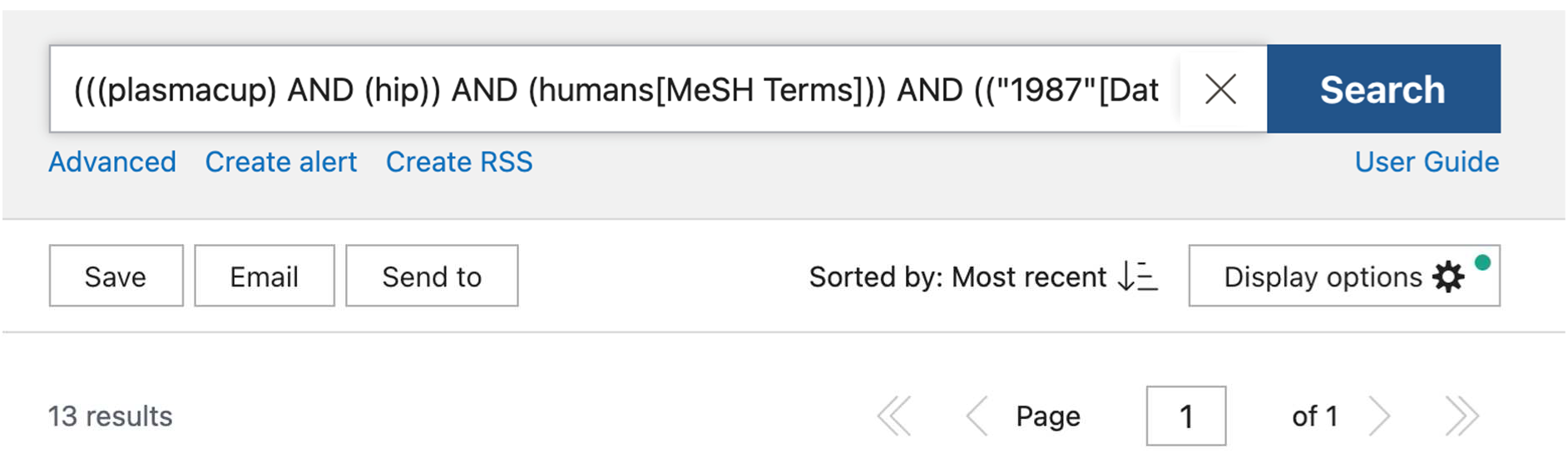

Web of Science: 12

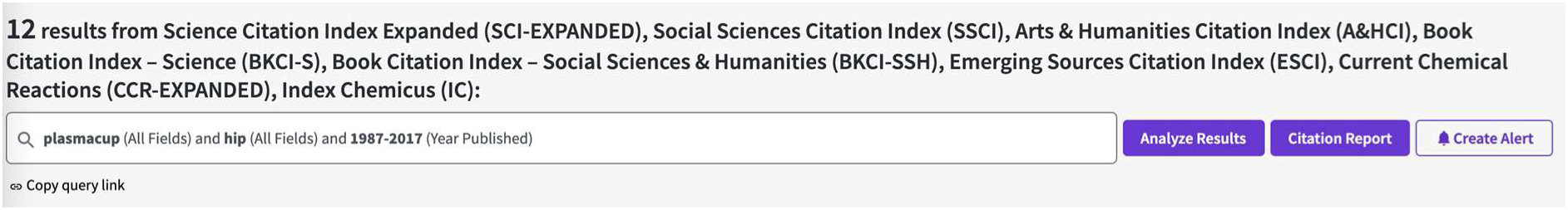

#### Exceed ABT 30^th^ December 2021

Embase 35

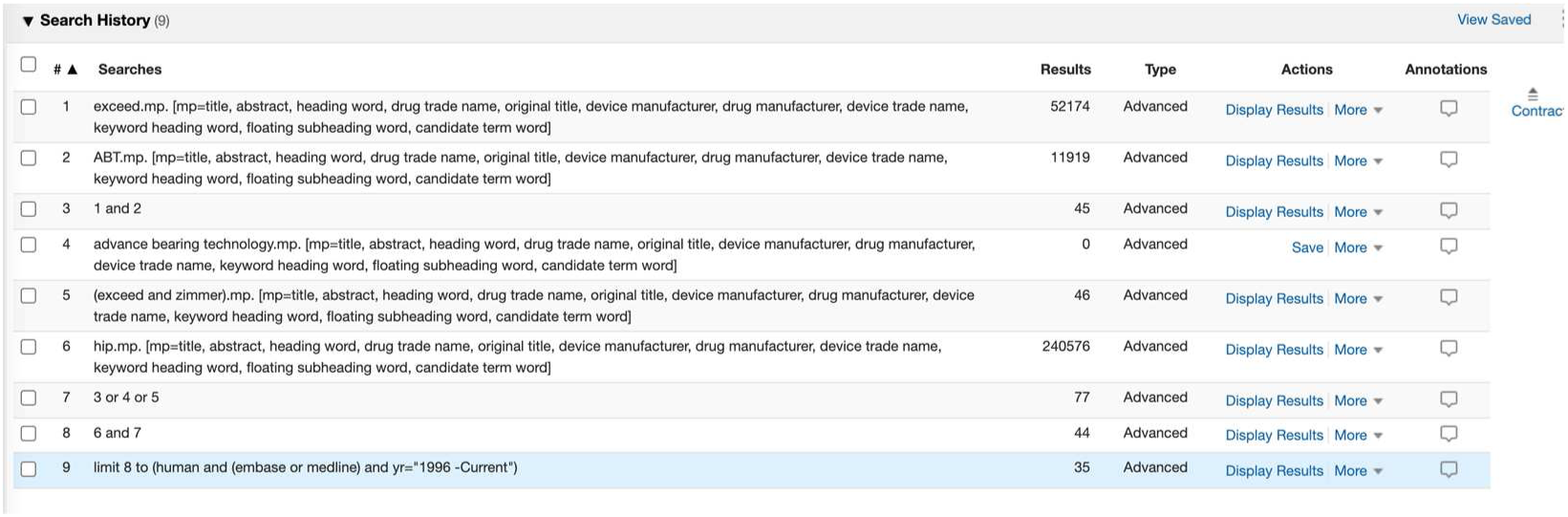

PubMed: 8

“advance bearing technology” does not match anything so pubmed changes the search to not quote it. It was therefore not searched.

(((exceed AND ABT) OR (exceed AND zimmer)) AND (hip)) AND ((“1996”[Date - Publication]: “2025”[Date - Publication]))

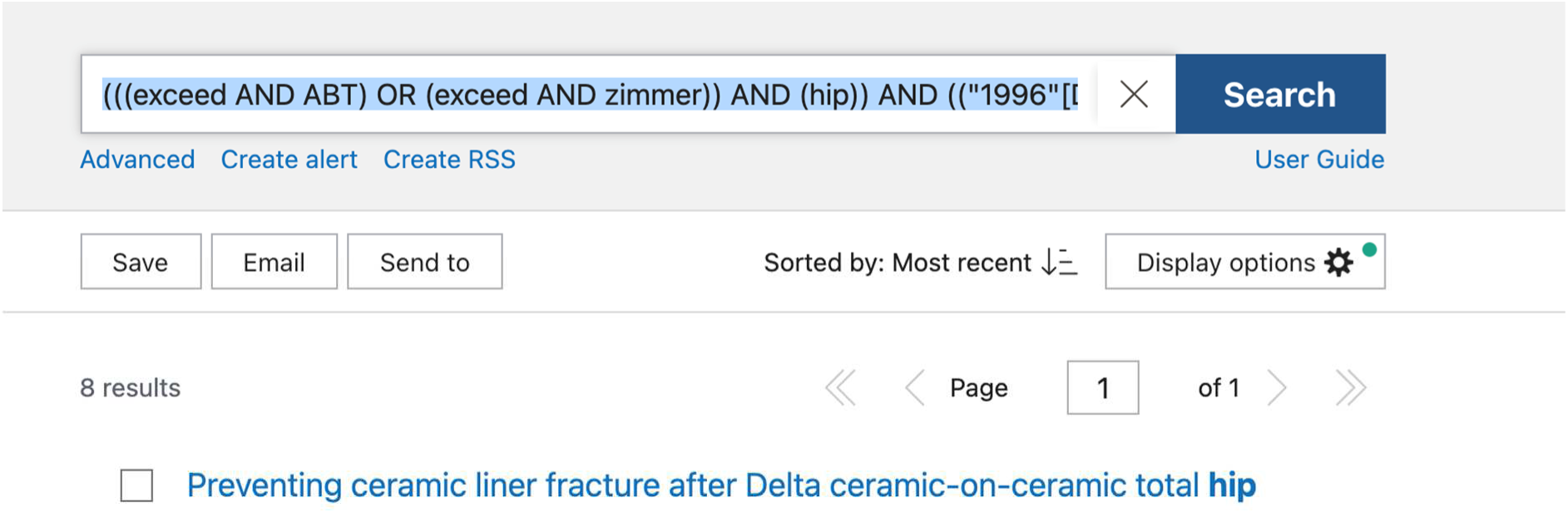

Web of Science: 31

“advance bearing technology” also gives no results

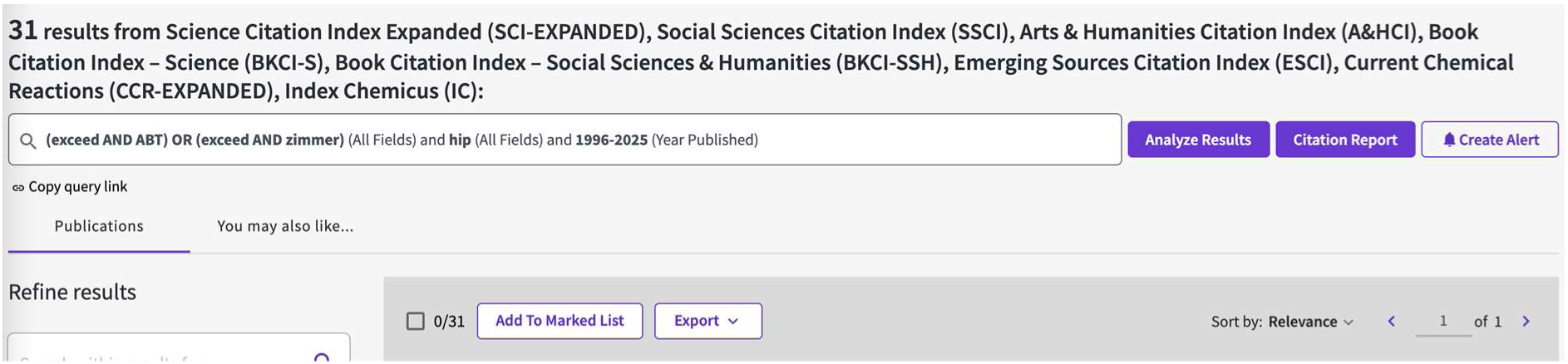

#### Anexys 30^th^ December 2021

Embase: 1

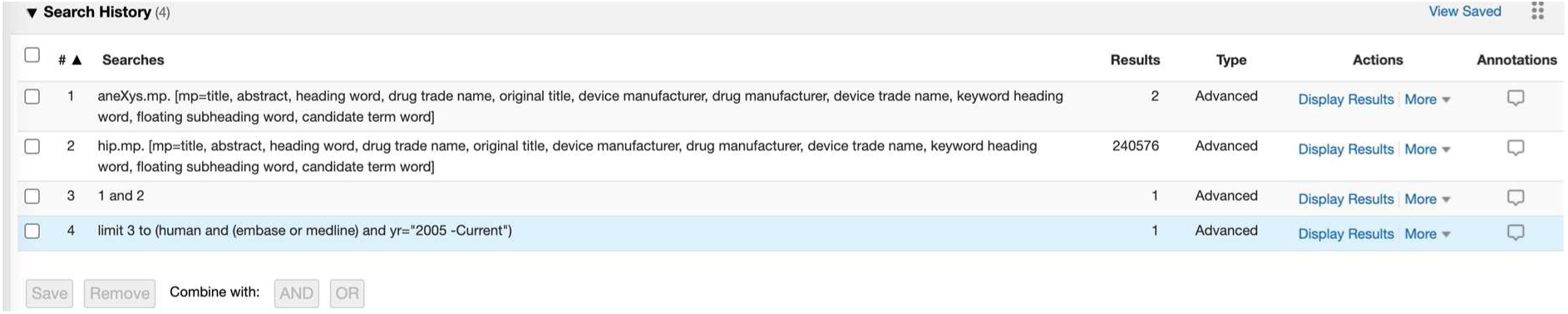

PubMed: 1

(((anexys) AND (hip)) AND (humans[MeSH Terms])) AND ((“2005”[Date - Publication]: “3000”[Date - Publication]))

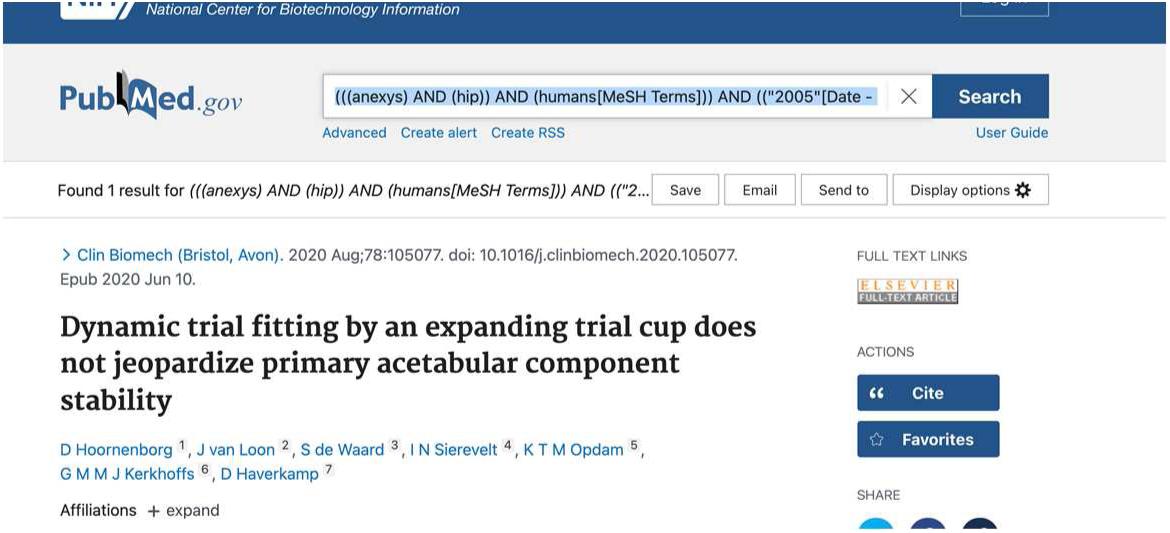

Web of Science: 2

One of them is the one from pubmed and embase

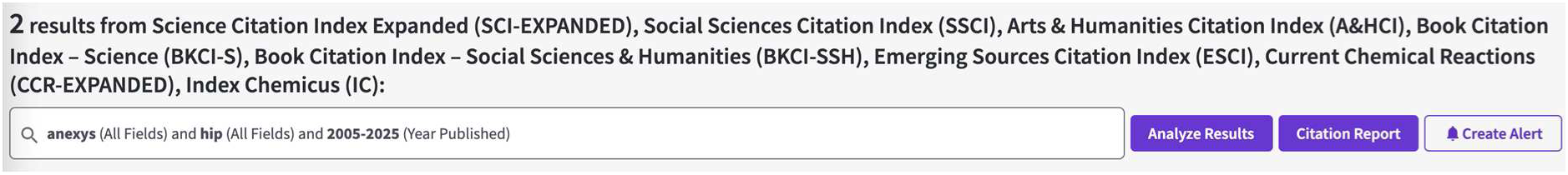

#### Cenator 30^th^ December 2021

Embase: 2

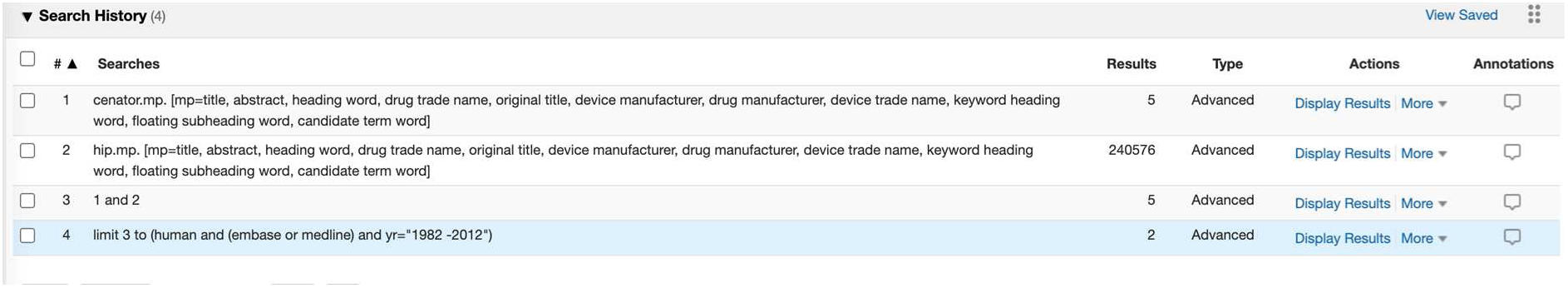

PubMed: 0

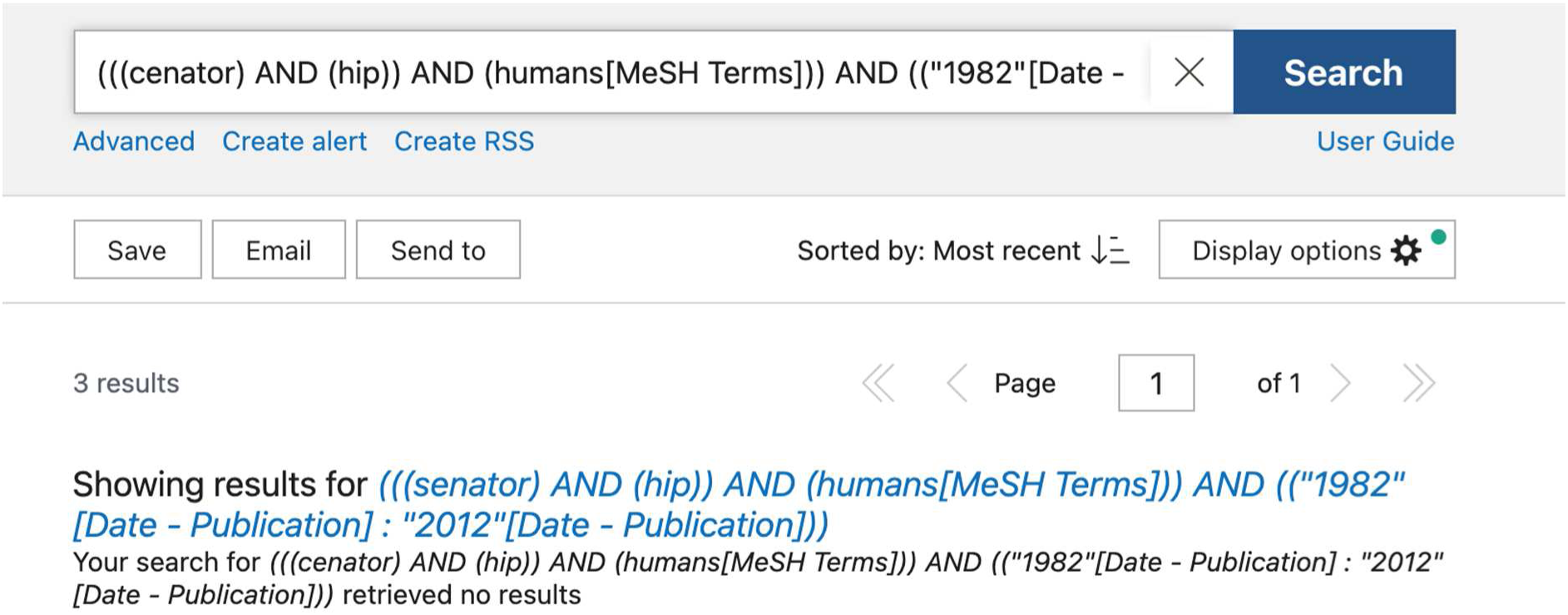

Web of Science: 0

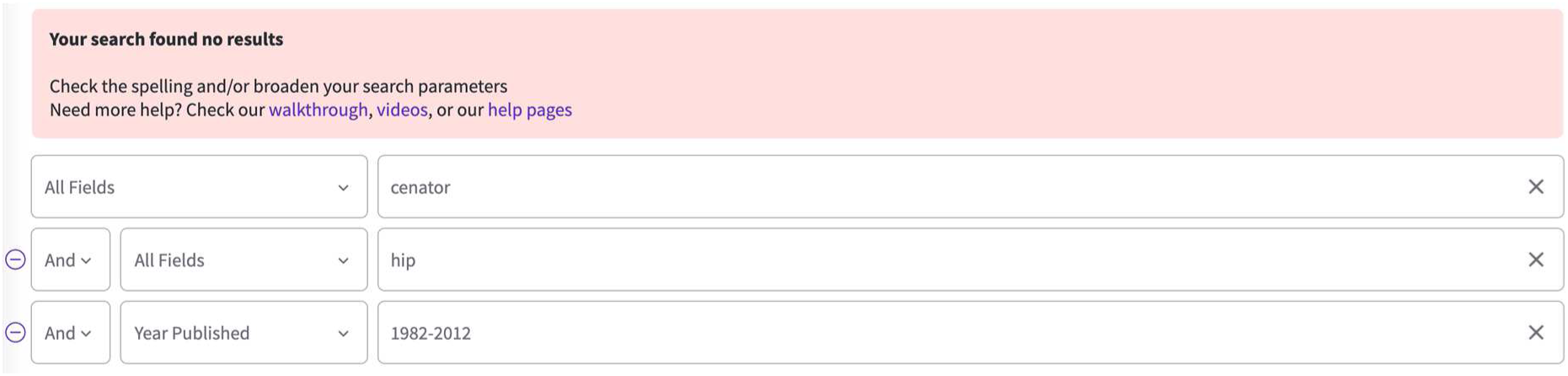

#### Ecofit 30^th^ December 2021

Embase: 13

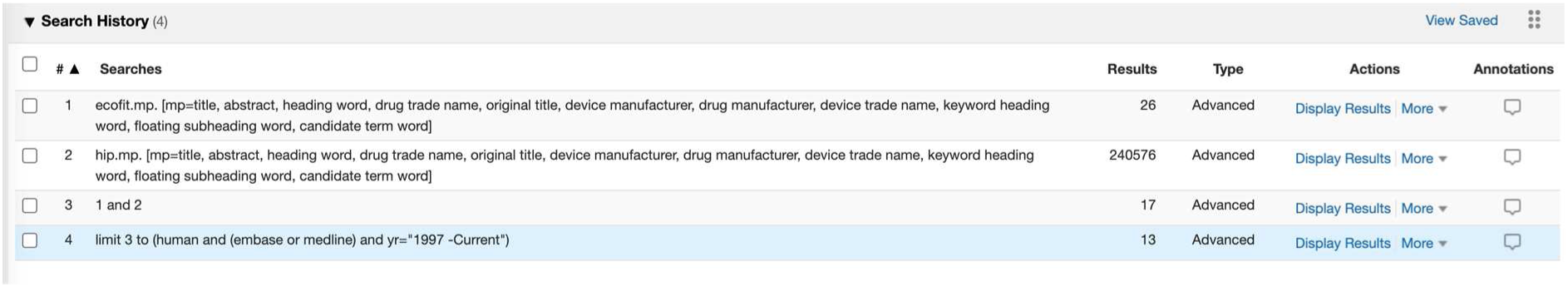

PubMed: 4

(((ecofit) AND (hip)) AND (humans[MeSH Terms])) AND ((“1997”[Date - Publication]: “3000”[Date - Publication]))

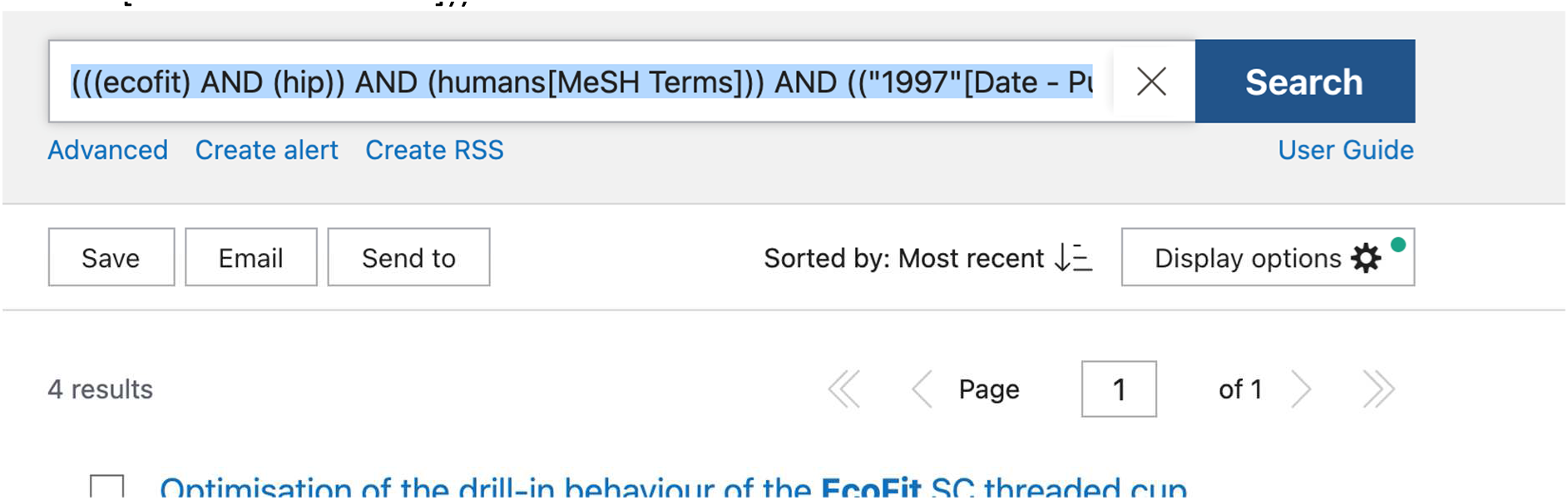

Web of Science: 3

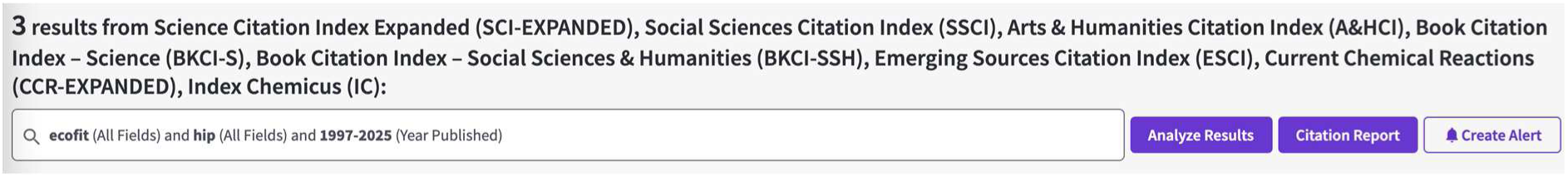

#### Link IP 4^th^ Jan 2022

Embase: 14

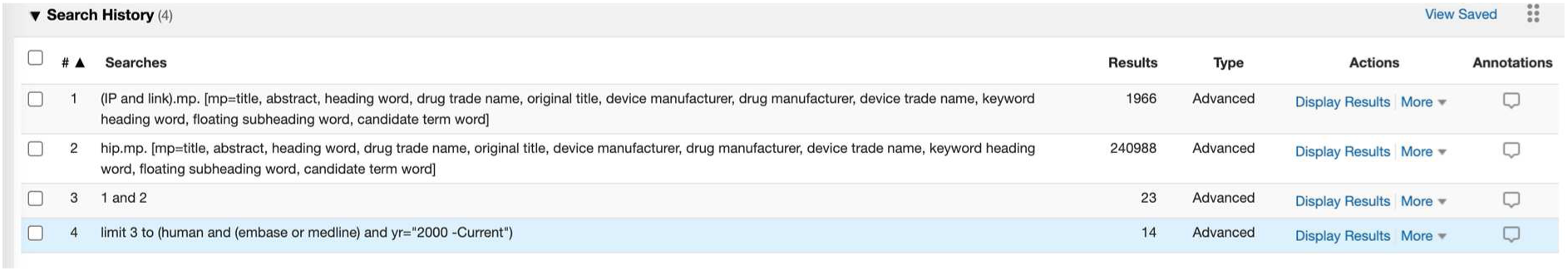

PubMed: 3

(((Link AND IP) AND (hip)) AND (humans[MeSH Terms])) AND ((“2000”[Date - Publication]: “3000”[Date - Publication]))

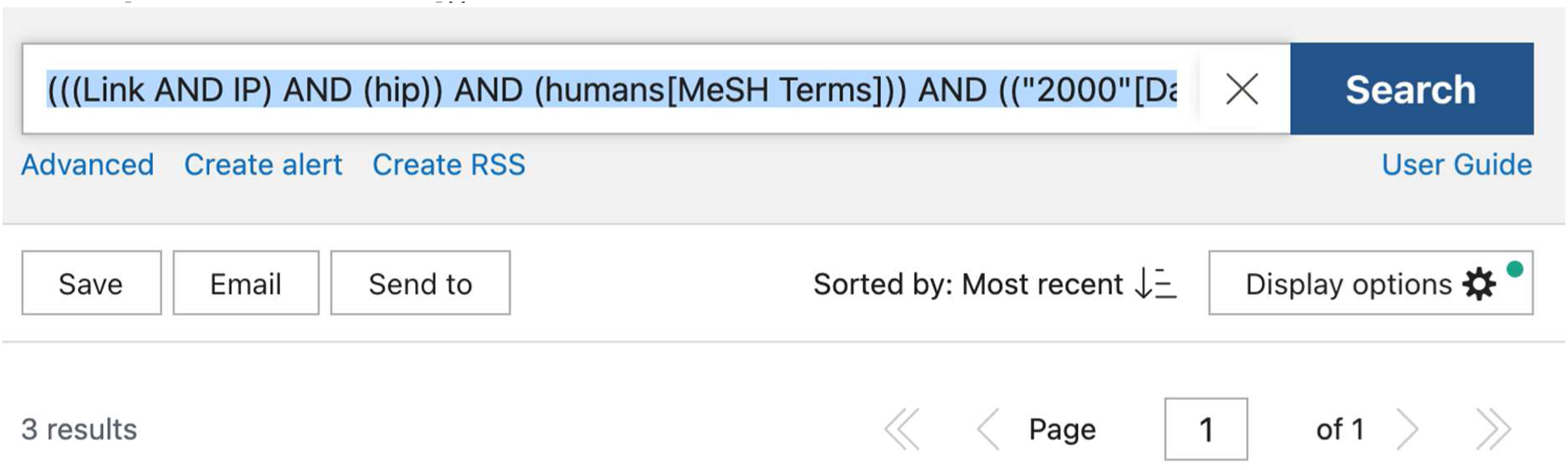

Web of Science: 37

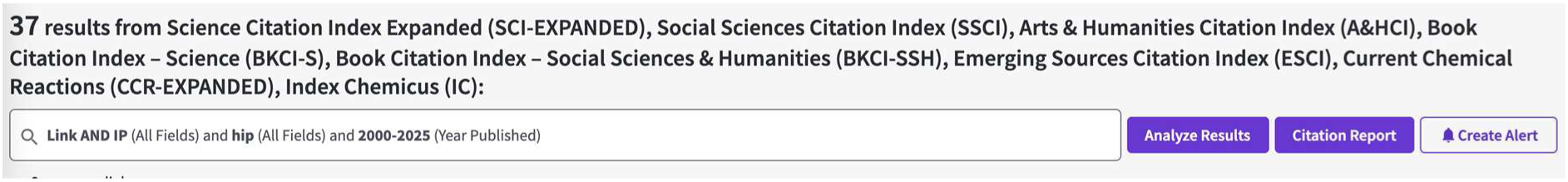

#### ANA.NOVA 4^th^ Jan 2022

Embase: 5

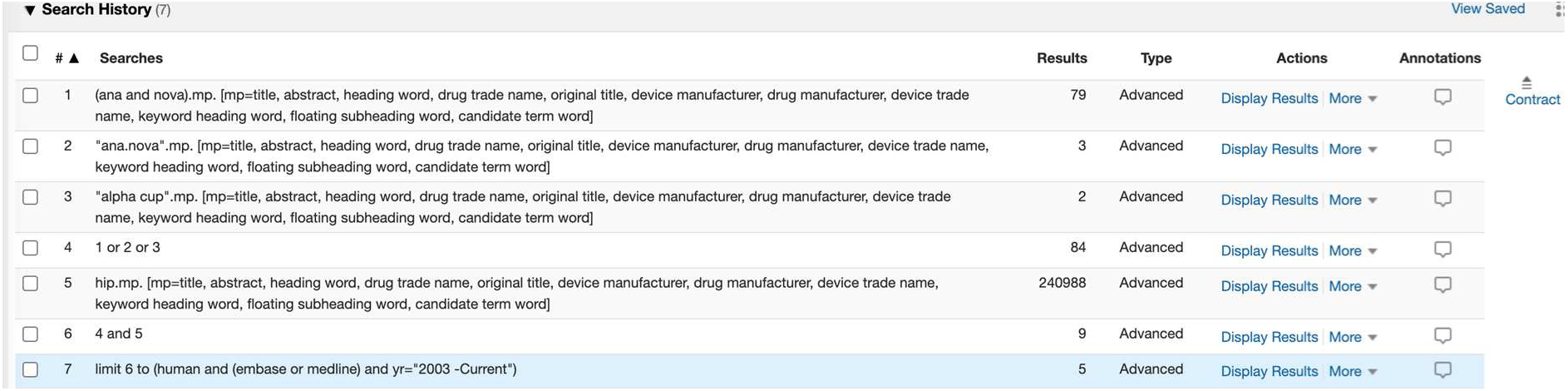

PubMed: 2

“alpha cup” not found

“ana.nova” not found

((((ana AND nova) OR (ana nova)) AND (hip)) AND (humans[MeSH Terms])) AND ((“2003”[Date - Publication]: “3000”[Date - Publication]))

Web of Science: 29

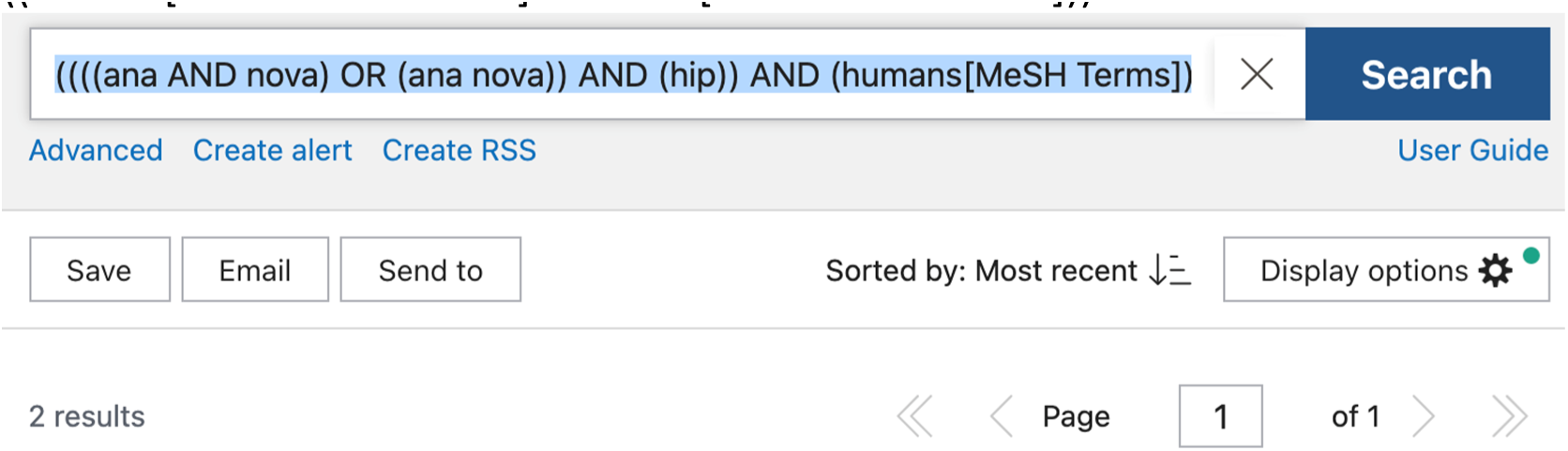

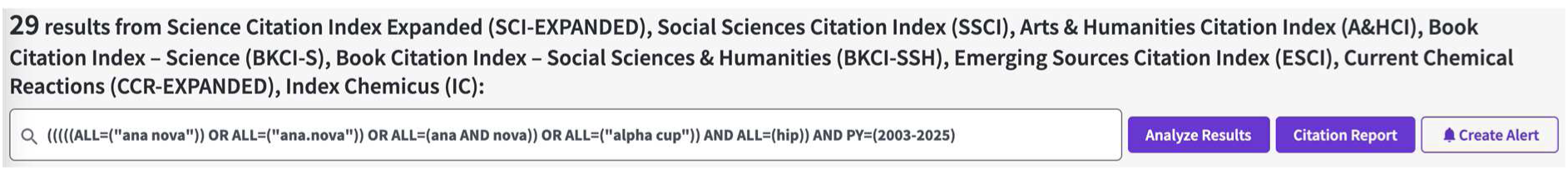

#### RM pressfit vitamys 22^nd^ Jan 2022

(Mathys)

Just searched RM pressfit as vitamys is a particular type

CE-marking date from Olga in “Re: Update and another question” of 2009

Embase: 24

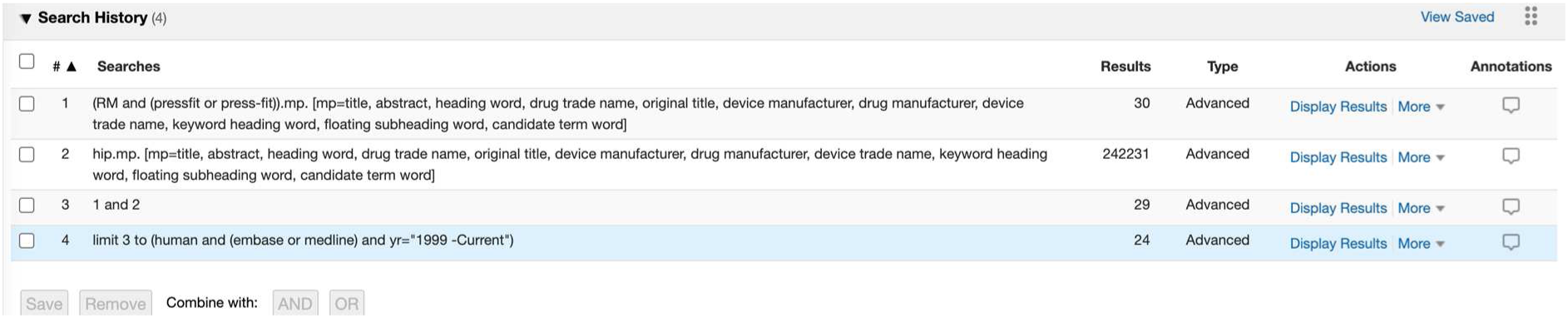

PubMed: 15

((RM AND (pressfit OR press-fit)) AND (humans[MeSH Terms])) AND ((“1999”[Date - Publication]: “3000”[Date - Publication]))

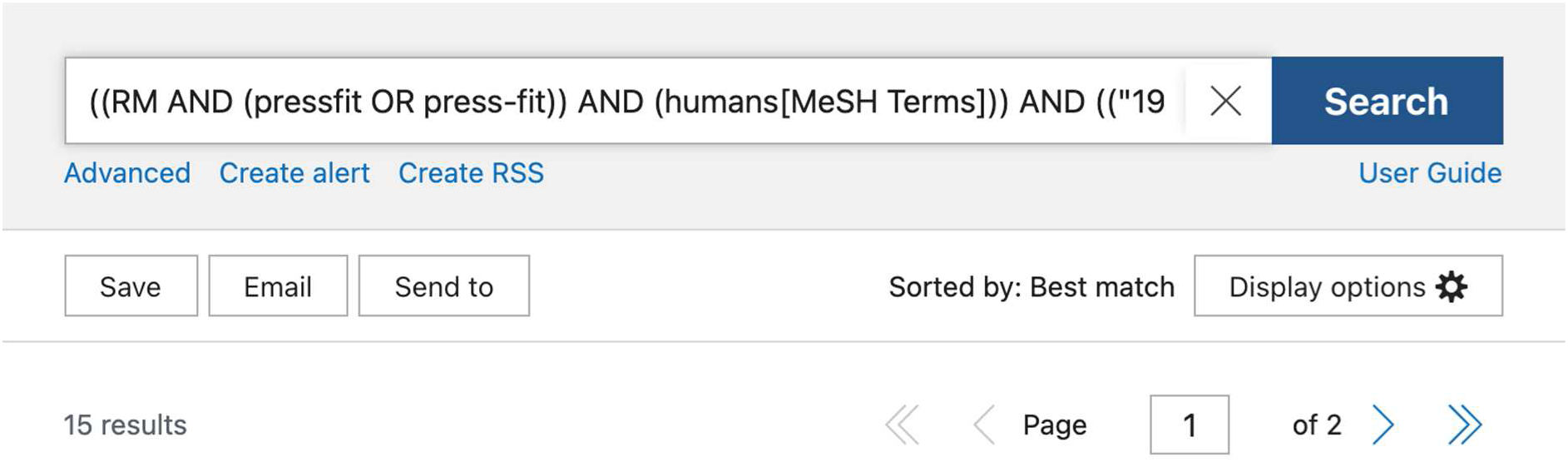

Web of Science: 19

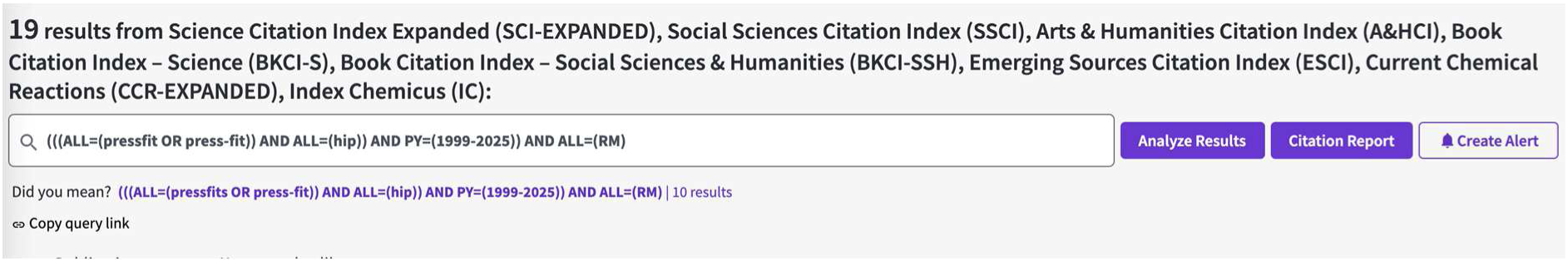

### Knee searches

#### Balansys CR 12^th^ October 2021

Embase: 15

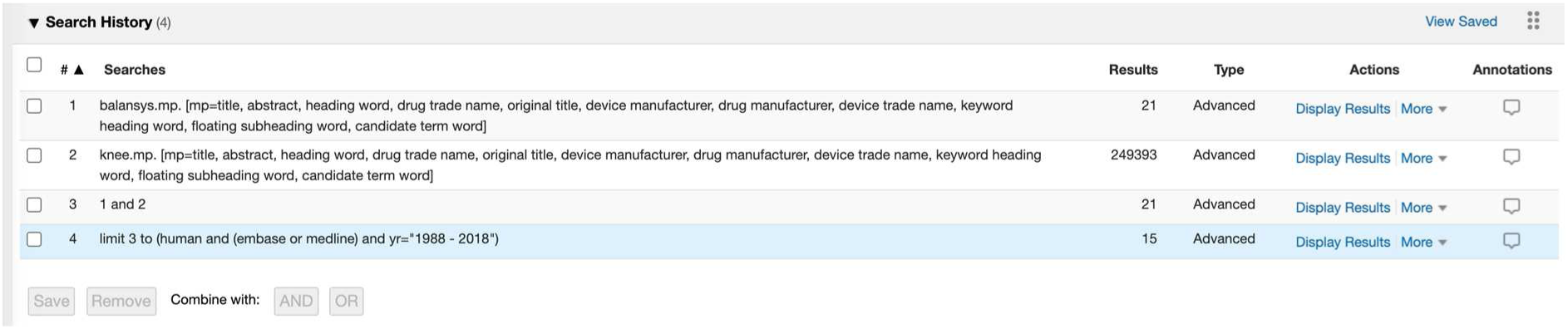

PubMed: 7

(((balansys) AND (knee)) AND (humans[MeSH Terms])) AND ((“1988”[Date - Publication]: “3000”[Date - Publication]))

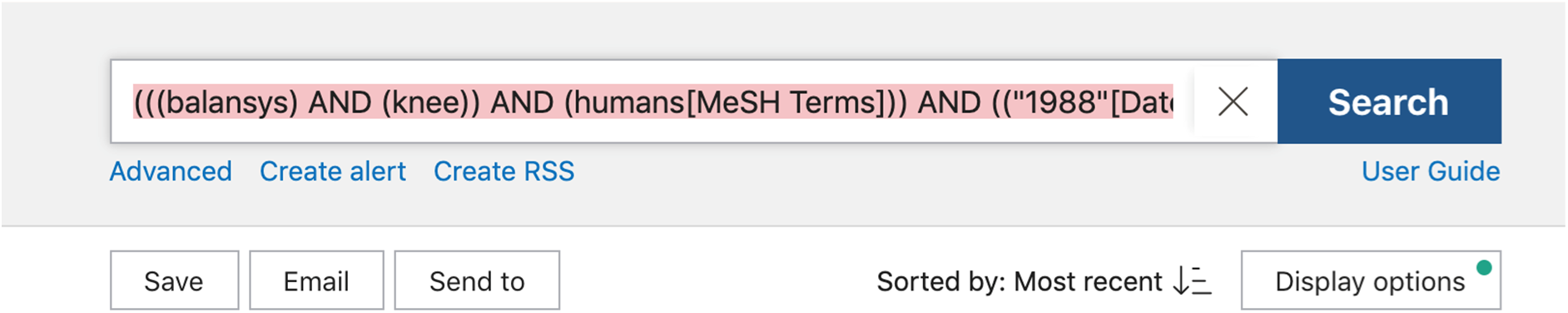

Web of Science: 7

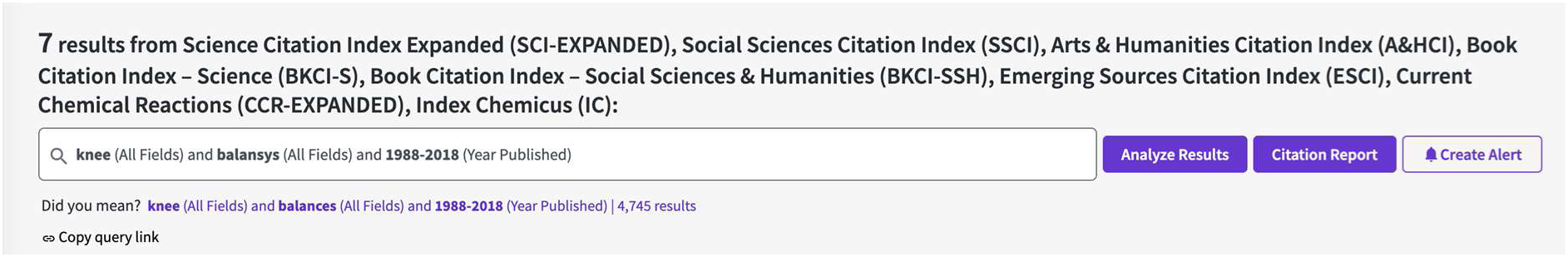

#### Sigma high performance partial knee 13^th^ October

Embase: 37

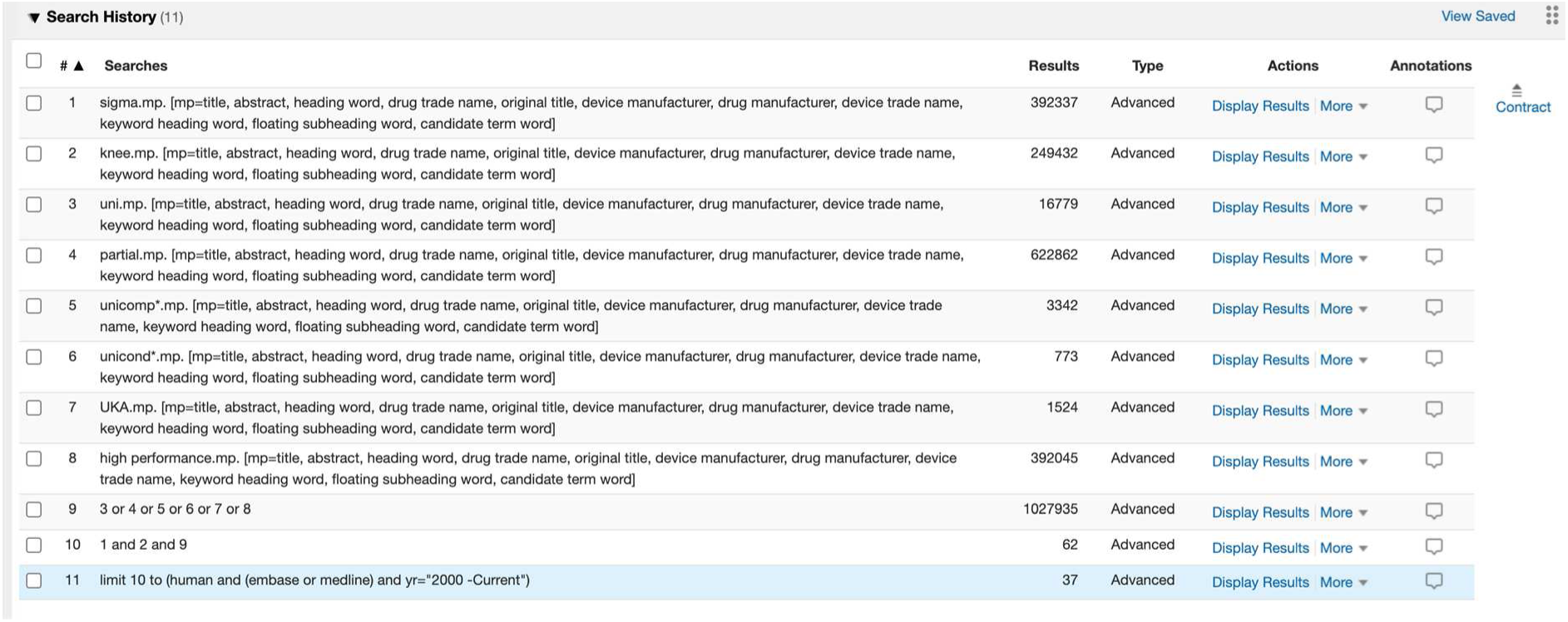

PubMed: 7

((((knee) AND (uni OR UKA OR unicomp* OR unicond* OR “high performance”)) AND (sigma)) AND (humans[MeSH Terms])) AND ((“2000”[Date - Publication]: “3000”[Date - Publication]))

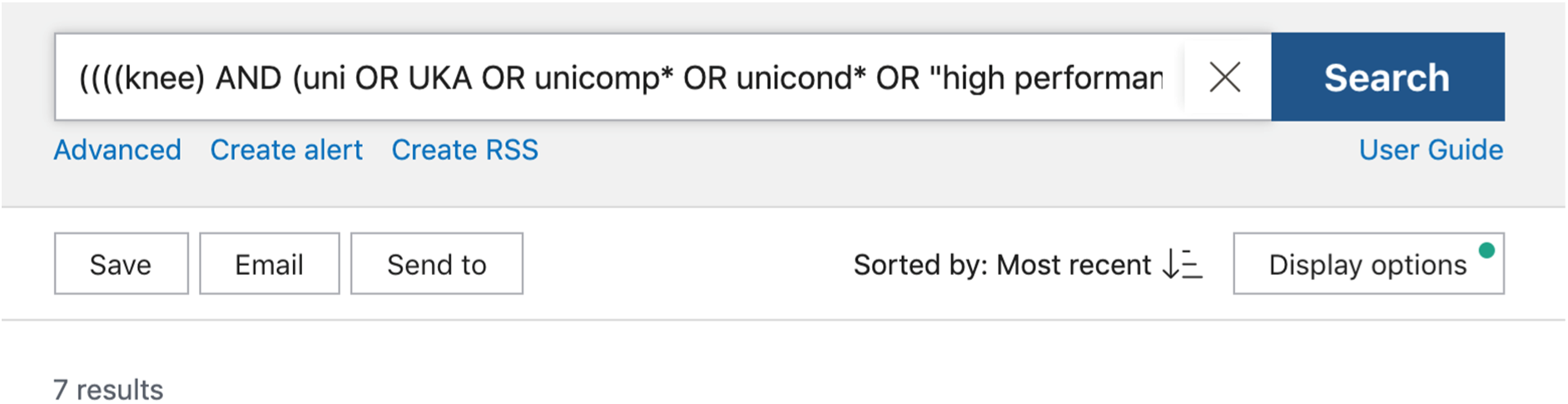

Web of Science: 7

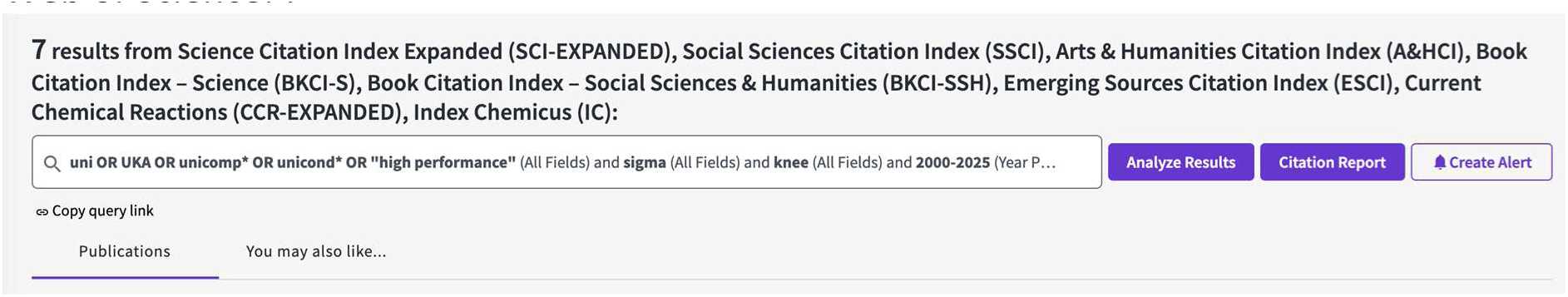

#### Innex 13^th^ October 2021

Embase: 22

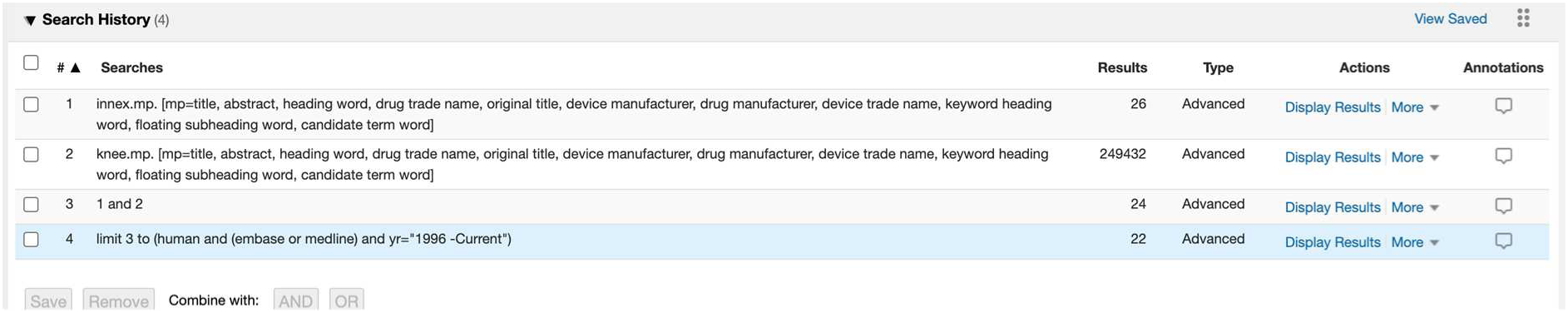

PubMed: 8

(((innex) AND (knee)) AND (humans[MeSH Terms])) AND ((“1996”[Date - Publication]: “3000”[Date - Publication]))

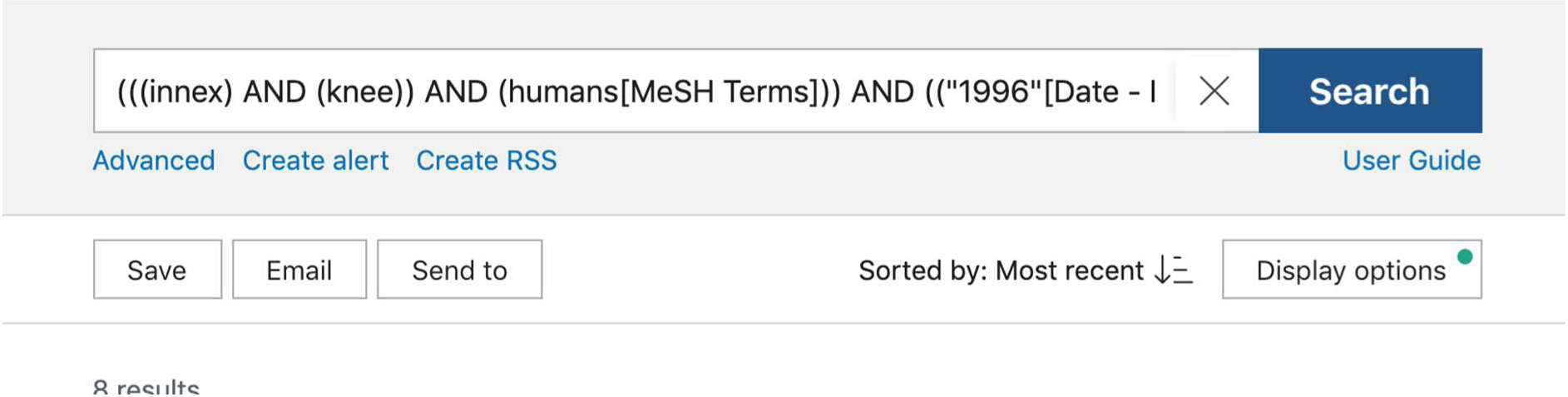

Web of Science: 10

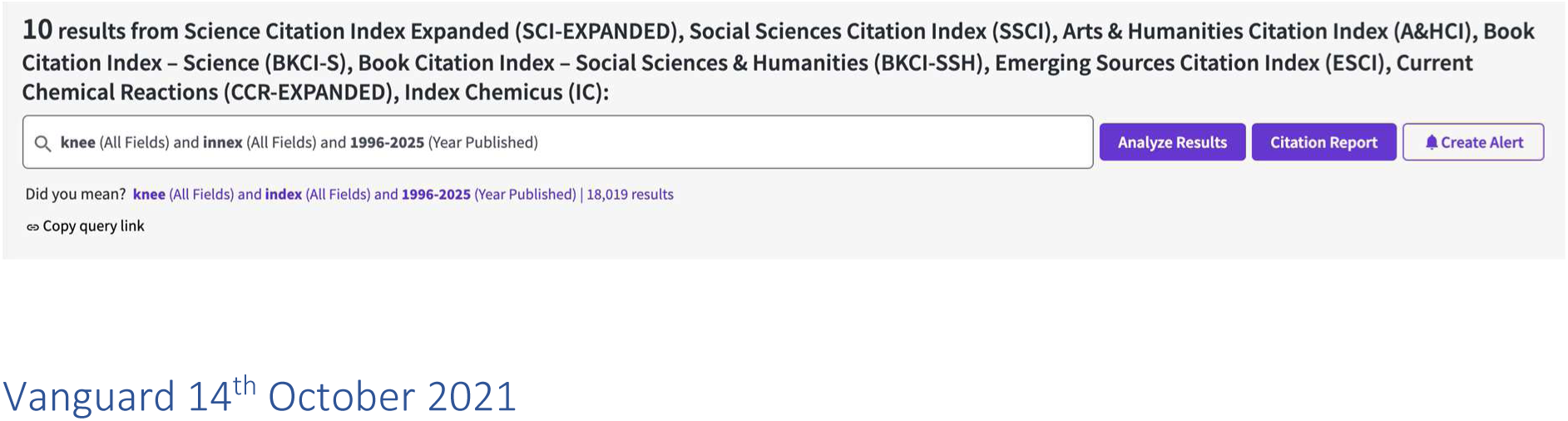

#### Vanguard 14^th^ October 2021

Embase: 211

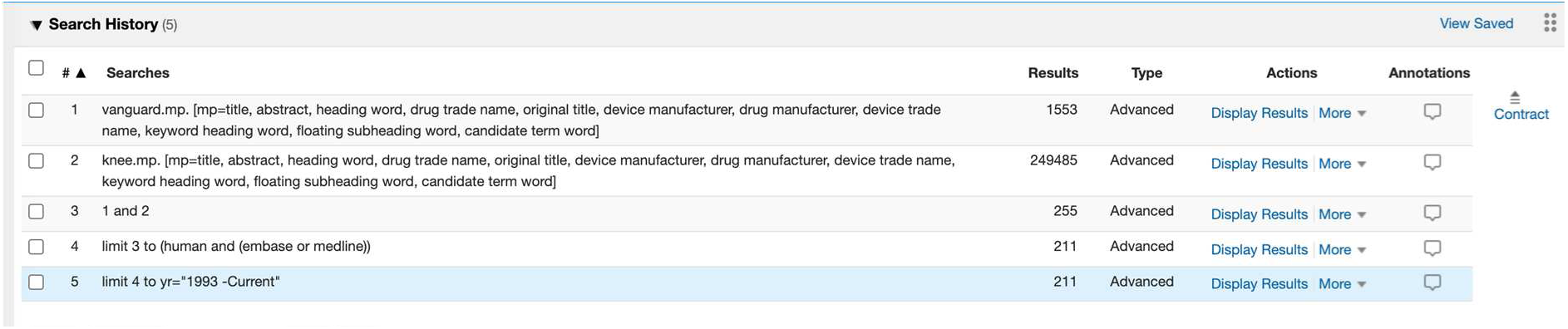

PubMed: 61

(((knee) AND (vanguard)) AND (humans[MeSH Terms])) AND ((“1993”[Date - Publication]: “3000”[Date - Publication]))

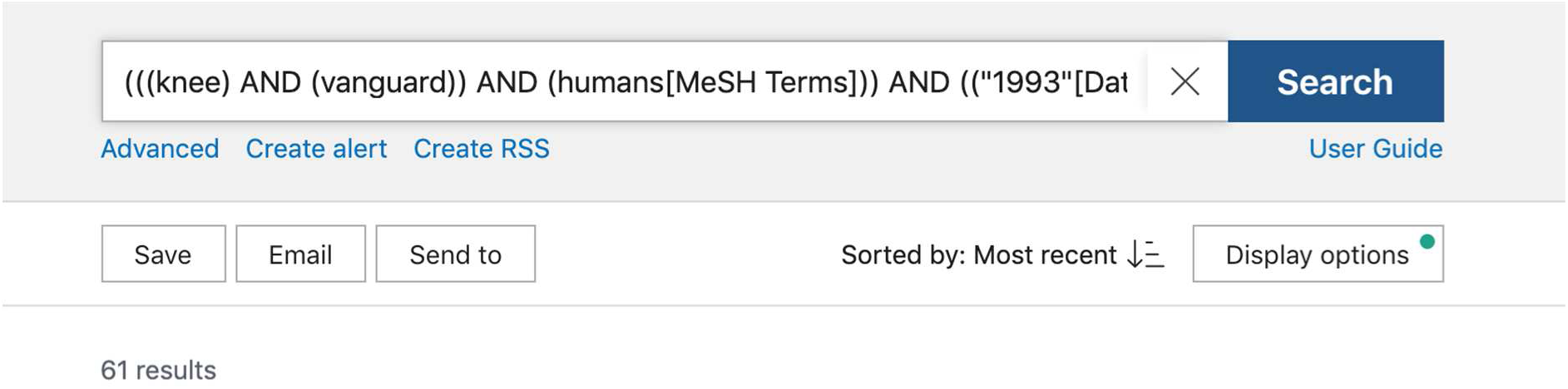

Web of Science: 77

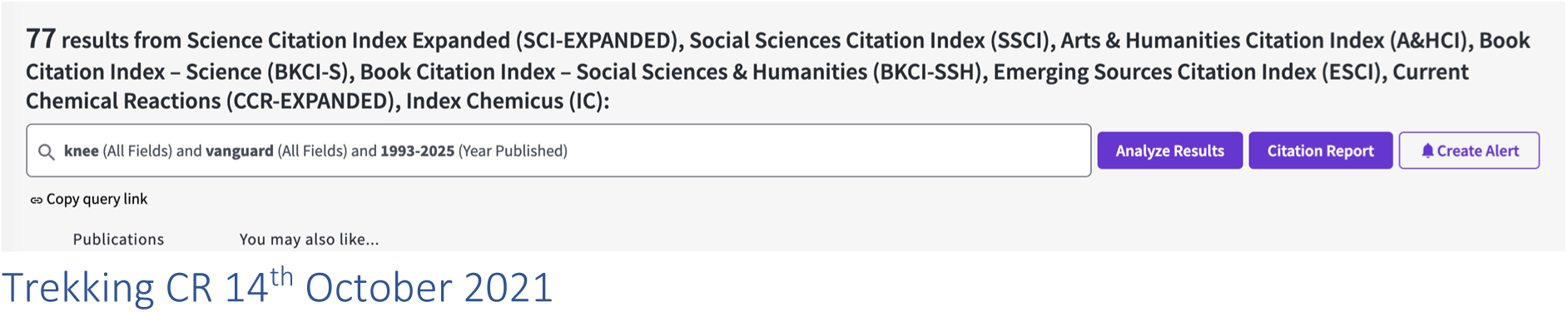

#### Trekking CR 14^th^ October 2021

Embase: 13

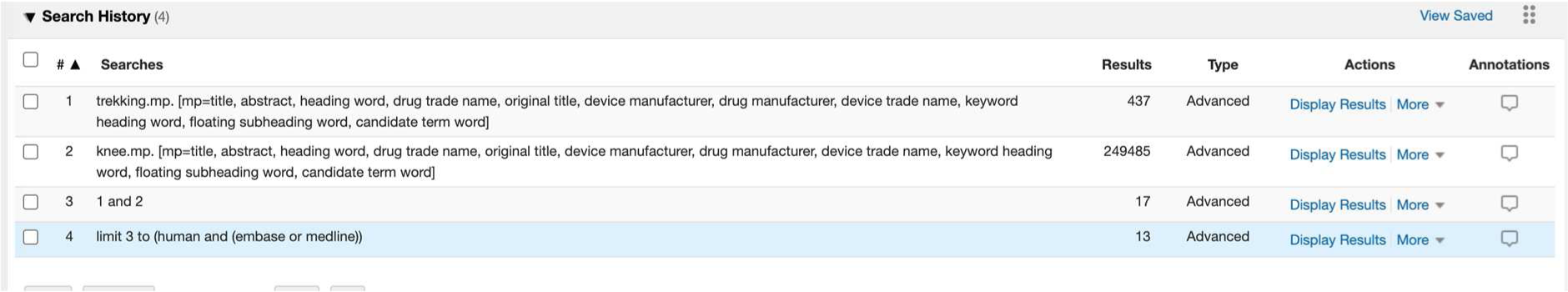

PubMed: 7

((trekking) AND (knee)) AND (humans[MeSH Terms])

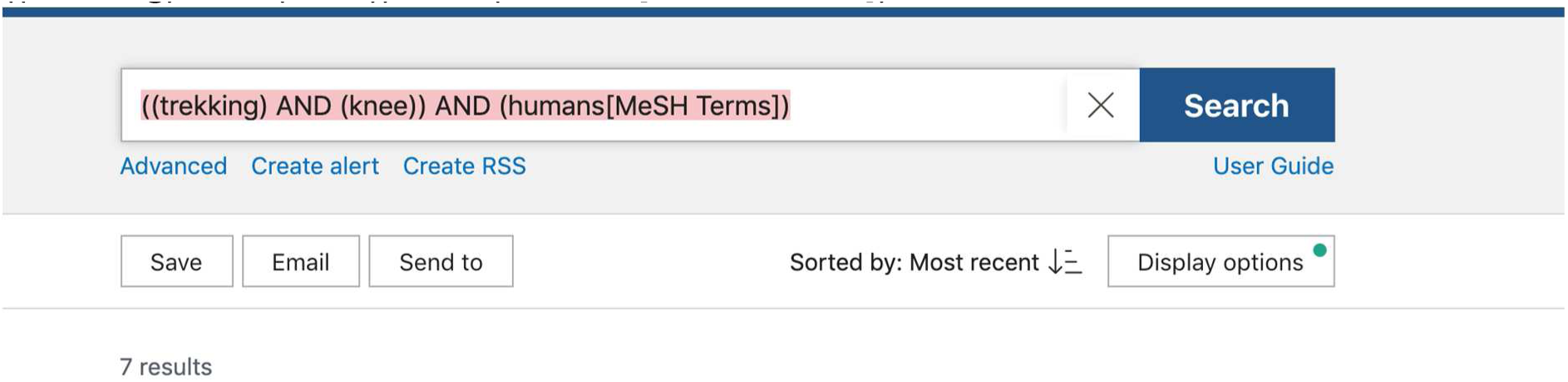

Web of Science: 18

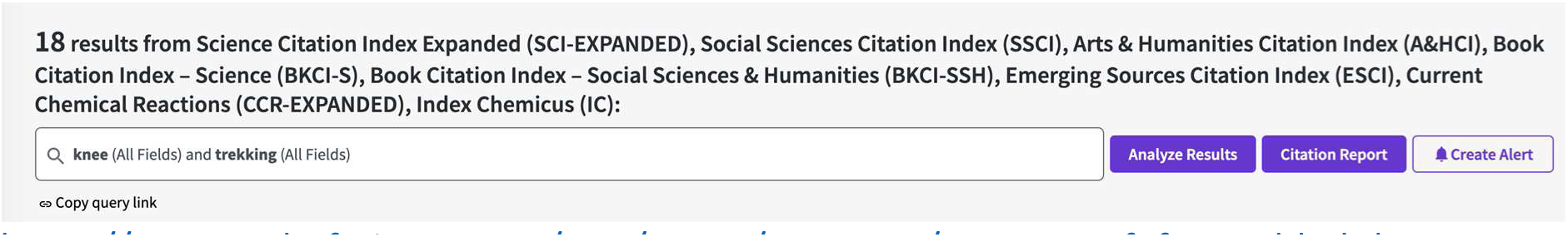

https://www.webofscience.com/wos/woscc/summary/04743834-f0f0-4c2d-b0bd-3c4490712c5f-0d1361c5/relevance/1

#### Nexgen 14^th^ October 2021

Embase: 371

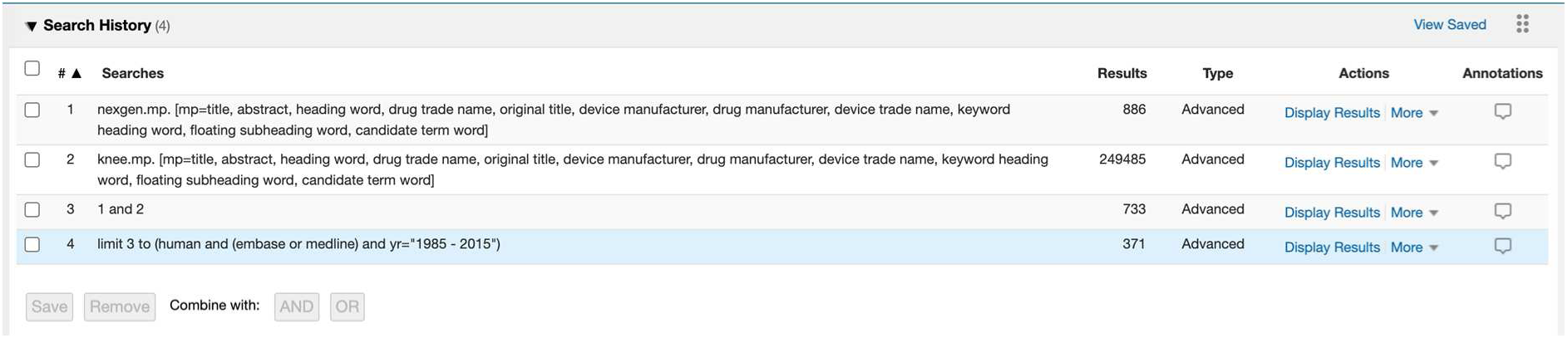

PubMed: 132

(((nexgen) AND (knee)) AND (humans[MeSH Terms])) AND ((“1985”[Date - Publication]: “2015”[Date - Publication]))

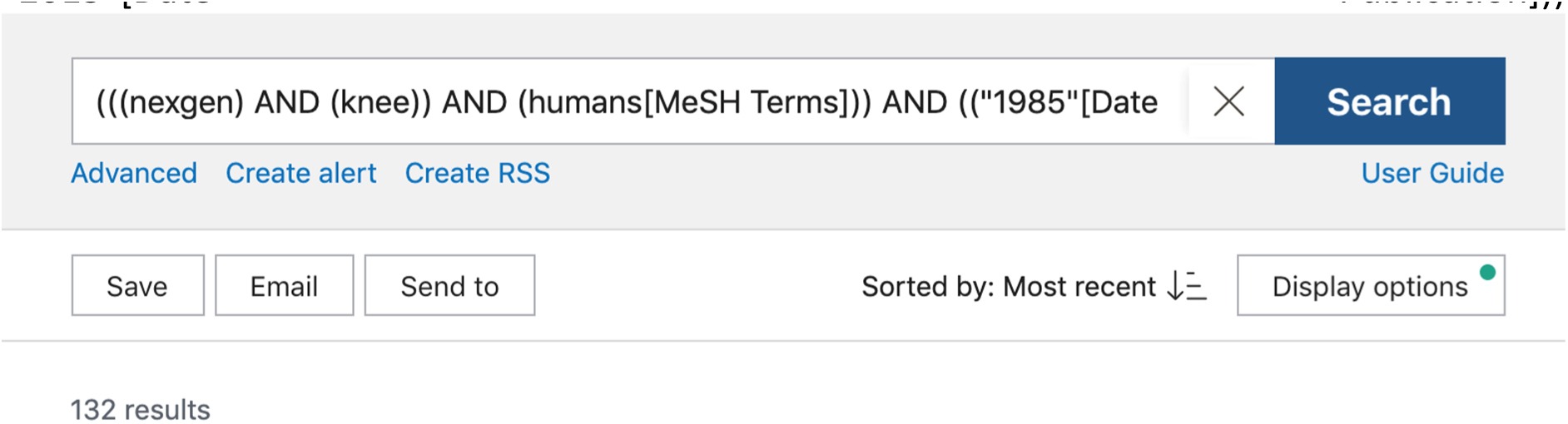

Web of Science: 113

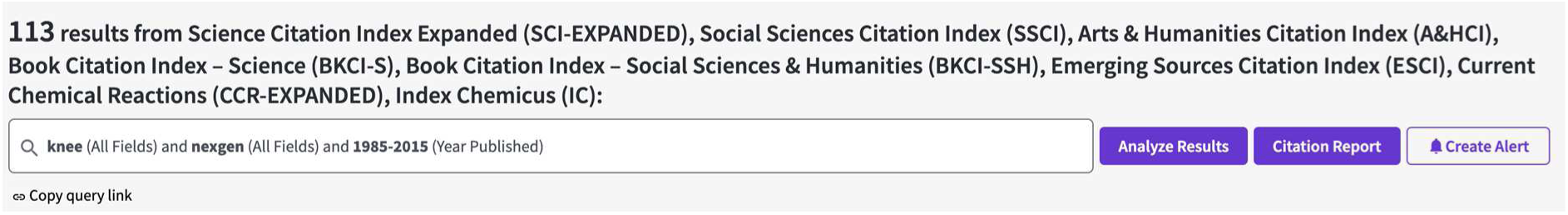

#### Optetrak CR 8^th^ December

Embase: 25

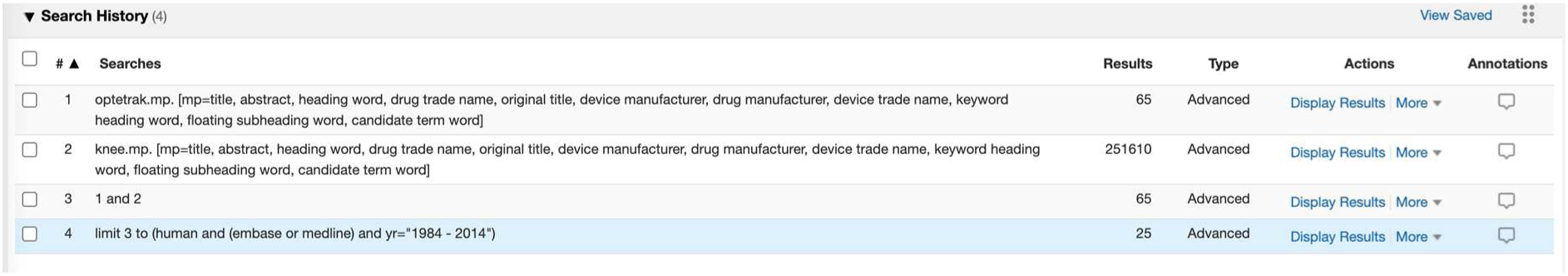

PubMed: 14

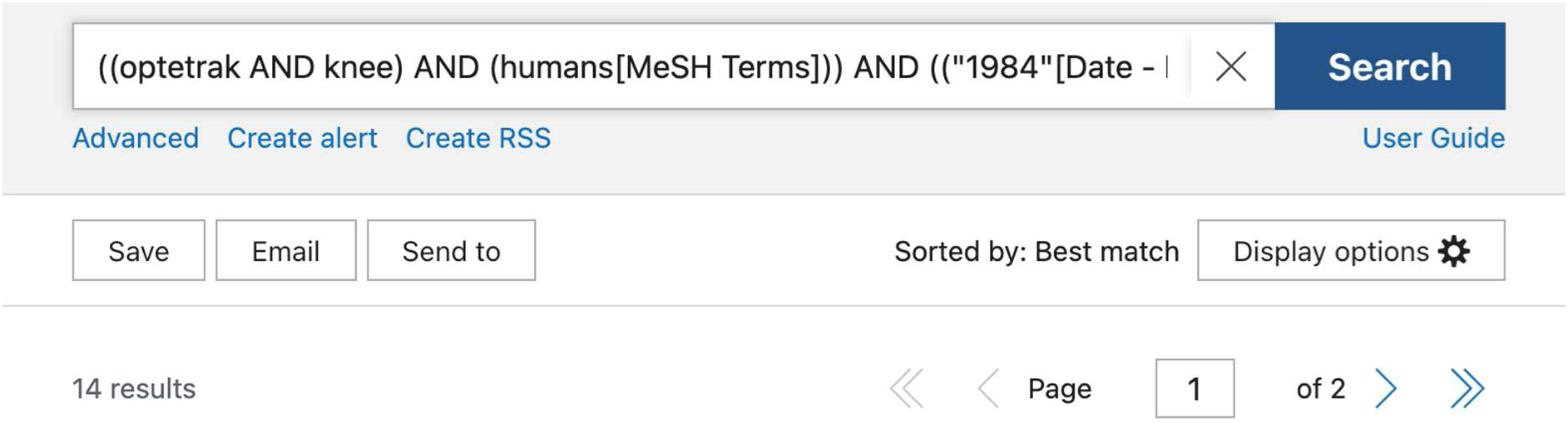

((optetrak AND knee) AND (humans[MeSH Terms])) AND ((“1984”[Date - Publication]: “2014”[Date - Publication]))

Web of Science: 13

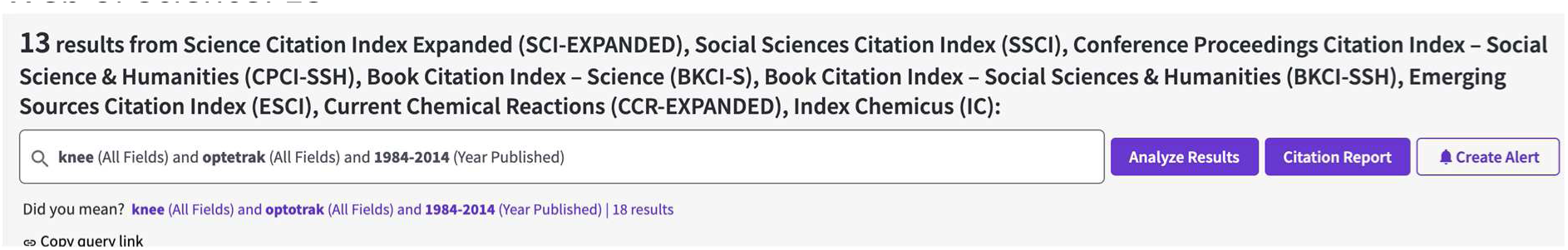

#### ACS unc 17^th^ Dec 2021

Embase: 6

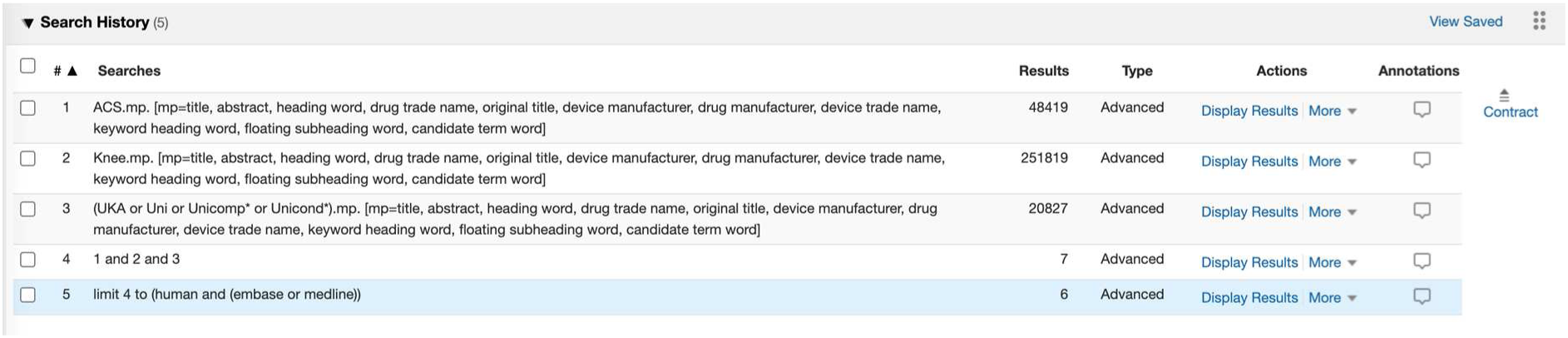

PubMed: 9

(((knee) AND (uni OR UKA OR unicomp* OR unicond*)) AND (ACS)) AND (humans[MeSH Terms])

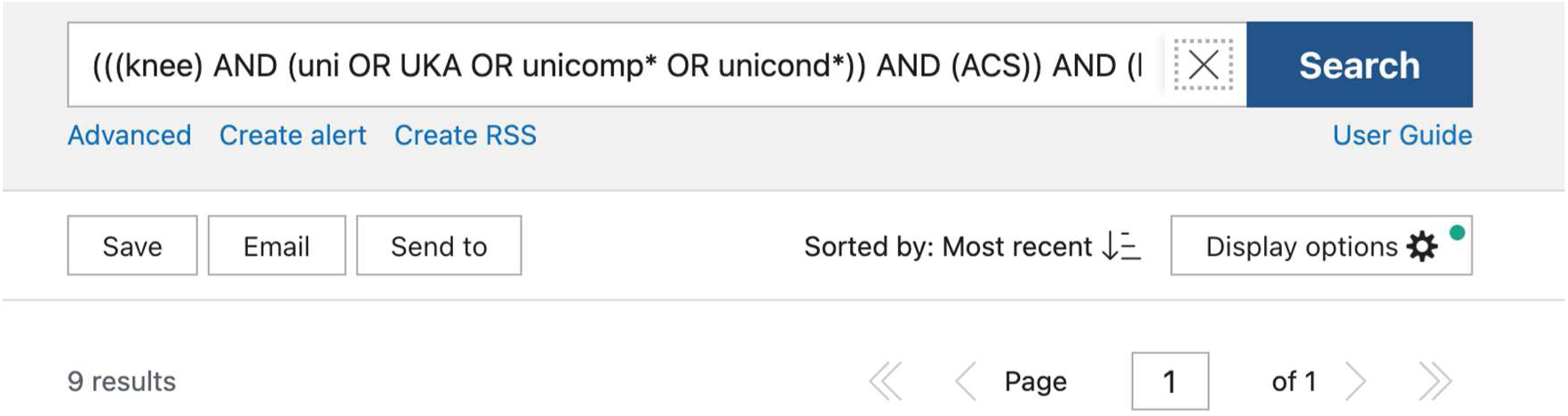

Web of Science: 7

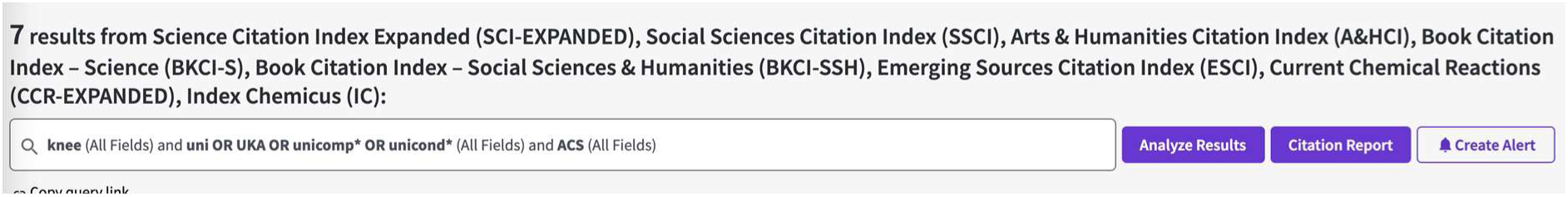

#### Logic Exactech 17^th^ December 2021

Embase: 77

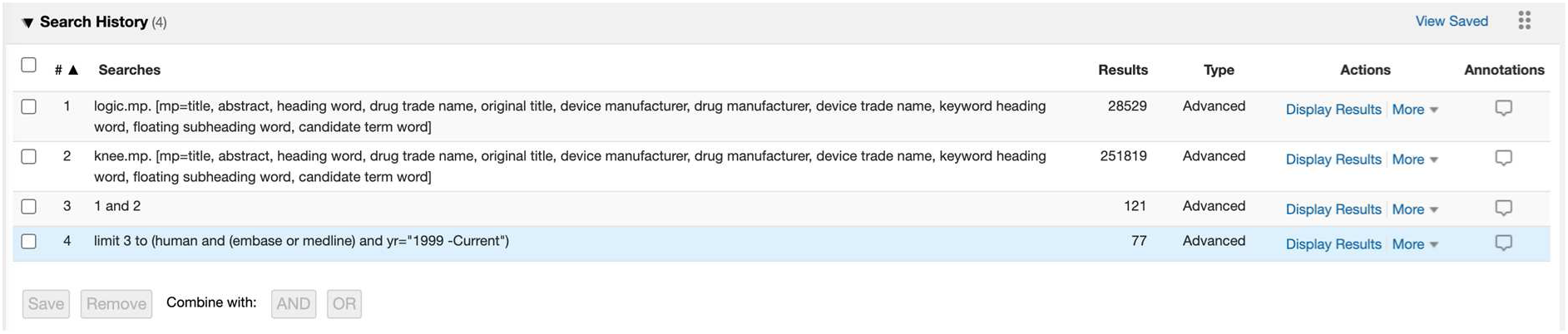

PubMed: 136

(((logic) AND (knee)) AND (humans[MeSH Terms])) AND ((“1999”[Date - Publication]: “3000”[Date - Publication]))

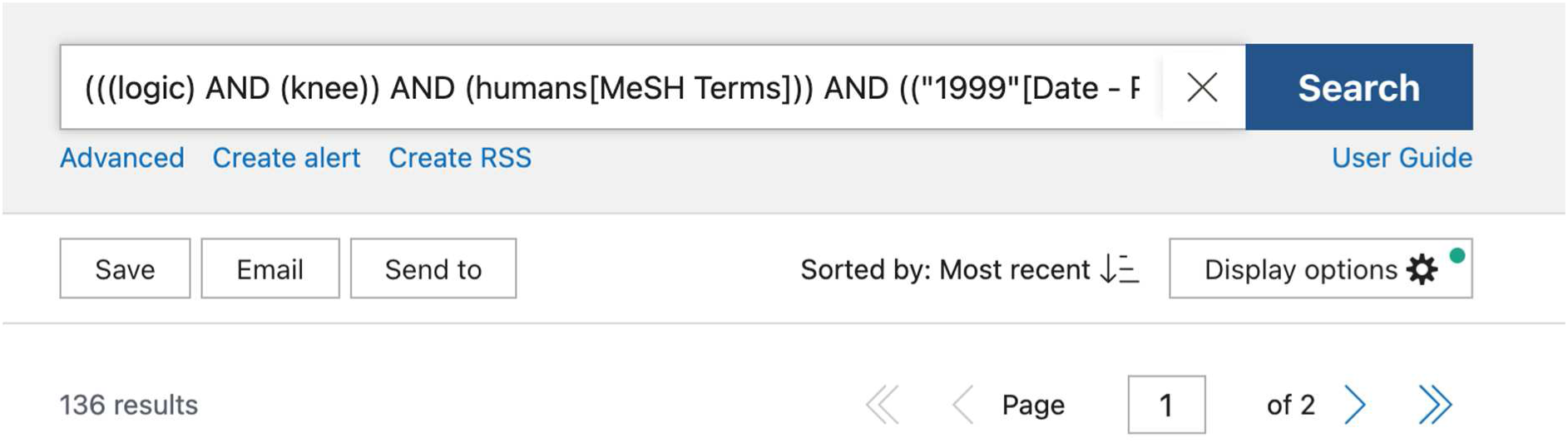

Web of Science: 78

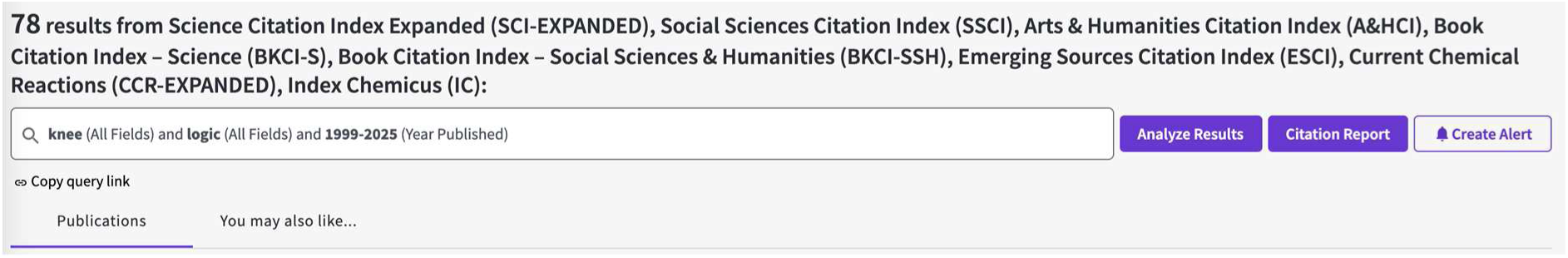

#### LCS complete 18^th^ December 2021

Embase: 48

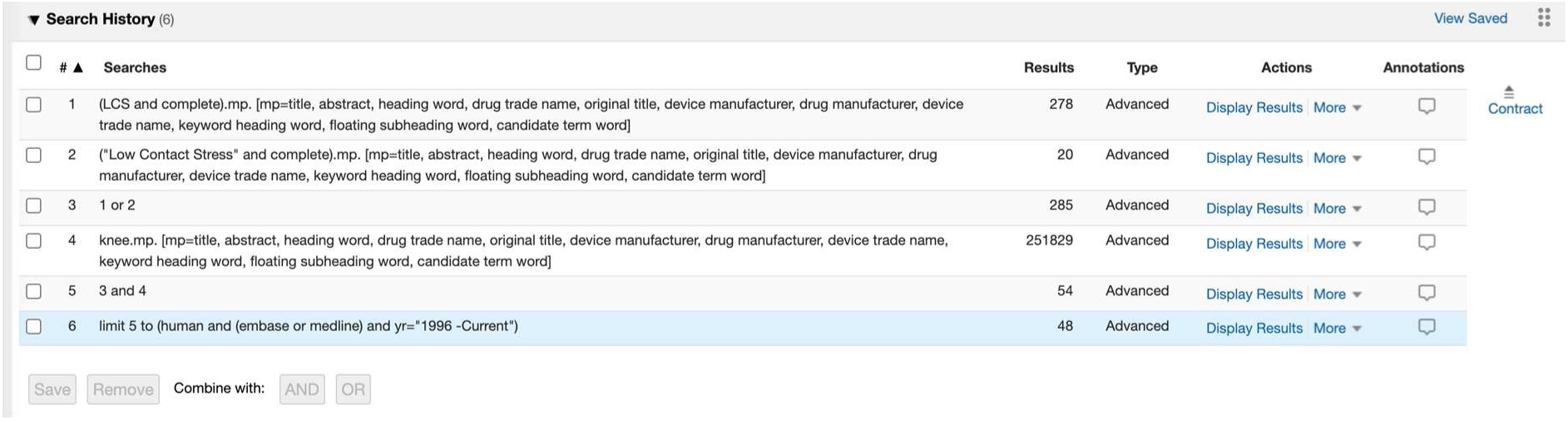

PubMed: 18

((((LCS AND complete) OR (“Low contact stress” AND complete)) AND (knee)) AND (humans[MeSH Terms])) AND ((“1996”[Date - Publication]: “3000”[Date - Publication]))

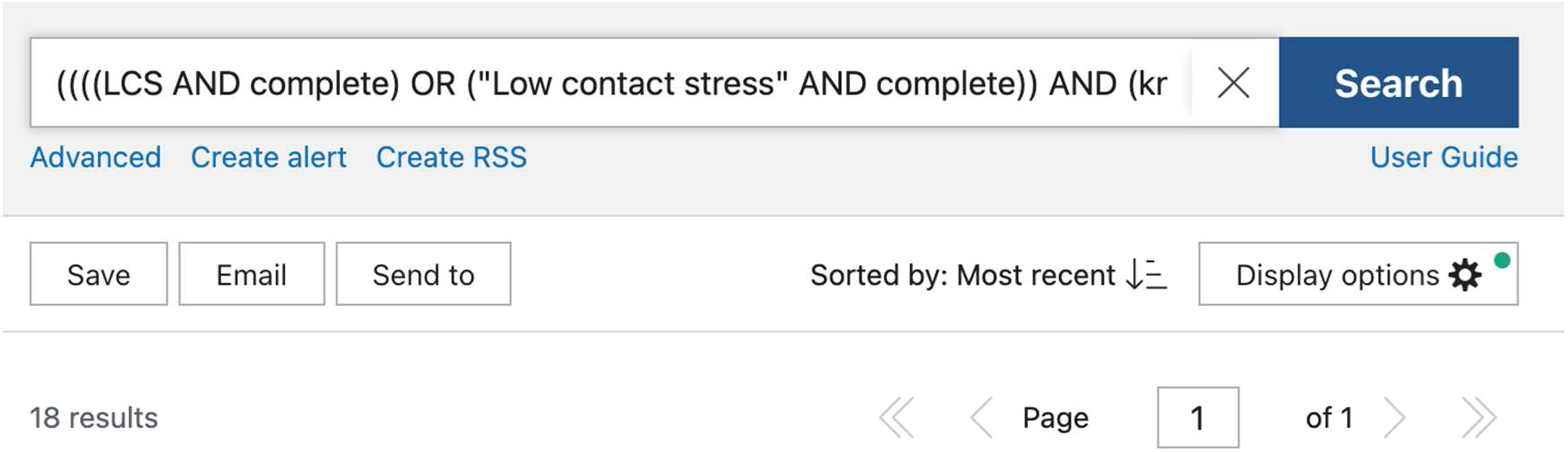

Web of Science: 22

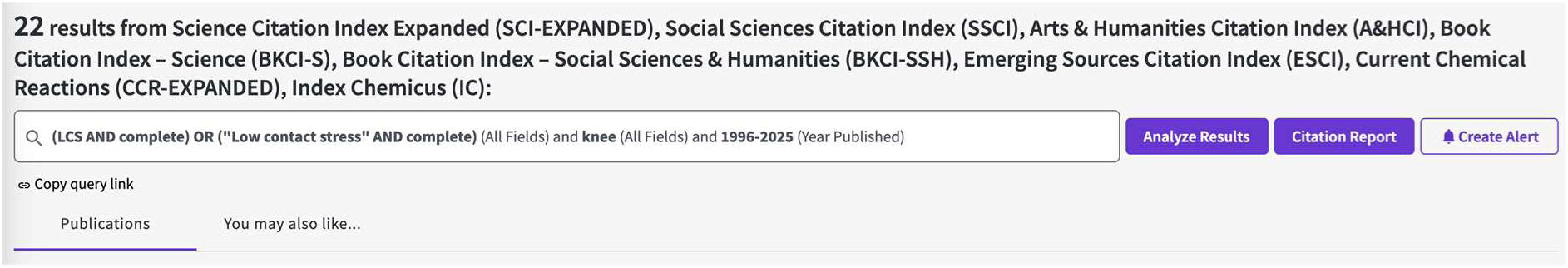

## Appendix III: List of data items that were collected

### Devices

The following information will be included for each device included in the study.

- Name of device
- Manufacturer
- Implant reference number, if available
- Date of CE approval
- Date of FDA approval
- Date of first use (ODEP)

Procedure:

For knee arthroplasty:
  - Stability: Cruciate ligament preserved (yes/no), Medial Pivot design, other or not recorded
  - Mobility: fixed bearing /mobile bearing/ or not recorded
  - Fixation (all cemented/all uncemented/other) or not recorded
  - Patella resurfaced (yes/no) or not recorded
For to hip arthroplasty:
  - Fixation of stem/cup (cemented yes/no) or not recorded
  - Type of bearing surface or not recorded
Associated stem/cup (free text)
Fixation associated (free text)
  - Type of bearing or not recorded (yes/no)

### Papers

This section details the data that will be extracted from each paper identified in our literature search. We will not contact authors of papers for additional information not present in the papers because we are interested in assessing the published evidence rather than evidence that may have been generated but is not published.

Meta-data:

- First author
- Date of publication (first available online if available)
- Date publication first available
- Submission (or publication) before /after CE mark date
- Submission (or publication) before /after FDA approval date
- Journal
- Study location(s) (continent)
- First and last year of recruitment

Objective (free text, copied from paper)

Key finding (free text, copied from paper)

Study characteristics

- Study type (cohort, registry-based cohort, case control, randomised controlled trial, case series or reports)
- Retrospective, prospective, both elements
- Population-based or specific population (e.g. young patients only)
- Real-world or experimental setting
- Comparative study (yes/no)
- Which comparison implant/group (e.g. established vs. new)
- Study aim (superiority/non-inferiority)
- Randomisation (yes/no)
- Blinding (select from: participant, investigator, outcome assessor)
- Type of RCT (registry-nested, other)
- Clinical trial registration ID provided (yes/no)

Patient characteristics

- Number in study
- Number in device in question arm
- Age (mean/median)
- Women (%)
- Diagnostic (% primary OA)

Investigators and sponsors:

- Author affiliations (academic, industry, mix)

Outcomes reported:

- All-cause revision as outcome (yes/no)
  ◦ Revision rate at x years (upper CI, lower CI)
- Imaging (yes/no) if yes, which method
  ▪ Radiograph
  ▪ CT
  ▪ MRI
  ▪ EOS
  ▪ RSA
  ◦ Migration (yes/no)
  ◦ Osteolysis (yes/no)
  ◦ Other (yes/no)
- Patient reported outcome measures (yes/no)
  ◦ Oxford knee score (yes/no)
  ◦ Knee Injury and Osteoarthritis Outcome Score (KOOS) (yes/no)
  ◦ Oxford hip score (yes/no)
  ◦ Hip disability and osteoarthritis outcome score (HOOS) (yes/no)
  ◦ WOMAC (yes/no)
  ◦ EQ-5D (yes/no)
  ◦ SF-36/SF-12 (yes/no)
- Performance (yes/no)
  ◦ Gait (yes/no)
  ◦ Flexion (yes/no)
  ◦ Posterior stability (yes/no)
  ◦ Other (yes/no)

Are analyses stratified or outcomes presented by gender (yes/no) Are analyses stratified or outcomes presented by age (yes/no)

Safety:

- Did paper report safety concerns? (yes/potential/no) if yes, which:
  ◦ Higher revision rate
  ◦ Imaging abnormality
  ◦ Inferior clinical results
  ◦ PROMS
  ◦ Biomechanical

Safety concern reported in which section of paper (e.g. abstract, discussion)

We will also record

- Mean and max. length of follow-up
- Adverse events/complications
  ◦ Infection (N and %)
  ◦ Dislocation (N and %)
  ◦ Fracture (N and %)
  ◦ Thromboembolic event (N and %)
  ◦ Myocardial infarction (N and %)
- Mortality (N and %)

#### Risk of bias

Attrition

- Lost to follow-up (N and % [per group if comparative])
  ◦ Reasons for loss mentioned yes/no

Information bias

- Exposure identification = Procedure (see above) details provided yes/no
- Outcome definition provided yes/no
- Response rate PROs (see above)

Selection bias in observational comparative studies (for RCTs bias assessed above)

- Measures used to reduce bias yes/no and which: Adjustment/Restriction/Matching

#### All-cause revision

The following will be extracted per device:

- Number of devices included
- Total number of observed events
- Timing of measurement (all time points available)
- Point estimates (cumulative incidence of revision or cumulative survival)
- Confidence intervals

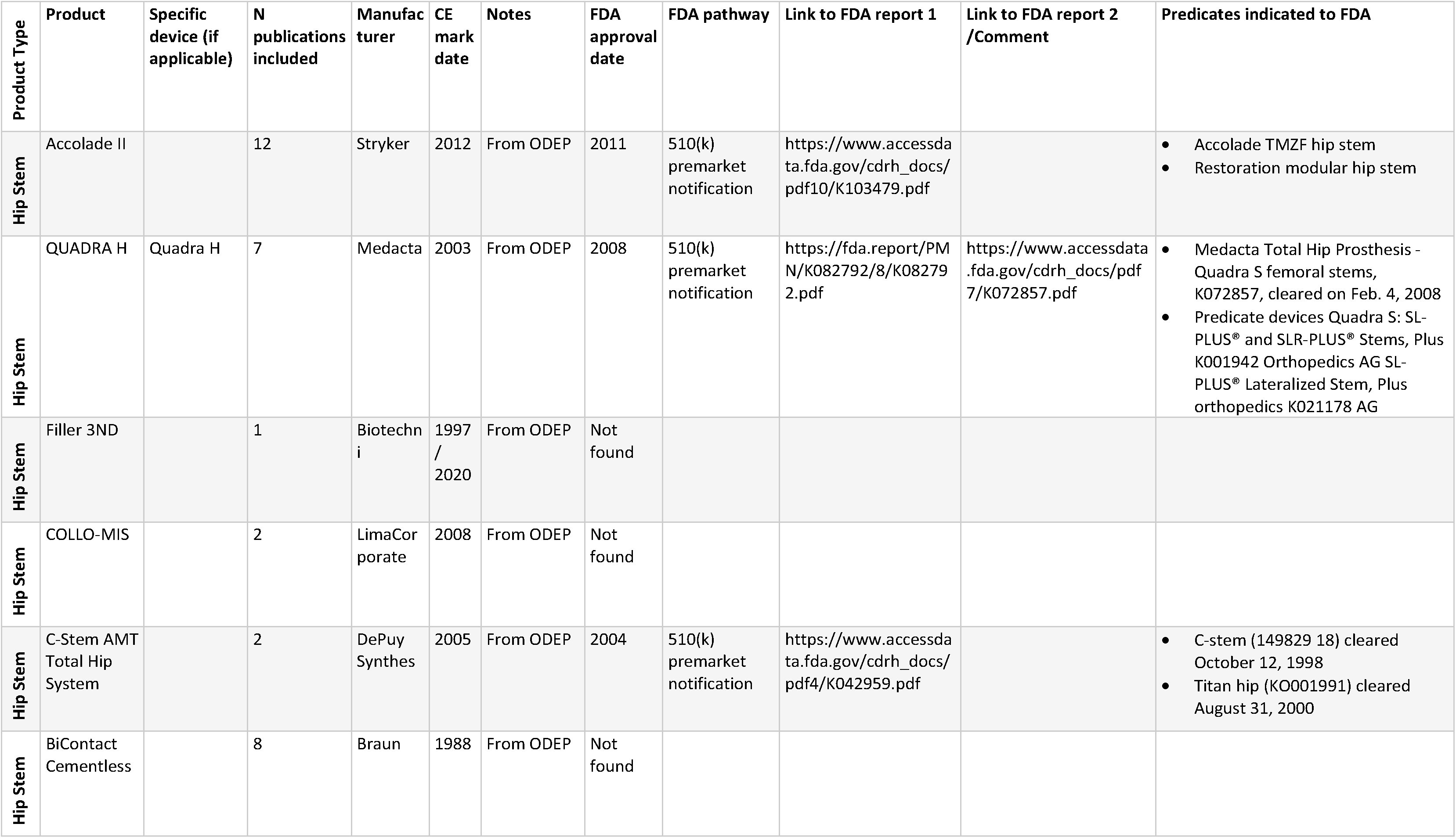

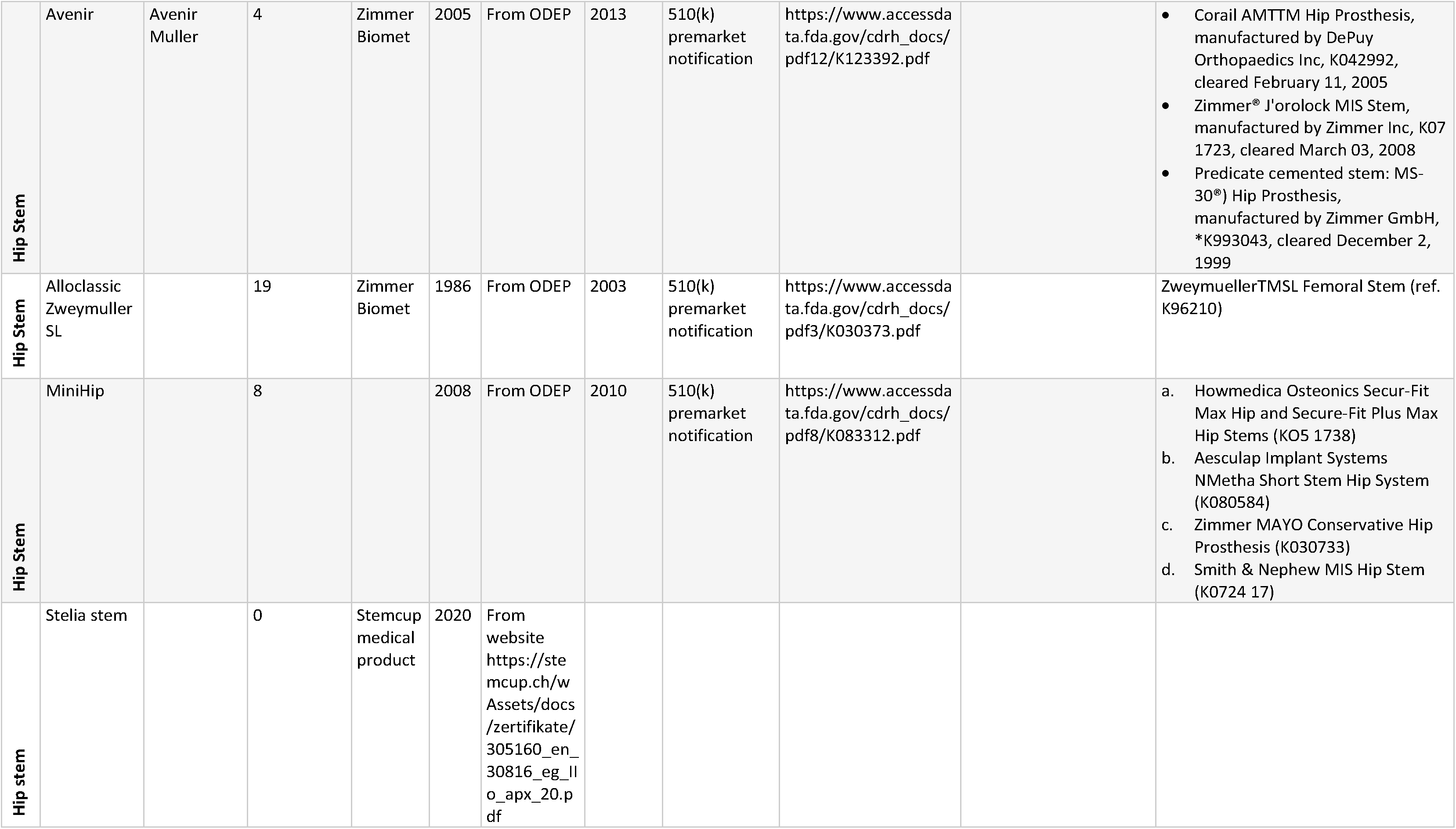

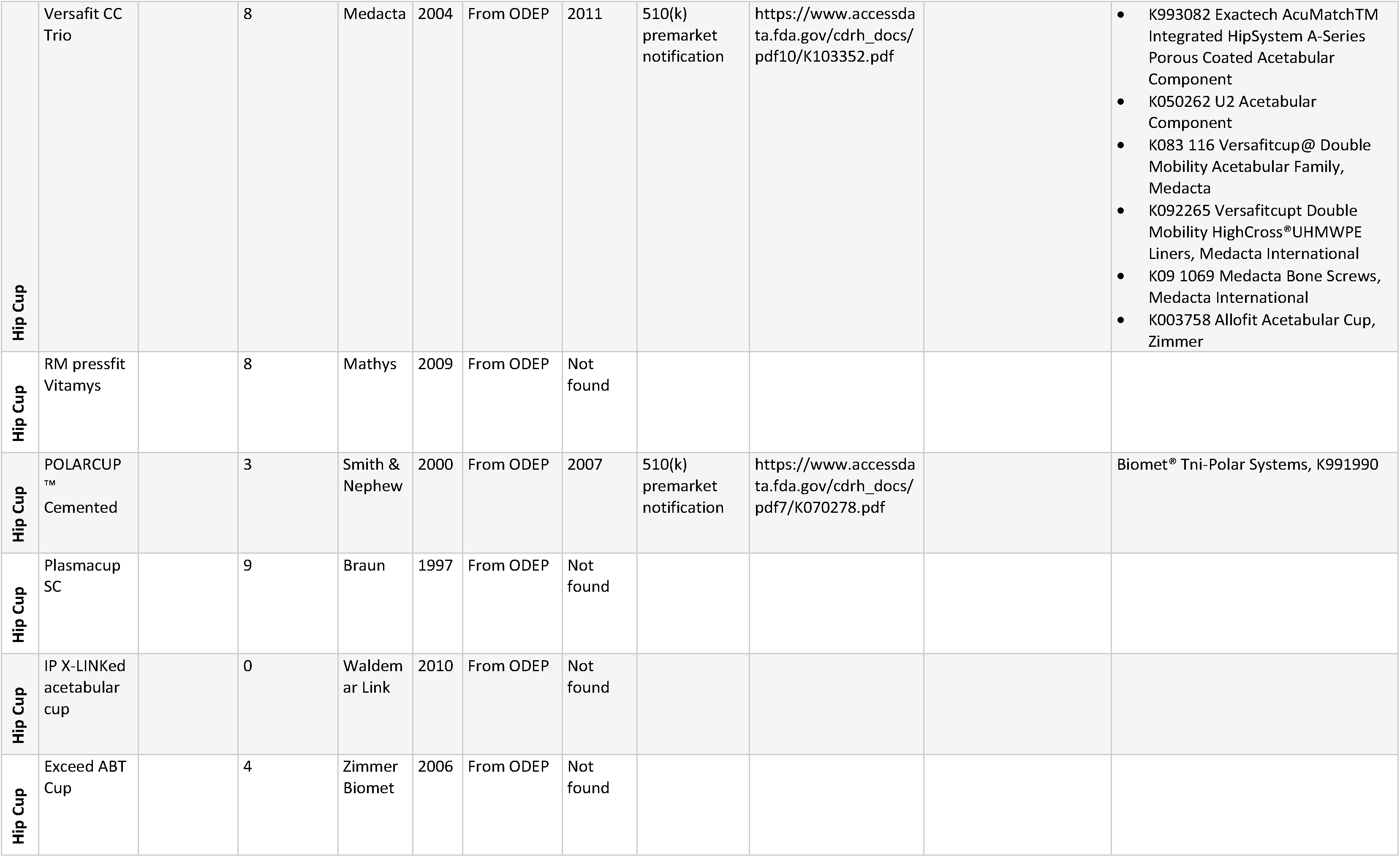

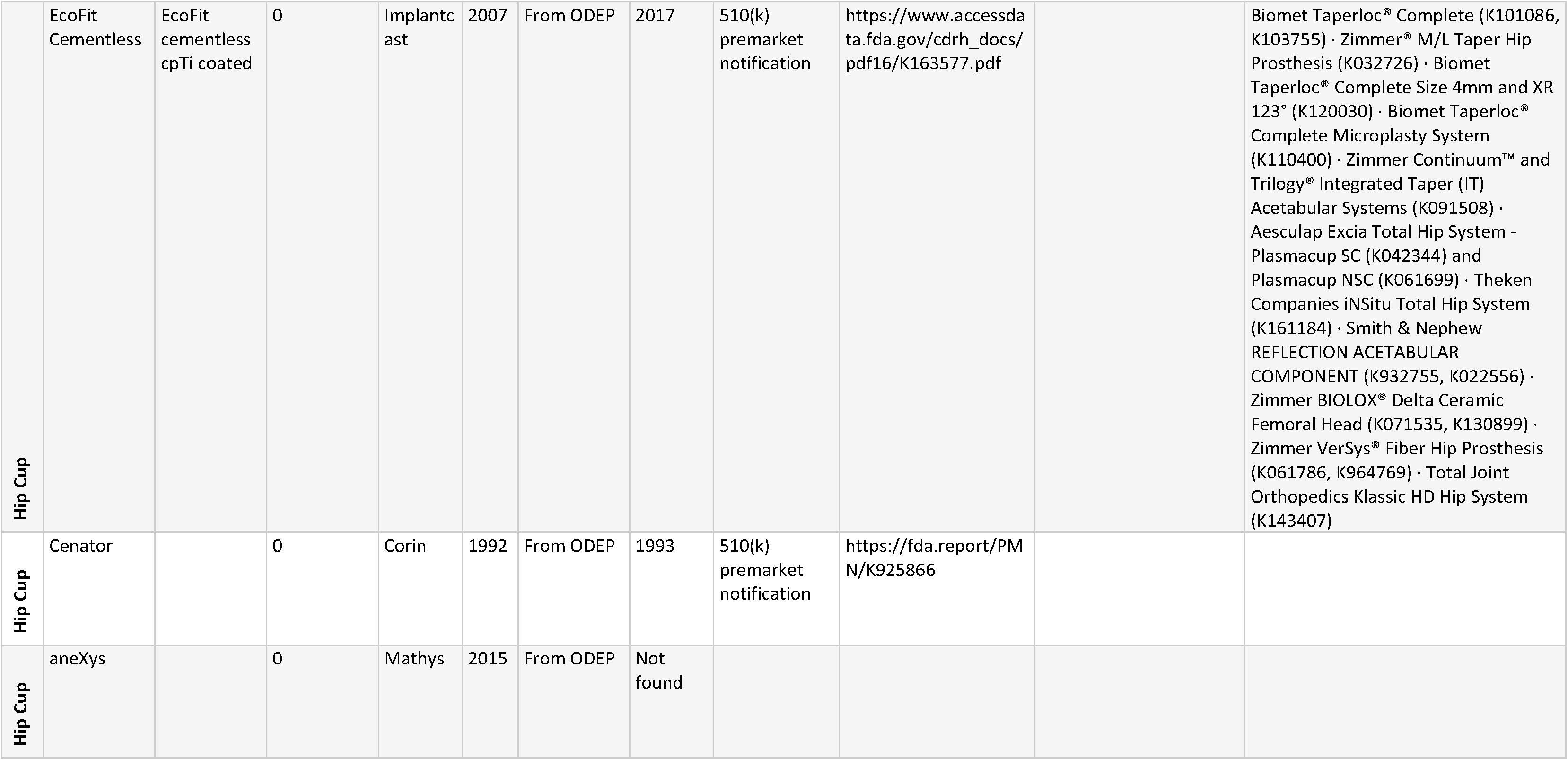

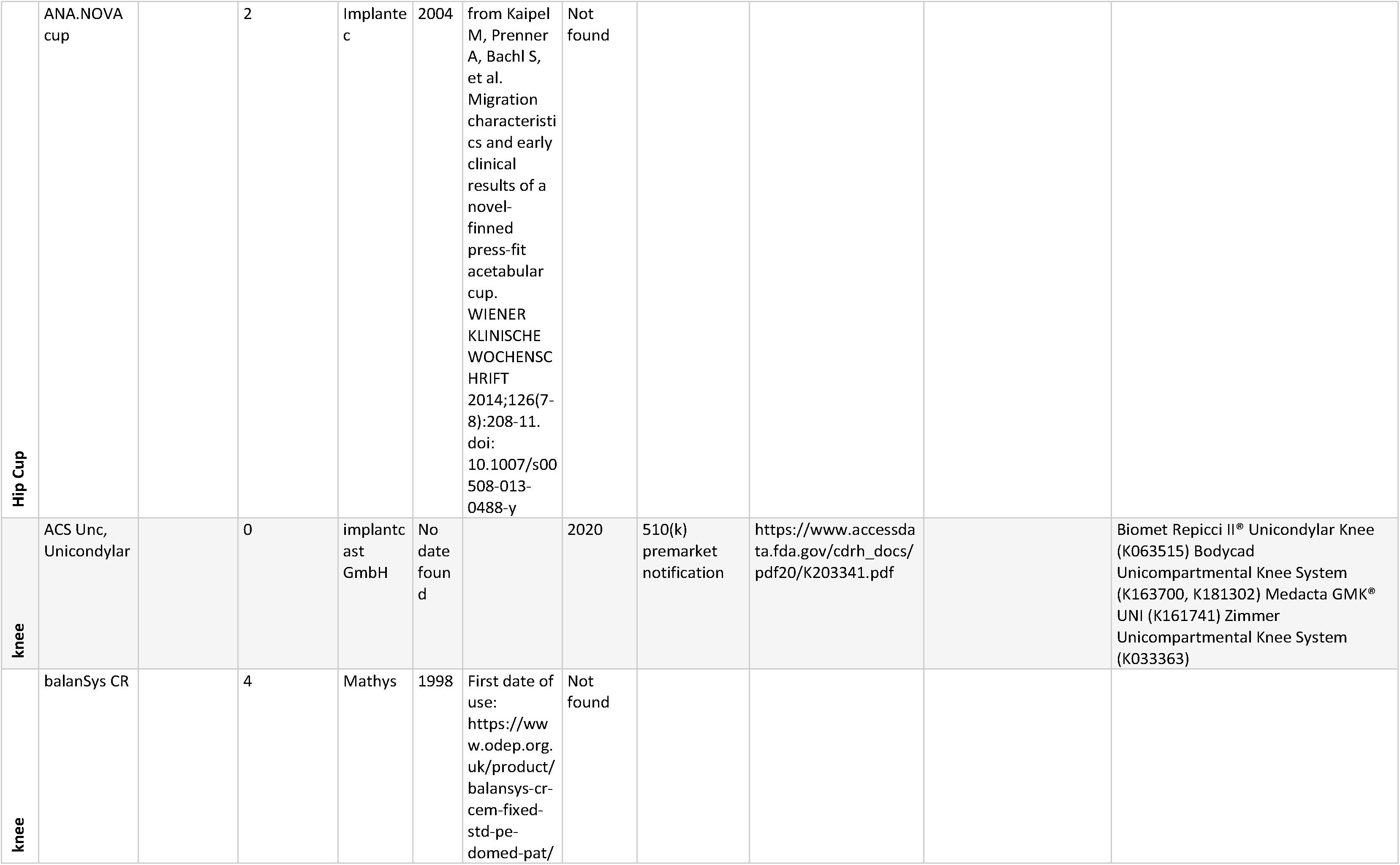

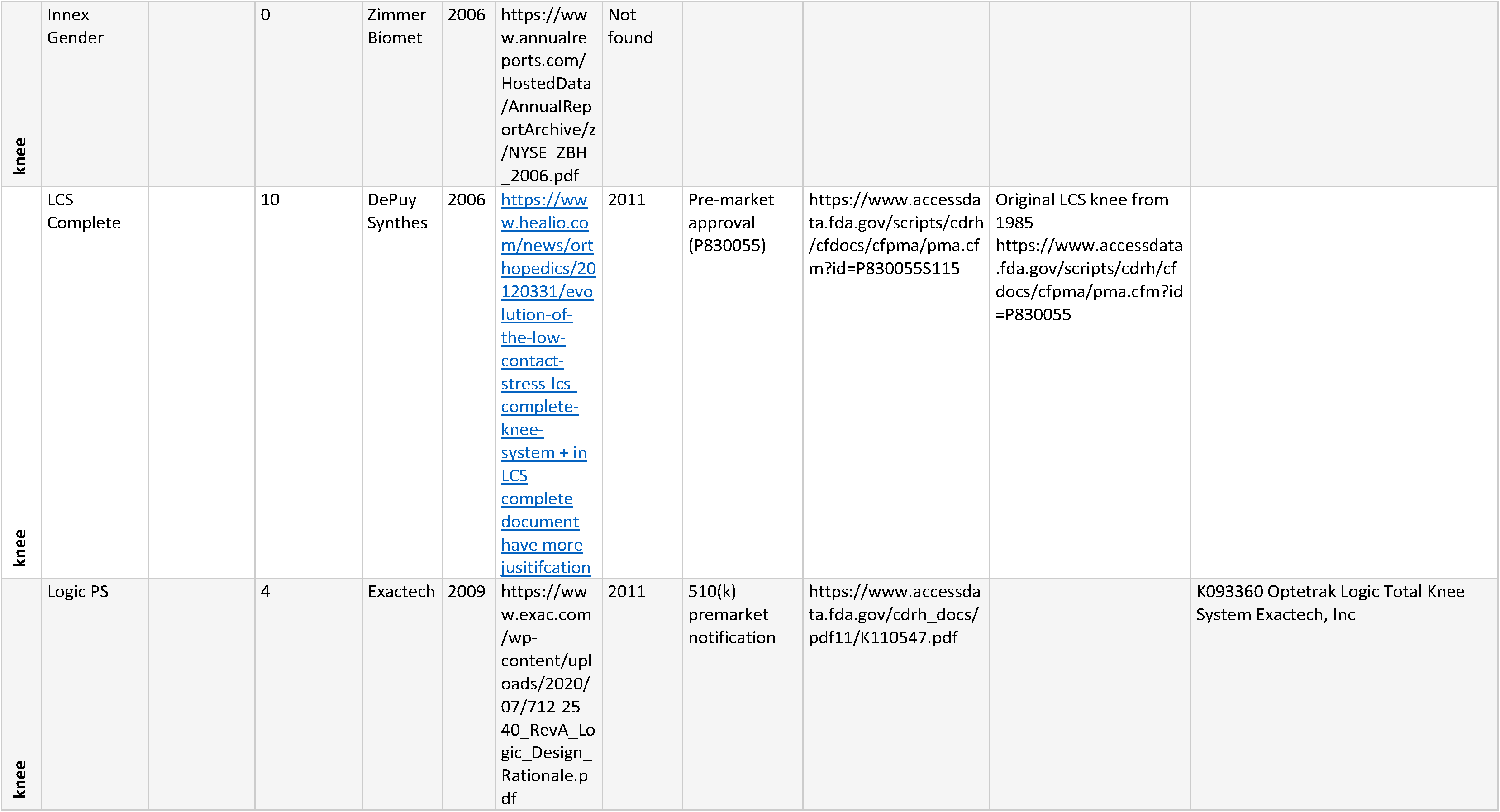

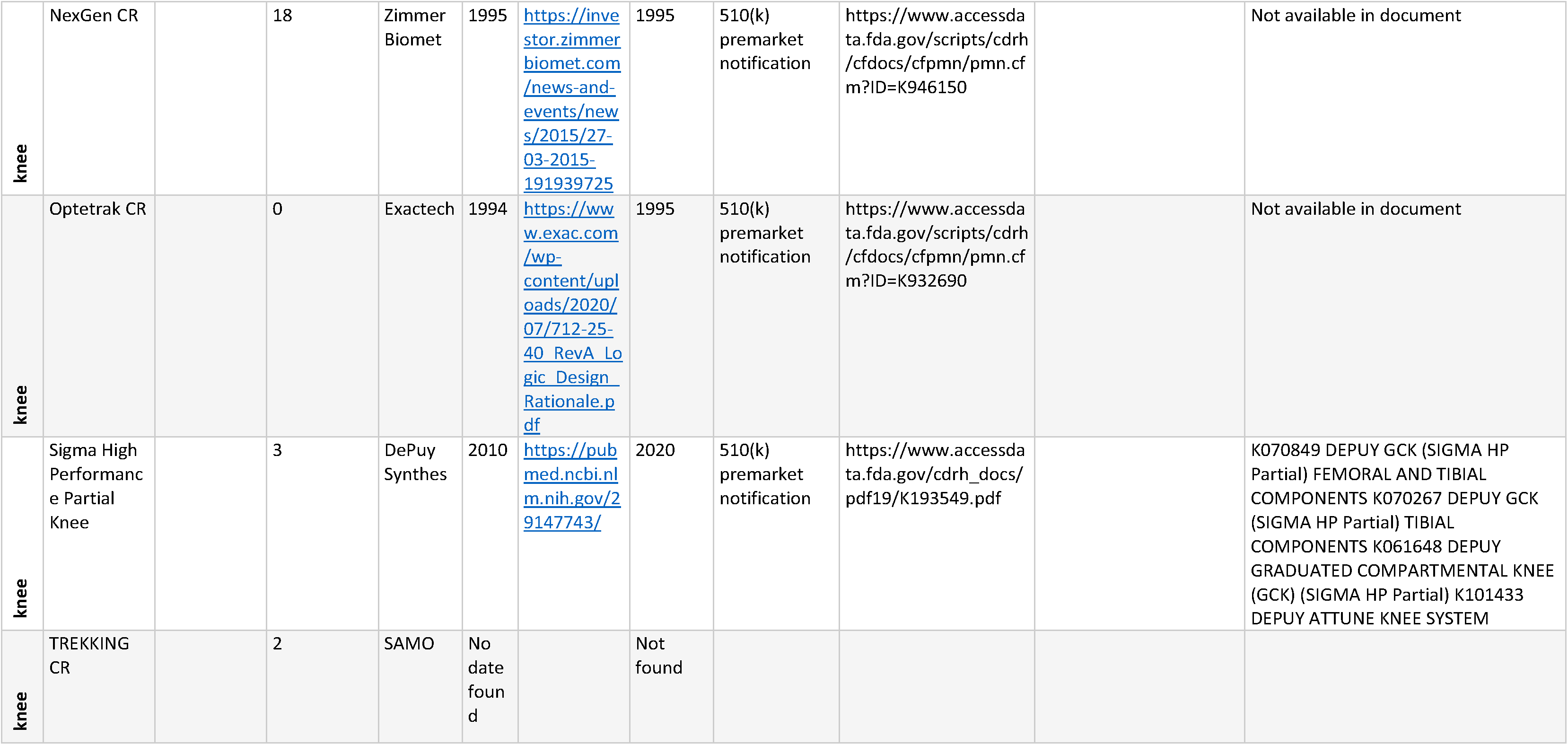

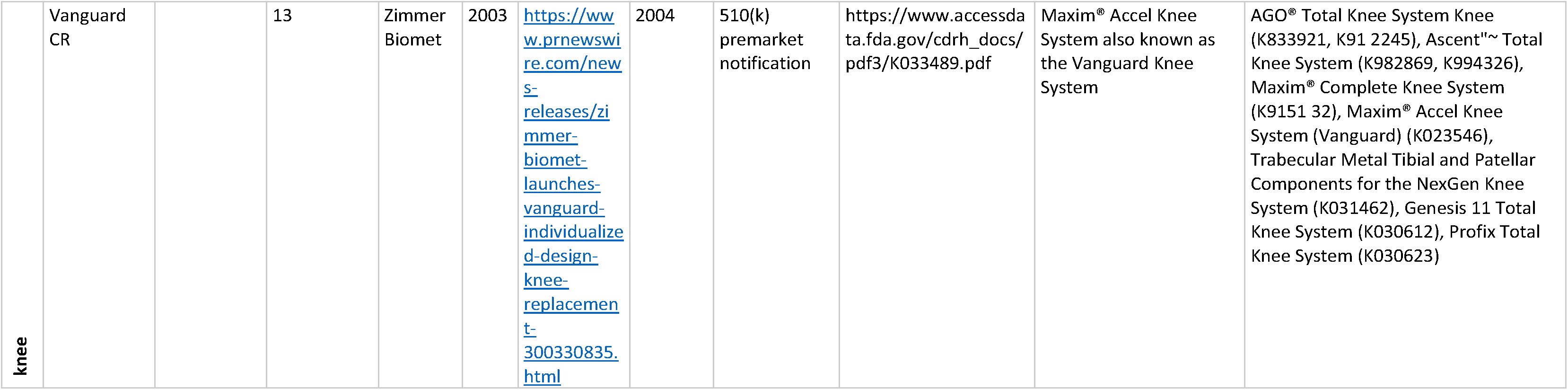

## References

1. MDCG 2020-6. Regulation (EU) 2017/745: Clinical evidence needed for medical devices previously CE marked under Directives 93/42/EEC or 90/385/EEC. A guide for manufacturers and notified bodies. April 2020.

2. Lübbeke A, Silman AJ, Prieto-Alhambra D, Adler AI, Barea C, Carr AJ. The role of national registries in improving patient safety for hip and knee replacements. BMC Musculoskelet Disord. 2017;18:414.

3. https://www.ema.europa.eu/en/documents/work-programme/european-collaboration-between-regulators-health-technology-assessment-bodies-joint-work-plan-2021_en.pdf.

4. Fraser A, Nelissen R, Kjaersgaard-Andersen P, Szymanski P, Melvin T, Piscoi P, CORE-MD investigators (see Appendix). Improved clinical investigation and evaluation of high-risk medical devices: the rationale and objectives of CORE-MD (Coordinating Research and Evidence for Medical Devices). EFORT Open Rev. 2021;6(10):839–849.

5. Hoogervorst LA, Geurkink TH, Lübbeke A, Buccheri S, Schoones JW, Torre M, Laricchiuta P, Piscoi P, Pedersen AB, Gale CP, Smith JA, Maggioni AP, James S, Fraser AG, Nelissen RGHH, Marang-van de Mheen PJ. Quality and reliability of clinical registries for the regulatory evaluation of medical device safety and performance across the implant lifecycle: a systematic review of European cardiovascular and orthopaedic registries. Manuscript under review.

6. Page MJ, McKenzie JE, Bossuyt PM, Boutron I, Hoffmann TC, Mulrow CD, Shamseer L, Tetzlaff JM, Akl EA, Brennan SE, Chou R, Glanville J, Grimshaw JM, Hróbjartsson A, Lalu MM, Li T, Loder EW, Mayo-Wilson E, McDonald S, McGuinness LA, Stewart LA, Thomas J, Tricco AC, Welch VA, Whiting P, Moher D. The PRISMA 2020 statement: An updated guideline for reporting systematic reviews.

7. International Society of Arthroplasty Registries (ISAR). International Prosthesis Benchmarking Working Group guidance document. May 2018. https://www.isarhome.org/publications, accessed 19 May 2021.

8. Kynaston-Pearson F, Ashmore AM, Malak TT, RombachI, Taylor A, Beard D, Arden NK, Price A, Prieto-Alhambra D, Judge A, Carr AJ, Glyn-Jones S. Primary hip replacement prostheses and their evidence base: systematic review of literature. BMJ 2013;347:f6956.

9. Aamodt A, Nordsletten L, Havelin L, Indrekvam K, Utvåg SE, Sundberg KH. Documentation of hip prostheses used in Norway. A critical review of the literature from 1996–2000. Acta Orthopaedica Scandinavica. 2004;75:663–676.

10. Chaverri-Fierro D, Lobo-Escolar L, Espallargues M, Martínez-Cruz O, Domingo l, Pons- Cabrafiga M. Primary total hip arthroplasty in Catalonia: What is the clinical evidence that supports our prosthesis? Rev Esp Cir Ortop Traumatol. 2017;61:139–145.

11. Naci H, Salcher-Konrad M, Kesselheim AS, Wieseler B, Rochaix L, Redberg RF, Salanti G, Jackson E, Garner S, Stroup TS, Cipriani A. Generating comparative evidence on new drugs and devices before approval. Lancet. 2020;395:986–997.

12. Hulstaert F, Neyt M, Vinck I, Stordeur S, Huić M, Sauerland S, Kuijpers MR, Abrishami P, Vondeling H, Flamion B, Garattini S, Pavlovic M, van Brabandt H. Pre-market clinical evaluations of innovative high-risk medical devices in Europe. Int J Technol Assess Health Care. 2012;28:278–284.

13. Cunningham BP, Harmsen S, Kweon C, Patterson J, Waldrop R, McLaren A, McLemore R. Have levels of evidence improved the quality of orthopaedic research? Clin Orthop Relat Res 2013;471:3679–3686.

14. Siljander MP, McQuivey KS, Fahs AM, Galasso LA, Serdahely KJ, Karadsheh MS. Current Trends in patient-reported outcome measures in total joint arthroplasty: A study of 4 major orthopaedic journals. J Arthroplasty. 2018;33:3416–3421.

15. Nelissen RG, Pijls BG, Kärrholm J, Malchau H, Nieuwenhuijse MJ, Valstar ER. RSA and registries: the quest for phased introduction of new implants. J Bone Joint Surg Am. 2011;93 Suppl 3:62–5.

16. https://journals.lww.com/jbjsjournal/Pages/Concise-Format-Guidelines.aspx

17. Malak TT, Broomfield JAJ, Palmer AJR, Hopewell S, Carr A, Brown C, Prieto-Alhambra D, Glyn-Jones S. Surrogate markers of long-term outcome in primary total hip arthroplasty: A systematic review. Bone Joint Res 2016;5:206–214.

18. Kärrholm J, Gill RH, Valstar ER. The history and future of radiostereometric analysis. Clin Orthop Relat Res. 2006 Jul;448:10–21.

19. Bohm ER, Kirby S, Trepman E, Hallstrom BR, Rolfson O, Wilkinson JM, Sayers A, Overgaard S, Lyman S, Franklin PD, Dunn J, Denissen G, W-Dahl A, Ingelsrud LH, Navarro RA. Collection and reporting of patient-reported outcome measures in arthroplasty registries: multinational survey and recommendations. Clin Orthop Relat Res. 2021;479(10):2151–2166.

20. Fraser AG, Byrne RA, Kautzner J, Butchart EG, Szymanski P, Leggeri I, de Boer RA, Caiani EG, Van de Werf F, Vardas PE, Badimon L. Implementing the new European Regulations on medical devices—clinical responsibilities for evidence-based practice. European Heart Journal 2020;41:2589–2596.

21. Wilkinson J, Crosbie A. A UK medical devices regulator’s perspective on registries. Biomed Tech (Berl). 2016;61:233–237.

22. Sedrakyan A, Campbell B, Merino JG, Kuntz R, Hirst A, McCulloch P. IDEAL-D: a rational framework for evaluating and regulating the use of medical devices. BMJ 2016;353:i2372.

23. Malchau H, Garellick G, Berry D, Harris WH, Robertson O, Kärrholm J, Lewallen D, Bragdon CR, Lidgren L, Herberts P. Arthroplasty implant registries over the past five decades: development, current, and future impact. J Orthop Res 2018;36:2319–2330.

24. Medical Device Clinical Evaluation Working Group. Post-Market Clinical Follow-Up Studies. International Medical Device Regulator Forum (IMDRF). 25 March 2021. https://www.imdrf.org/sites/default/files/docs/imdrf/final/technical/imdrf-tech-210325-wng65.pdf.

25. Derbyshire B, Prescott RJ, Porter ML. Notes on the use and interpretation of radiostereometric analysis. Acta Orthop. 2009;80:124–30.

26. Ochen Y, Gademan MG, Nelissen RG, Poolman RW, Leenen LP, Houwert RM, Groenwold RH. The potential value of observational studies of elective surgical interventions using routinely collected data. Ann Epidemiol. 2022;76:13–19.

27. Fraser AG. Post-market surveillance of high-risk medical devices needs transparent, comprehensive and independent registries. BMJ Surg Interv Health Technologies 2020;2:e000065.

